# What works for whom with telemental health? A rapid realist review

**DOI:** 10.1101/2022.03.21.22272706

**Authors:** Merle Schlief, Katherine R.K. Saunders, Rebecca Appleton, Phoebe Barnett, Norha Vera San Juan, Una Foye, Rachel Rowan Olive, Karen Machin, Prisha Shah, Beverley Chipp, Natasha Lyons, Camilla Tamworth, Karen Persaud, Monika Badhan, Carrie-Ann Black, Jacqueline Sin, Simon Riches, Tom Graham, Jeremy Greening, Farida Pirani, Raza Griffiths, Tamar Jeynes, Rose McCabe, Bryn Lloyd-Evans, Alan Simpson, Justin J. Needle, Kylee Trevillion, Sonia Johnson

## Abstract

**Background:** Telemental health (delivering mental health care via video calls, telephone calls or text messages) is increasingly widespread. Telemental health appears to be useful and effective in providing care to some service users in some settings, especially during an emergency restricting face-to-face contact such as the COVID-19 pandemic. However, important limitations have been reported, and telemental health implementation risks reinforcing pre-existing inequalities in service provision. If it is to be widely incorporated in routine care, a clear understanding is needed of when and for whom it is an acceptable and effective approach, and when face-to-face care is needed.

**Objective:** The aim of this rapid realist review was to develop theory about which telemental health approaches work, or do not work, for whom, in which contexts and through what mechanisms.

**Methods:** Rapid realist reviewing involves synthesising relevant evidence and stakeholder expertise to allow timely development of context-mechanism-outcome (CMO) configurations in areas where evidence is urgently needed to inform policy and practice. The CMOs encapsulate theories about what works for whom, and by what mechanisms. Sources included eligible papers from (a) two previous systematic reviews conducted by our team on telemental health, (b) an updated search using the strategy from these reviews, (c) a call for relevant evidence, including “grey literature”, to the public and key experts, and (d) website searches of relevant voluntary and statutory organisations. CMOs formulated from these sources were iteratively refined, including through (a) discussion with an expert reference group including researchers with relevant lived experience and front-line clinicians and (b) consultation with experts focused on three priority groups: 1) children and young people, 2) users of inpatient and crisis care services, and 3) digitally excluded groups.

**Results:** A total of 108 scientific and grey literature sources were included. From our initial CMOs, we derived 30 overarching CMOs within four domains: 1) connecting effectively; 2) flexibility and personalisation; 3) safety, privacy and confidentiality; and 4) therapeutic quality and relationship. Reports and stakeholder input emphasised the importance of personal choice, privacy and safety, and therapeutic relationships in telemental health care. The review also identified particular service users likely to be disadvantaged by telemental health implementation, and a need to ensure that face-to-face care of equivalent timeliness remains available. Mechanisms underlying successful and unsuccessful application of telemental health are discussed.

**Conclusions:** Service user choice, privacy and safety, the ability to connect effectively and fostering strong therapeutic relationships, need to be prioritised in delivering telemental health care. Guidelines and strategies co-produced with service users and frontline staff are needed to optimise telemental health implementation in real-world settings.

## Introduction

Telehealth is defined as “the delivery of health-related services and information via telecommunications technologies in the support of patient care, administrative activities, and health education” [3]. Telemental health refers to such approaches within mental healthcare settings. It can include care delivered by means such as text messaging and chat functions, but most commonly refers to telephone calls and video calls, which are central to telemental health care.

Before the COVID-19 pandemic, there was interest in many countries and settings in integrating new technologies, including telemental health approaches, more widely and effectively in mental healthcare services. This was of particular interest in countries where face-to-face (i.e., in person) mental health care was largely inaccessible for remote communities [4]. Research has demonstrated that telemental health can be successful in various contexts, although studies prior to the pandemic tended to relate to relatively small-scale and well-planned applications of telemental health with volunteer participants rather than large-scale implementation across whole service systems. Telemental health has been found to be effective in reducing treatment gaps and improving access to mental health care for some service users [5–7]. This includes those who live far from services or where caring responsibilities affect their ability to travel [8–10]. Positive outcomes and experiences have been reported across a range of populations (including adult, child and adolescent, older people and ethnic minority groups) and settings (including hospitals, primary care and community) [11–13]. Some evidence has suggested that telemental health modalities such as videoconferencing are equivalent to, or even better than, face-to-face in terms of quality of care, reliability of clinical assessments, treatment outcomes, or adherence for some service users [11, 12, 14, 15]. High levels of service user acceptance and satisfaction with telemental health services have also been reported in research samples [6], and for certain populations, including those with physical mobility difficulties, social anxiety or severe anxiety disorders [8, 9]. Conversely, however, telemental health services are not appropriate for all service users, and there is no one-size-fits-all approach. In particular, service users experiencing social and economic disadvantages, cognitive difficulties, auditory or visual impairments, or severe mental health problems, such as psychosis, have benefitted less from telemental health interventions [2, 16]. Digitally excluded service users tend to be people who are already experiencing other forms of disadvantage and already at risk of poorer access to services and less good quality care: a switch to telemental health may thus exacerbate existing inequalities [17, 18]. Additionally, concerns have been raised around impacts of telemental health on privacy and confidentiality of clinical contacts, especially for the many service users who do not have an appropriate space and facilities for its use, as well as its appropriateness for certain purposes, such as conducting assessments or risk management [2].

Encouraging evidence of telemental health acceptability and effectiveness from pre-pandemic research tended to relate to limited populations who had opted into well-planned remote services [1]. However, during the COVID-19 pandemic, the use of telemental health around the world greatly accelerated and telemental health became a routine approach for maintaining and delivering mental health services. Telemental health initiatives were central to delivering mental health services in the context of this emergency. Technological initiatives have also helped to address social isolation, which worsened throughout the pandemic [8, 19]. In the UK, there were large increases in remote consultations in National Health Service (NHS) primary care [20] and national data reported that most contacts in NHS mental health settings were delivered remotely in 2020 [21], particularly during the first UK lockdown (March to July 2020).

Following the rapid adoption of telemental health at the start of the crisis, service planners, clinicians and service users have expressed interest in greater use of telemental health long-term [2, 19, 22, 23]. However, several challenges have been identified as arising from this widespread implementation [2, 17, 19, 24, 25]. These include i) reaching digitally excluded populations, who may, for example, have limited technological access and/or expertise, thus compounding existing inequalities experienced by disadvantaged groups; ii) a lack of staff competence in using telemental health devices and confidence in delivering telemental health care; iii) a lack of technological infrastructure within health services; iv) challenges in managing clinical and technological risks in remotely delivered care; v) developing and maintaining strong therapeutic relationships online, especially when the first contact is remote rather than face-to-face; vi) maintaining service user safety and privacy; and vii) delivering high quality mental health assessments without being able to see or speak to the service user face-to-face. It is also more difficult to undertake physical assessments, including of physical signs linked to mental health, and side effect monitoring.

Both for future emergency responses and to establish a basis for the integration of telemental health into routine service delivery (where appropriate) beyond the pandemic, evidence is needed on how to optimise telemental health care given the unique relational challenges associated with mental health care, and to identify what works best for whom in telemental health care delivery in which contexts. It is also important to identify contexts in which telemental health is unlikely to be safe and effective, where face-to-face delivery should remain the default.

A methodological approach developed to address questions of which interventions work for whom in which contexts in a timely way is the rapid realist review (RRR) [26]. This methodology has been developed to rapidly produce policy-relevant and actionable recommendations through a synthesis of peer-reviewed evidence and stakeholder consultation. A key characteristic of realist methodology is the focus on interactions between contextual factors (for example, a certain population, geographical location, service setting or situation) and relevant mechanisms (for example, behavioural reactions, participants’ reasoning and/or resources), which impact on the outcomes of interest, for example, intervention adherence or service user satisfaction [26–28]. Together, these are used to develop context-mechanism-outcome (CMO) configurations, which comprise the fundamental building blocks of realist synthesis approaches. Evidence from the wider literature is also drawn upon to develop mid-range theories, which help to elaborate and refine the developed CMOs by shedding further light on how their mechanisms operate [26, 29–31].

This is a unique opportunity to establish the characteristics of high quality telemental health services and to use these findings to identify key mechanisms for acceptable, effective, and efficient integration of telemental health services into routine mental health care. Employing a realist methodology, we aim in this RRR to answer the question of what telemental health approaches work for whom, in which contexts and how? Specifically, we investigate in this review: 1) What factors or interventions improve or reduce adoption, reach, quality and acceptability, or other relevant outcomes in the use of telemental health in any setting? 2) Which approaches to telemental health work best for which staff and service users in which contexts? 3) In what contexts are phone calls, video calls or text messaging preferable, and in which contexts should mental health care be delivered face-to-face instead? We focus particularly on groups and contexts identified as high priority by policymakers (process described in detail in the methods section), including 1) children and young people, 2) crisis care and inpatient settings, and 3) groups at high risk of digital exclusion: examples from these groups are included wherever possible.

## Methods

The RRR was conducted by the NIHR Mental Health Policy Research Unit (MHPRU), a team established to deliver evidence rapidly to inform policymaking, especially by the Department of Health and Social Care in England and associated government departments and NHS policy leadership bodies. The project constitutes the final stage in a programme of work on telemental health delivery carried out to meet urgent policy need, which included an umbrella review of pre-COVID evidence [1], a qualitative investigation of service user experiences of telemental health [24], a systematic review of literature on telemental health adoption conducted during the early phase of the pandemic [2] and a systematic review on the cost-effectiveness of telemental health approaches (personal communication by Clark et al., 2022). This RRR was registered on PROSPERO (CRD42021260910).

We conducted the RRR during the COVID-19 pandemic, with video conferencing as the primary means of communication among the research team.

### Study design

An expert reference group of 23 people, including academics, experts by experience (lived experience researchers from the MHPRU Lived Experience Working Group with personal experiences of using mental health services and/or supporting others) and by profession (including frontline clinicians), guided and contributed to the RRR throughout. The group met weekly throughout this process from July until November 2021. The expert reference group meetings served to develop and refine the study protocol, plan the searches for evidence (particularly the targeted additional searches supplementing the initial planned strategy), iteratively examine, refine, and validate the CMOs derived from our evidence synthesis, with reference to their expertise by experience and/or profession, and to plan wider consultation on our emerging findings. Members of the expert reference group also contributed to the literature searches, data extraction, synthesis, and interpretation of data.

The stages of our RRR were based on the following five steps, variations of which have been described and used in previous studies [26, 30, 31]:

1. Developing and refining research questions
2. Literature searching and retrieving information (data/stakeholder views)
3. Screening, appraising and extracting information/data
4. Synthesising information/data
5. Interpreting information/data

Our approach to these steps was iterative rather than linear, particularly for steps three, four and five, where there were multiple phases of extraction, synthesis, and interpretation. This is described in detail below.

### Developing and refining the research question

We formulated the research question in response to policymaker need. We reviewed findings from earlier stages of the MHPRU’s programme on COVID-19 impact on mental health care and on telemental health [1, 8, 19, 24, 32] with policymakers, including senior officials and mental health teams in the Department for Health and Social Care, NHS England and Public Health England. We then identified questions to be addressed from their perspective to plan for future implementation and delivery of telemental health. This included considering how best to incorporate telemental health in routine practice once the need for its emergency deployment passes. Early in these discussions, three priority groups about which evidence is currently lacking were identified as especially important for policy and planning: children and young people, users of inpatient and crisis care services, and digitally excluded groups. Our primary question of which telemental health approaches work for whom in what context originated in these discussions and was further refined by the MHPRU core research team, who identified a rapid realist review methodology as appropriate, and further refined the primary and secondary questions and methodology with the expert reference group before registering the protocol.

### Selection criteria

Sources were included if they met the following criteria:

#### Participants

Staff working in the field of mental health, people receiving care from mental health services, family members and other supporters of people receiving mental health care.

#### Interventions

Any form of remote (spoken or written) communication between mental health professionals, or between mental health professionals and service users, family members and other supporters, using video calls, telephone calls, text messaging services, or hybrid approaches combining face-to-face and online modalities. Peer support communications were also included alongside any strategies or training programmes to support the implementation of the above. Self-guided online support and therapy programmes were excluded.

#### Types of evidence

Qualitative or quantitative evidence on: 1) what improves or reduces adoption, reach, quality, acceptability, or clinical outcomes in the use of telemental health; 2) impacts of introducing interventions or strategies intended to improve adoption, reach, quality, acceptability or clinical outcomes; 3) interventions or strategies intended to help mental health staff make more effective use of telemental health technologies; 4) impacts of telemental health on specific service user groups and settings, including people who are digitally excluded, users of inpatient and crisis care services, and children and young people; and 5) the appropriateness of the use of telemental health versus face-to-face care in particular contexts. As well as outcomes, sources were required to include information on mechanisms (i.e., what works for whom and how).

#### Study design

Any qualitative, quantitative, or mixed-methods study design, including relevant service evaluations, audits, and case series. Grey literature and other sources, such as websites, stakeholder feedback and testimonies from provider organisations and service user and carer groups. Sources were also included if the focus was not solely remote working but the results contained substantial data relevant to our research questions. Editorials, commentaries, letters, conference abstracts and theoretical studies were excluded.

### Search strategy

The search strategy was in accordance with PRISMA guidelines (Figure 1) [33]. Resources and literature were identified through the following sources:

1. We screened peer-reviewed studies included in two previous reviews on telemental health conducted by the MHPRU. The umbrella review by Barnett et al. (2021) included systematic reviews, realist reviews, and qualitative meta-syntheses on remote working before the onset of the COVID-19 pandemic [1]. The systematic review by Appleton et al. (2021) synthesised primary research on the adoption and impacts of telemental health approaches during the pandemic [2]. An updated search of the latter review was conducted on 19/05/2021.
2. We worked with our expert reference group to identify additional peer-reviewed and grey literature. Searches were conducted of the websites of relevant national and international voluntary and statutory organisations identified by the expert reference group and by internet searches (for example, Mind and the Royal College of Psychiatrists). Identified literature was noted on a shared Microsoft Excel spreadsheet. This process was supported using Slack, an online messaging application, to coordinate this complex and rapidly changing process.
3. Lastly, the MHPRU disseminated a call for evidence via Twitter and email to relevant organisations and individuals (such as charities supporting digital inclusion, chief information officers and telehealth leads within NHS trusts) inviting them to submit relevant evidence, including evaluations, audits, surveys, stakeholder feedback and testimonies from provider organisations and service user and carer groups.

**Figure 1.**
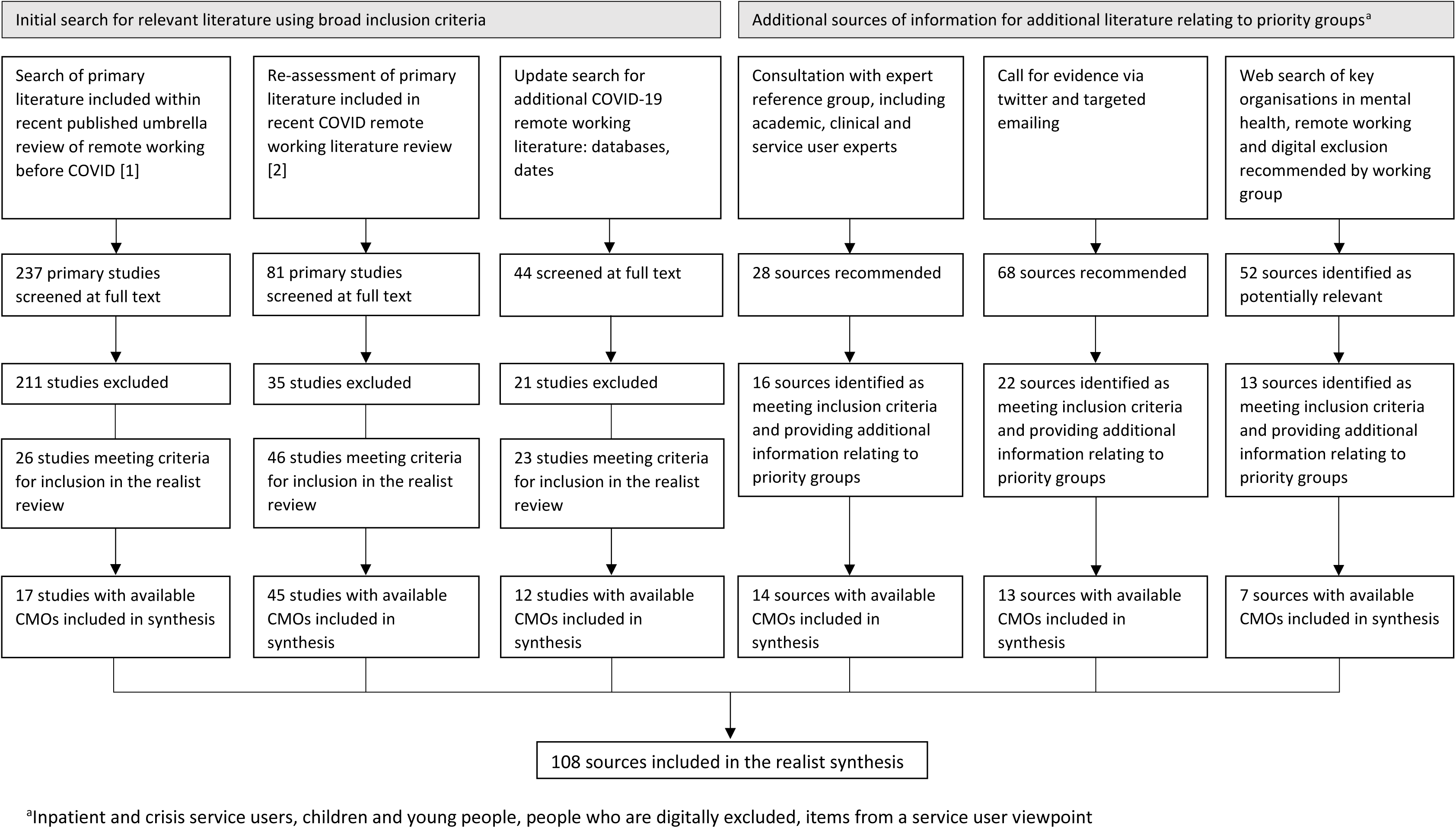
Prisma Diagram.

### Study selection

References included in the umbrella and systematic review were downloaded and screened for inclusion in the RRR using EPPI-Reviewer 4.0 [34]. One reviewer screened abstracts and titles of the references identified through the updated searches of the umbrella review and systematic review. Full texts were reviewed for inclusion, with included and ‘unsure’ sources checked by another reviewer. Sources were included in the final review if they met our inclusion criteria and provided relevant information for the development of CMOs. Disagreements were resolved through discussion with the wider research team.

### Process of data extraction, synthesis and interpretation

#### Data extraction

The source characteristics were extracted and input on EPPI-Reviewer 4.0 [34]. Extracted characteristics included study aim and design (if applicable), type of service, telemental health modalities employed, mental health diagnosis of service users, and staff occupation. MHPRU researchers screened each included source for information that could be assembled into CMOs relating to telemental health, i.e., information on contexts, outcomes as well as underlying mechanisms. Underlying CMOs were extracted by MHPRU researchers and LEWG members. Each week, samples of the extracted CMOs were reviewed by the expert reference group to ensure coherence, relevance, validity, and format consistency.

#### Data synthesis

The research team then began the process of synthesising the underlying CMOs by reviewing the extracted CMOs and identifying emerging themes. We developed four domains to encapsulate key aspects of the evidence: 1) connecting effectively; 2) flexibility and personalisation; 3) safety, privacy and confidentiality; and 4) therapeutic quality and relationship. Each of the four identified domains was allocated to a MHPRU researcher to lead on the synthesis, with input from LEWG members, clinicians, and MHPRU senior researchers.

To develop content for each of these four domains, underlying CMOs were first reviewed in terms of their similarities and differences, and then grouped together based on similar mechanisms and outcomes. Each group of CMOs was then synthesised and refined to create a single overarching CMO, which reflected key content across the underlying CMOs which contributed to it. Each overarching CMO was assigned to one of the four domains.

#### Interpretation

An iterative process of revising and refining overarching CMOs from the perspective of stakeholder experience followed. Revisions, refinements and additions were first made through discussion with the expert reference group. Summaries were then also discussed at three two-hour stakeholder webinars each focusing on one of our priority groups: children and young people, inpatient and crisis care services, and digitally excluded groups. The webinars were primarily attended by groups representing these constituencies and services who work with them, including experts and stakeholder representatives from research, policy and clinical settings (nationally and internationally), the voluntary sector, as well as representatives of lived experience groups and community organisations working with marginalised groups, and from telehealth technology initiatives. There were between 30-40 participants at each webinar. During the webinars, participants were divided into breakout rooms, with a facilitator and a note taker from the core research team. High-level summaries of preliminary data were presented by domain and attendees were asked to discuss the following questions: whether the preliminary summaries captured their own knowledge and experience of telemental health; whether/how the summaries applied to/were relevant for the priority group at hand; and whether they were aware of any additional challenges or recommendations related to delivering telemental health to the priority group.

Based on the feedback from these webinars, the overarching CMOs within each of the four domains were then further revised and refined. We actively sought additional information relating to each overarching CMO, including relevant contexts, further detail about mechanisms, real-life examples of strategies and solutions (such as for overcoming barriers identified within the CMO), and points of particular importance or concern, from the webinar notes, the expert reference group meetings and related literature. We noted this information alongside the relevant overarching CMO and used it to refine the CMOs. Additionally, we drew upon mid-range theories (evidence-based theories derived from the wider literature) to provide more theoretically informed explanations of mechanisms. Throughout this process, the core research team and the expert reference group were iteratively consulted, and their feedback integrated into the overarching CMOs. The revised theories were shared for a final email consultation with the stakeholders who we invited to our webinars. Their feedback was incorporated and resulted in the final overarching CMO models presented under each domain in this paper.

## Results

Underlying CMOs were extracted from 17 of the studies included in the previous umbrella review [1] and from 45 studies included in the systematic review [2]. The updated search yielded 44 potentially relevant studies, of which 21 were excluded. CMOs were extracted from 12 of the remaining 23 studies that met our inclusion criteria and were included in the realist synthesis. Through consultations with our expert reference group, we identified 28 sources, of which 16 met our inclusion criteria and provided additional information relating to our priority groups, and 14 yielded CMOs that were included in the synthesis. We received 68 potential sources through the call for evidence, of which 22 met our inclusion criteria and provided relevant information on our priority groups. CMOs were extracted from 13 of these. Lastly, website searches identified 52 potentially relevant sources. Thirteen of these met our inclusion criteria and seven provided information relevant for CMOs. The realist synthesis includes a total of 108 sources.

Of the 108 included sources with primary data or detailed accounts of what works for whom and in what context, most were primary research studies (n=72), followed by service descriptions/ evaluations/audits (n=19), guidance documents (n=4) and briefing papers (n=3), commentaries/ editorials/discussions (n=4) and letters (n=2), as well as one review (n=1), a news article (n=1), one webpage (n=1) and one service user led report (n=1). Of the sources that included primary research data, 32 sources employed quantitative, 19 qualitative, and 33 mixed methods (including two case studies).

The majority of sources were published in the USA (n=41) and the UK (n=34). The remaining sources collected data in Canada (n=7), the Dominican Republic (n=1), Australia (n=7), China (n=2), India (n=3), Egypt (n=1), Nigeria (n=1), as well as ten European countries including Austria (n=1), France (n=1), Germany (n=1), Ireland (n=1), Italy (n=2), Netherlands (n=1), Portugal (n=1), Spain (n=1), Sweden (n=1), and Switzerland (n=1). Details of the included sources are presented in Appendix 1.

Overarching CMOs for each domain are summarised in Tables 2 to 5, and details of the underlying CMOs and summary notes on stakeholder discussions also shaping the overarching CMOs are in Appendix 2. For each overarching CMO, we include in Tables 2 to 5 notes on key contexts to which overarching CMOs are particularly relevant and examples of strategies and solutions addressing the challenges or opportunities identified in the CMO: these are drawn from underlying CMOs and stakeholder discussions. In the text outlining each domain, we also identify major mid-range theories that elucidate mechanisms and outcomes for overarching CMOs.

**Table 2.** Domain 1. Connecting Effectively.

| CMO title | References | Overarching CMO | Key Contexts | Example strategies and solutions |
| --- | --- | --- | --- | --- |
| CMO 1.1: Providing access to digital devices | [19, 49-59] | When service users who do not have access to digital devices are given access to up-to-date devices (and chargers), paid for or loaned to them (C), this results in improved access to and implementation of telemental health services (especially via video platforms) (O1), some inequalities in accessing a digital device are addressed (O2), exacerbation of existing inequalities is less likely (O3), and service users are more able to maintain personal contact with family and friends if they wish, and to access a range of online services (O4), as this reduces the burden of having to purchase a device for the service users, and provides more financially viable access to devices required for online connections (M). | Lack of access to devices particularly affected people living in poverty or with unstable living circumstances (such as homeless people or refugees), as well as other groups at risk of exclusion not only from telemental health but from a range of services and networks (such as people who are cognitively impaired, or with psychosis or substance abuse disorders). It can also particularly affect inpatients, who may not have access to devices and/or charging facilities on the ward, and children and young people, who may not have access to their own device. | 1) Schemes organised by bodies including healthcare providers, libraries, schools and colleges, charities and community organisations that lend or give digital devices and chargers; 2) Inpatient wards providing devices such as iPads and/or short cable or wireless chargers, or charging lockers; 3) Health services providing separate and private rooms with video conferencing capabilities. 4) Promoting awareness among service providers of the tendency for digital exclusion to exacerbate existing disadvantage and inequalities, with development of active strategies to mitigate this. 5) Phone calls rather than video calls may be more appropriate for service users who do not have access to digital devices which facilitate video calls. |
| CMO 1.2a: Lack of access to stable, secure, and/or adequate internet connection | [19, 49, 51, 52, 54, 57, 60-70] | When staff deliver telemental health via video from workplaces or homes with unstable and poor internet connection (C), tele-consultations are difficult (or impossible) to conduct with service users (O1), fewer tele-consultations are conducted (O2) and telemental health is viewed less positively (O3), as staff experience frustration and there is reduced motivation to arrange online appointments (M). | This is particularly relevant for staff working within those healthcare providers that frequently have insufficient Wi-Fi or internet connection to deliver sessions smoothly. It also affects staff who do not have adequate Wi-Fi connectivity in their own homes or when working in the community. | 1) Investing in high quality IT infrastructure to ensure disruptions to calls do not originate from poor provider connections; 2) providing devices with access to data, or data/Wi-Fi allowances, for use by staff when working away from health service premises (at home or in community settings). |
| CMO 1.2b: Lack of access to stable, secure, and/or |  | When service users only have access to an insecure, unstable and poor-quality internet connection, and/or consistent technological problems (C), it is difficult for telemental health to be viewed positively (O1) they are able and/or willing to accept fewer online consultations (O2), | This is especially relevant for service users on low incomes or from socially marginalised groups (such as homeless people), those living in multiple occupancy households where Wi-Fi is overstretched, and | 1) Signposting to low-cost plans for people on low incomes; 2) providing free access to Wi-Fi, data and phone connections (may be included with devices that are lent or given); 3) providing or signposting to community hubs with access to data; 4) providing face- |
| adequate internet connection |  | may continue to struggle with their mental health (if face-to-face consultations are unavailable) (O3) and this may result in digital exclusion which could exacerbate existing inequalities (O4), as the service users struggle to engage in sessions with sufficient clarity and mutual comprehension, and experience frustration (M). | people from Low-to-Middle Income countries. Lack of access to reliable internet or even electricity also differentially affects people in rural and remote areas. People in marginalised groups may also lack the means to pay for a telephone service. | to-face appointments where the above problems cannot be resolved. |
| CMO 1.3a: Benefits of providing support and guidance for using technology | [19, 24, 45, 52-54, 56, 63, 71-85] | When staff who lack the confidence and/or knowledge to deliver mental health care online (particularly via video calls) receive practical instruction and guidance on how to use technology to deliver mental health services, including clear information about how to operate within local policies, procedures and platforms, troubleshoot issues during telemental health sessions, and formulate and implement back-up plans (C), they feel an increased sense of confidence in managing and delivering telemental health services (M), which leads to increased use of telemental health services (O1) and fewer delays, resulting in more appointments being completed on time (O2). | This is especially relevant for staff who are new to delivering mental health care remotely, or who are unclear or unfamiliar with using locally recommended platforms and procedures. | 1) Provision of training sessions relevant to local context, including information on troubleshooting technology and maintaining privacy, safety and confidentiality; 2) access to guidance on video calls, including clear information on processes and policies for providing telemental health in the organisations where staff work; 3) peer support and group or team sessions for practising technology; 4) refresher training session and rolling training for new joining staff. |
| CMO 1.3b: Benefits of providing support and guidance for using technology |  | When service users with access to a technology device who struggle with the confidence, knowledge and/or ability to use telemental health receive guidance, reassurance and instruction (tailored to their healthcare provider and to their language, reading ability and any sensory disability) on how to use technology (particularly video calls) to access mental health care, engage with back up plans, and receive timely technical support and troubleshooting during treatment sessions (C), they feel an increased sense of confidence in accessing telemental health (M), which reduces anxiety using telemental health | Receiving guidance tailored to local policies and procedures for telemental health access is relevant to all service users. It is likely to be particularly relevant to groups identified as at high risk of digital exclusion through lack of confidence in using technology, including older people, people with severe mental health problems and people with intellectual disabilities or cognitive impairments. Production of accessible guidance is especially | 1) One-to-one training or support from a digital facilitator/champion/mentor, peer supporters, family, friends, or a dedicated member of staff who can provide guidance; 2) written or video information about how to access telemental health, including material tailored to age, cognitive abilities, sensory impairments, and language, and to the platforms that are in local use (for example, guidance in other languages, easy-read material with pictures; personalised workbooks sent to children before a telemental health contact); 3) opportunities |
|  |  | and digital technologies (including in their personal lives) (O1), facilitates adoption of and adherence to telemental health (O2), improves service users' ability to adjust to remote care (O3), reduces interruptions in care delivered via telemental health (O4) and increases satisfaction with telemental health (O5). | relevant for people with cognitive impairments or intellectual disabilities, children, people with sensory disabilities and people who do not understand English well. | for practice and repetition of training as needed; 4) written guides for different technology platforms that give step-by-step instructions, including for how to set up IDs and passwords, sent out with appointment letters; 5) brief and direct training sessions offered to service users before their online appointment, doing a trial run of using the technology beforehand, or increasing virtual contact duration to accommodate learning. |
| CMO 1.4:<br>Impact of technology related disruptions | [38, 55, 66, 86-92] | When technological issues (including connection problems and device issues) lead to disruptions to online sessions and there is no pre-arranged back-up method of contact (e.g. a plan to connect by telephone instead of video-call if needed) (C), the quality of the intervention is diminished (O1) there is a loss of empathic connection between client and therapist (O2) and the sessions may not be able to continue (O3), as the flow of the conversation is interrupted and session time reduced, for example, when having to ask the other person to repeat what has been said, or when cut off completely, leaving staff and service users potentially feeling distracted, frustrated, awkward and upset (particularly if there is a threat of therapy withdrawal due to missed sessions) (M). | This is particularly relevant for the use of videoconferencing, where phone back-up can reduce risk of abandoning appointments due to failure to make a stable connection. | 1) Agree a back-up method of connecting, e.g., reverting to telephone calls in the event of disruption; 2) where connection issues cannot be readily resolved, and back up methods are not sufficient, strategies to address connection problems (as above) are required or moves to face-to-face care should be facilitated. |
| CMO 1.5:<br>Preparing service users for telemental health | [93-95] | When staff prepare service users for telemental health appointments and communicate clearly with service users about what to expect (C), this leads to more accepted calls and fewer missed service user contacts (O), because service users have more relevant knowledge of the process, including when to expect contacts and from what number, and feel more comfortable engaging with telemental health services (M). | This is relevant across telemental health contexts but may be particularly relevant for service users who have not used telemental health before, or are anxious or worried about telemental health. | 1) Informing service users when they are going to be contacted and via what platform; 2) sending texts to remind a service user that a call is scheduled for a specific time; 3) informing service users about when phones are manned and how long they can expect to wait for a call back; 4) routing staff telemental health calls to service users through unblocked, local and familiar clinic or hospital numbers, or if a private number has to be used, ensuring that service users are expecting this. |
| CMO 1.6:<br>Familiarity with platform and ease of use | [51, 76, 96-100] | Where service users already use remote technologies for social, educational or work purposes, and/or where the online platforms are relatively easy to use (C), offering a choice of familiar and accessible technology platforms that may be less difficult and time-consuming for staff and service users to understand or learn (M) may increase the likelihood of engagement with services via telemental health, especially video calls (O). | Especially relevant to service users who are already making some use of technology, for example for social, educational or work purposes. | 1) Service providers prioritising allowing use of a variety of platforms, especially those likely to be widely familiar (e.g. Zoom and WhatsApp video); 2) attempting to address governance concerns and provide guidance for safe use, balancing online safety and other risks such as disengagement from services. |
| CMO 1.7:<br>Telemental health may be a better alternative to receiving no care during an emergency, such as the COVID-19 pandemic | [38, 53, 70, 100-102] | When service users are offered telemental health appointments because face-to-face appointments are restricted (e.g. due to COVID-19 or another emergency) (C), these appointments are likely to be accepted by some service users on the basis that they are the main way by which mental health care can continue (O1) and there is a reduced risk of infection from COVID-19 (O2), as face-to-face options are lacking or very restricted and telemental health is seen as preferable to receiving no support at all and as an alternative to cancelling appointments entirely (M). | This is applicable across mental health services in the context of any emergency which restricts face-to-face meetings, especially in services where all face-to-face contacts have been discontinued or where they are limited to immediate crises. It is especially relevant for service users who need to self-isolate because of high personal risk, or where clinicians need to self-isolate after contact with others. | 1) Widespread implementation of telemental health has proved a successful strategy for maintaining contact with many mental health service users, but not all, during an emergency that restricts face-to-contact, with some service users accepting telemental health in this situation who would be reluctant or unable to do so if not in a relatively short-term emergency; 2) Some continuing face-to-face contacts are still required if care is to be offered to all who need it. |

### Domain 1. Connecting Effectively

The content of this domain relates to establishing a good online connection to join a video call of sufficient quality, or to engage in telemental health via phone or message, with a particular focus on digital exclusion. Table 2 outlines seven overarching CMOs identified in relation to this domain, addressing issues concerning device and internet access (1.1, 1.2), technology training (1.3), the impact of preparation and technological disruptions (1.4, 1.5), the familiarity and usability of the platforms (1.6) and the acceptability of telemental health as an alternative to receiving no care during emergency situations (1.7). Three of the CMOs related to trying to resolve three main challenges: i) access to a charged, up-to-date device that enables internet access (1.1); ii) an internet (Wi-Fi or data) or signal connection (1.2); and iii) the knowledge, ability and confidence to engage online (1.3). Much of the content relates to challenges service users encounter in engaging with telemental health, but the literature and stakeholder discussion also yielded significant challenges for staff and service providers in the practicalities of connecting online.

Theories regarding the relationship between digital exclusion with other forms of exclusion and deprivation, and the potential of digital exclusion to amplify inequalities, contribute to our understanding of key mechanisms and outcomes in this domain. Widening inequalities have been described as an inevitable consequence of expansion of the role of technology in healthcare, with loss of access to community facilities such as libraries making this a still greater risk during the pandemic [35]. The “digital inverse care law” [17] describes a tendency for groups in most need of care (for example older people or people experiencing social deprivation) to be least likely to engage with technological forms of healthcare. This is highly salient in mental health care, given the strong associations between experiencing mental health challenges and experiencing one or, often, many forms of disadvantage [35].

Access to devices (1.1) is one contributor to digital exclusion, and groups who are especially likely to be affected include homeless individuals and people living in poverty, those receiving inpatient or crisis care, and young children who may not have their own devices. The type of device may be important for accessing telemental health. For example, smartphones may be less suitable for video therapy due to their small screens [36], although this may be less relevant for young people who are familiar with and consistently use smartphones for connecting online [37]. This raises future research questions around which types of digital device work for whom in what context when it comes to continuing telemental health treatment. It may also have implications for the provision of suitable equipment to certain populations.

Our consultations and the wider literature revealed that lack of access to good quality Wi-Fi, including poor Wi-Fi in hospitals and offices, was a further key barrier to successful and equitable delivery of telemental health (1.2). It was emphasised that modernisation of software and hardware, particularly within the NHS, is needed in many healthcare sites to allow for the requirements of telemental health. Service users also reported relying on their own mobile phone data to connect to telemental health services, often depleting their data completely after or during just one video call or consultation, which is expensive to replenish and may also deter engagement. This could amplify existing inequalities, leaving some service users at risk of digital exclusion and unable to access the internet and mental health support. Disruptions to telemental health appointments due to poor connection are a significant barrier to engagement (1.4). Our consultations and the existing literature highlighted the importance of having an alternate form of communication (for example, a telephone call) as a back-up plan in case of a technology or connection failure [38–41].

We identified the importance of technology training and sustained formal and informal support for service users (1.3b). Variations in ability to use telemental health are likely to disproportionately affect certain service user populations, often groups who also experience high levels of need for mental health care and inequalities in its provision. This includes people living in deprived circumstances, people with cognitive difficulties, people with paranoia or who do not speak the same language as service providers. Understanding how to use technology is also important for service users’ social engagement and connection, which is relevant for wider recovery and citizenship [42–44]. Young children may also be disproportionately affected as they may not be able to resolve difficulties they experience during telemental health sessions without the help of their parents or other supporters.

For staff (1.3a), our evidence suggests that staff training provided more widely and accessibly on using technology for telehealth would be helpful to ensure high quality service provision and overcome barriers around staff not having time allocated to training or being reluctant to ask for support. Evidence suggests that there are also benefits of having access to technical support to troubleshoot issues during sessions [45], and practising new skills and learning with colleagues and peers [46].

The importance of the familiarity and usability of platforms was highlighted throughout our stakeholder consultations and weekly reference group meetings, as well as in the published literature (1.6), in keeping with previous research on acceptability of telemedicine by service users [47]. Many of the platforms and devices commonly used for telemental health services, for example during the COVID-19 pandemic, were not designed for use in health care settings, and therefore may be less user-friendly. The importance of usability is emphasised by Nielsen (1993): three of five main usability attributes of a programme are that it should be easy to learn, efficient to use and easy to remember [48]. This is in keeping with our findings related to familiarity and usability in telemental health.

Finally, preparing service users for telemental health sessions was key (1.5). This was relevant across telemental health contexts, but information tailored to individual communication needs may be especially helpful for service users who may experience additional challenges connecting online (for example, those who are inexperienced with technology or anxious about using telemental health, young children, older people, and people with cognitive difficulties). For people with significant sensory impairments, specialist adaptations will need to be available if telemental health is to be a viable modality e.g., mobile phones that flash when receiving a call, or providing guidance in Braille or sign languages.

### Domain 2. Flexibility and Personalisation

Table 3 presents the CMOs, key contexts, and example strategies and solutions for the domain flexibility and personalisation. The need for flexibility and personalisation was a key theme identified in both the literature and stakeholder consultations when considering using telemental health in place of (or in conjunction with) face-to-face mental health support. A total of eight over-arching CMOs were identified in this domain, which can be divided into three main categories: taking individual preferences into account (2.1, 2.5, 2.7), convenience (2.2), and allowing for more collaborative and potentially specialised care (for example, involving specialists, family or friends in care) (2.3, 2.4, 2.6, 2.8).

**Table 3.** Domain 2. Flexibility and Personalisation.

| **CMO title** | **References** | **Overarching CMO** | **Key Contexts** | **Example strategies and solutions** |
| --- | --- | --- | --- | --- |
| CMO 2.1: Taking individual preferences into account – offering alternatives | [24, 38, 58, 60, 75, 77, 85, 87, 100, 101, 111-123] | When services using remote mental health care allow service users to choose the modality of telemental health and/or a choice of remote versus face-to-face care, and regularly check their preferences (C), this allows service users to have greater autonomy and choice (M) leading to them feeling more satisfied with and able to engage with the type of care received (O1), leading to improved uptake (O2), and improved therapeutic relationships with their clinician (O3). | Allowing service user choice and delivering services flexibly is a key principle across settings and populations, with the overall aim that care of equivalent quality should be available in a timely way whatever modality is chosen. Hybrid care, with a flexible mixture of face-to-face and telemental health based on the purpose or function of appointment (e.g. prescription review versus first visit to see clinician), preference, and circumstances is especially relevant to service users receiving relatively complex care with multiple types of appointments, for example, from multidisciplinary community teams. Children and young people may particularly benefit from being offered choice as it increases their feelings of autonomy and improves engagement in care. | 1) Initial conversations about telemental health with all service users, in which their preferences regarding the mode of appointments and their access to and expertise and interest in using technology are explored (a shared decision making tool could be used to structure this); 2) ensuring clinicians making collaborative plans with service users for telemental health use are aware of risk factors for difficulty engaging with telemental health and digital exclusion, including individual difficulties and wider contextual factors, such as poverty and poor or shared housing; 3) ensuring equal access to timely care of good quality regardless of choice of modality; 4) regularly revisiting preferences and collaboratively planning how care will be delivered; 5) ensuring that service users engaging in group therapies and activities have understood and consented to the ways of working of the group and that face-to-face alternatives are of equivalent quality. |
| CMO 2.2: Removing barriers – greater convenience for users and family/friends | [19, 24, 38, 45, 50, 52, 55, 64, 65, 77, 80, 85, 86, 89, 91, 92, 94, 100-102, 114, 116, 124-131] | Among some service users and family and other supporters experiencing specific practical barriers to attending face-to-face services (childcare or other caring responsibilities, location, work, mobility limitations travel difficulties/costs, work commitments) and who have good access to telemental health (C), telemental health may provide increased flexibility that addresses | 1) Parents with young children, people with caring responsibilities, people who struggle to travel due to work commitments/disability/costs; 2) children and young people in school or higher education (so they can access mental health care without having to leave their place of education); 3) people who live in remote areas or a long distance away from a specialist service; 4) people for whom travel is challenging due to impaired mobility or sensory impairments, or mental health | 1) Offering explicit choice wherever possible between telemental health and face-to-face care, including home visits where services are able to provide this, also taking into account that different modalities may be used for different purposes; 2) identification of people for whom attendance at office appointments is challenging so that telemental health (or home visits) can be considered; 3) continuing to offer choice and checking preferences throughout the duration of care (i.e. do not just ask once); 4) avoiding missed appointments by offering a |
|  |  | individual practical barriers (M), which can lead to telemental health being viewed by some service users and carers as more convenient and accessible than face-to-face care (O1), easing attendance (O2), increasing uptake and reducing missed appointments (O3). | difficulties such as agoraphobia. 5) there may be more advantage for treatments which involve the support of family and friends. | switch to telemental health as an option when a service user is unable at short notice to attend a face-to-face appointment. |
| CMO 2.3: Involvement and support for family and friends | [89, 101, 132-134] | When family and other supporters are invited (with service user agreement) to join telemental health sessions (C), this may result in more holistic treatment planning and greater engagement of family and others in supporting service users (O1), may help improve therapeutic relationships and treatment success (O2), increase engagement (O3), reduce some uncertainty and anxiety around treatment (O4), and may increase satisfaction of and support for family and friends (O5), as family and other supporters may be able to participate in care planning meetings and assessments that they would have found it difficult to attend face-to-face, increasing their engagement in supporting service users and their understanding of their difficulties and care plans (M). | This is especially helpful for 1) those living in locations different from their family and friends or where family and friends have caring or work commitments preventing them from attending meetings face-to-face 2) children and young people (as this may allow their parents to be more involved in their care); 3) service users in inpatient settings where family and friends cannot visit (for example, because of epidemic-related restrictions) or a hospital is in a remote location. | 1) Working with service users to identify any family and friends whose attendance at care planning and other clinical meetings (including on inpatient wards) would be helpful, including those for whom telemental health would facilitate access, such as people in distant locations or whose commitments would make it difficult to attend face-to-face meetings; 2) using strategies as for service users to provide guidance on using telemental health to family and friends and to prepare them for appointments; 3) offering children and their families the opportunity to have telemental health appointments (or, if feasible, home visits) if they find it easier to participate as a family without having to travel to an appointment and to be seen in a clinical setting; 4) on inpatient wards, providing charged iPads, short cables or charging lockers to allow service users to charge their own devices so that they can use technology to connect with family or other supporters. |
| CMO 2.4: Widening the range of available mental health services and treatments | [59, 135-137] | For service users who may benefit from services that they cannot readily access locally and that provide specialised forms of treatment and support regionally or nationally (C), telemental health can | People to whom this is relevant may include: 1) people who have complex clinical needs or rarer conditions such that they would potentially benefit from assessment, treatment and support from specialist services provided at regional and | 1) Development (including of funding arrangements) and dissemination of information about specialist services accessible via telemental health; 2) access for service users, their family and friends and clinicians to information and signposting regarding |
| via telemental health |  | widen the range of specialist assessment, treatment and support available (M), which potentially leads to improved access to services tailored to individual needs and culturally appropriate or specialist services (O1), and to improved satisfaction and treatment outcomes (O2), although an impoverished range of local face-to-face provision may be a risk if referral to distant specialist care via telemental health becomes routine (O3). | national rather than local levels; 2) people who may be able to access distant therapists who speak their own language or interpreters of rare languages not available locally; 3) people who would benefit from support from voluntary organisations that meet specific needs not catered for locally (for example, that support particular cultural groups, LGBTQ+ groups, or people with sensory impairments); 4) people who would benefit from a wider choice of therapies and support (including peer support) than is available locally. | community and voluntary sector organisations beyond their catchment area that are accessible via telemental health; 3) development of safeguards against erosion of local and/or in-person national specialist services in favour of routine specialist telemental health, in line with public sector equality duty to anticipate and provide for the needs of groups with protected characteristics under the Equality Act (for example, pregnant people who are at increased risk of domestic violence, people whose disabilities cause sensory hypersensitivity and struggle with screentime, etc.). |
| CMO 2.5: Adaptations for those with sensory or psychological barriers to telemental health | [50, 60, 87, 138] | Offering face-to-face (or telephone) appointments to people who struggle to cope with sensory (visual or auditory) aspects of telemental health or have symptoms that are exacerbated by it (C), may help to improve engagement with mental health care (O) as the adverse effects of the switch to telemental health for these symptoms and sensory or cognitive impairments may be avoided and service users are able to access their preferred modality of care (M). | This may be relevant for people with 1) symptoms that may interfere with or be exacerbated by engaging with telemental health, such as persecutory ideas or hearing voices, 2) autism; 3) sensory or cognitive impairments; and 4) migraines | 1) Ensuring that face-to-face appointments (including home visits if there are impediments to office appointments) remain available; 2) making clinicians aware of the kinds of clients who may find it particularly difficult to engage with telemental health; 3) adapting telemental health where helpful, for example, through switching off cameras, using telephone rather than video calls, or communicating via text message. |
| CMO 2.6: Inclusion of multidisciplinary and interagency teams in meetings | [99, 134] | When mental health consultations are conducted using telemental health (C), this enables inclusion of staff in appointments who are based geographically far away or who have schedules that would not have allowed them to join a face-to-face session (O1), meaning care and support has potential to be more holistic and integrated (O2), as it is possible for staff from different services and sectors to provide perspectives and contribute to plans (M). | Key contexts include: 1) Hospital inpatients, where telemental health may enable staff who work with them in community settings to join reviews and ward rounds (especially in pandemic conditions where they cannot attend in person); 2) people with complex treatment and support, who are receiving support from more than one team or sector. | 1) Working with service users to identify staff whom it would be helpful to involve in consultations such as review and care planning meetings, including in social care, housing and the voluntary sector; 2) facilitating the involvement of such staff in reviews via telemental health, especially where face-to-face attendance is not feasible. |
| CMO 2.7:<br>Continuing to offer face-to-face care | [63, 91, 102, 135, 139] | When service providers offer care of equivalent quality and timeliness face-to-face (including home visits where needed) rather than via telemental health to service users who do not wish or do not feel able to receive their care remotely (C), this ensures that care can continue and that inequalities in provision are not created or exacerbated (O), because it provides a choice to service users and avoids the negative impacts of digital exclusion(M). | Contexts where face-to-face options may be preferable, and choice is especially important include people who: 1) do not have access to a private space; 2) live with people they do not wish to be overheard by in their appointments (including perpetrators of domestic abuse); 3) do not feel comfortable communicating via remote means; 4) do not want therapy to intrude on their private lives. Also, some service users who value the time spent travelling to and from face-to-face appointments to process emotions may find face-to-face options particularly useful. | 1) Ensuring services are able to offer a choice between telemental health and equivalent care delivered face-to-face (especially when telemental health is part of routine care rather than a means of managing a national emergency); 2) ensuring (as in CMO 2.1) that clinicians are fully aware of service user preferences and circumstances (which may be elicited via a shared decision making tool) and continue to monitor these over time 3) that clinicians are alert for any changing circumstances during telemental health where a service user does not feel comfortable to speak, and make alternative arrangements accordingly (e.g. using text functions on videoconferencing platforms or arranging face-to-face appointments). |
| CMO 2.8:<br>Communication between staff | [19, 95, 140] | When remote technology platforms are used to facilitate real-time communication between staff members, including managers or clinicians working in different teams (C), this can lead to improved efficiency, more convenient working and staff management (O1), improved communication and collaborative planning (O2), and process improvement opportunities (O3), as staff have the ability to rapidly share information, keep track of evolving telemental health procedures (for example, during emergencies) and make collaborative decisions (M). | Contexts where this is relevant include: 1) multidisciplinary teams who are not working on the same site; 2) complex provider organisations with management teams and clinicians working on multiple sites; 3) situations in which people may be receiving care from multiple teams, for example, from an inpatient or crisis service as well as a continuing care service. | 1) Making use of telemental health platforms to strengthen liaison and collaboration between teams and professionals on different sites, for example, through increased enhanced liaison between managers across an organisation, or provide better access to a range of educational events; 2) using telemental health platforms to facilitate multidisciplinary team meetings between staff on different sites (especially if some are working from home). 3) However, awareness is needed that perceived pressure for staff to provide an immediate response may also negatively affect their work-life balance. |

Our findings emphasise the importance of taking individual service user preferences into account, when deciding whether to make use of telemental health, in selecting the modality of digital communication used (including the type of technology platform), and in decisions about involving others (clinicians or family members) in care. This finding underpins all other CMOs in this theme and coheres with theories regarding the importance of shared decision making, collaborative care planning and personalisation in mental health care [103–105]. Involving service users and carers in decisions and care planning as part of a collaborative approach to mental health care has been identified as central to best practice [106]: for example, a review of collaborative care for depression and anxiety found this approach to be more effective than usual care in improving treatment outcomes [107].

Flexible use of telemental health was also identified as being beneficial in reducing barriers to accessing mental health support for some service users, particularly those who may struggle to access face-to-face services, for reasons including caring or work commitments, problems travelling (for example, due to a physical disability, anxiety, or lack of transport), or a reluctance to attend the stigmatising places. Telemental health can also facilitate connections between clinicians, especially across different services or specialties, which can improve multi-disciplinary working and collaboration across teams and agencies, and give service users to a wider range of specialists or support for specific groups. This approach has been identified as having salience in a mental health setting [108–110]. In some cases, telemental health was viewed by both service users and clinicians as more convenient, as it reduced the need for (and cost of) travelling to face-to-face appointments.

Telemental health was also seen as an important tool on inpatient wards, especially during the COVID-19 pandemic when visiting was restricted, as it allowed service users to stay in touch with family and friends, and for them to be involved in their care. It also allowed staff supporting inpatients in the community to remain involved.

However, instances were identified where telemental health is not appropriate and face-to-face care needs to be available. For example, some service users do not wish or feel able to receive care by remote means, or do not wish to have all appointments by this means, while others may struggle with telemental health due to sensory or psychological factors, or a lack of access to appropriate technology and internet connectivity.

### Domain 3. Safety, Privacy and Confidentiality

Table 4 presents the four overarching CMOs, key contexts, and example strategies and solutions for the domain safety, privacy and confidentiality. Key messages were the importance of ensuring the availability of a private space for both service users and clinicians (3.1), the potential for telemental health to provide privacy to some service users experiencing stigma (3.2), the importance of considering how to manage risk when using telemental health, and the limits to how far this is possible (3.3), and data security and staff training (3.4).

**Table 4.** Domain 3. Safety, Privacy and Confidentiality.

| CMO title | References | Overarching CMO | Key Contexts | Example strategies and solutions |
| --- | --- | --- | --- | --- |
| CMO 3.1: Lack of privacy | [49, 52, 55, 58, 64, 65, 68, 70, 87, 89-92, 100, 101, 116, 120, 121, 123, 130, 139, 141-146] | When accessing telemental health sessions without access to a private space or secure private connection (C), service users and staff are at an increased risk of being overheard (M1), potentially leading to breaches of privacy and confidentiality (O1), risk of harm to those in unsafe domestic situations (O2) and reluctance to speak openly about sensitive topics (O3). It may also cause some service users to experience frustration, distress and anxiety (M2) leading to impacts on service user engagement and interactions (O4) and reduced willingness to use telemental health and continue therapy (O5). | Issues related to lack of privacy at home are especially relevant for: 1) young people who are distracted by their home environment, may not feel safe in their own home, or have siblings/parents/other family members unexpectedly appearing in the room; 2) parents with children at home; 3) those experiencing domestic abuse who are not able to be honest about symptoms, risk and/or violence experienced; 4) people who may be living with/caring for extended family, or in households which are crowded; 5) inpatients who may not have a space where they feel *psychologically safe*; 6) people living in houses of multiple occupation; 7) staff members who are not able to work in a private environment when providing remote therapy; 8) service users who experience cultural stigma in the home from their families relating to their mental health. | 1) Brainstorming with the service user whether there are potential options for private places or times when privacy is more likely; 2) offering face-to-face sessions when a private space is not available for telemental health, especially if there is any possibility that the person is at risk from someone in their home environment; 3) regular, discreet checking that the service user (and therapist) is in a private space (for example using the chat function in video calls) and taking steps to provide alternative locations if not; 4) being flexible regarding the time of appointments; 5) allowing people to turn off their camera or use virtual or blurred backgrounds (as well as ensuring that the option is available and they are aware how to do this); 6) working with schools to provide safe spaces away from home for children (although young people may not want to alert teachers/other pupils to their need for a space to use for therapy). 7) attention to clinicians' access to a private space, and disclosure to service users if they are not in a completely private environment, for example a shared office or a private home with other family members on the premises 8) use of headsets with microphones. |
| CMO 3.2: Privacy, anonymity, and reduced stigma | [50, 53, 102, 122, 124, 147-150] | For some service users who feel stigmatised when attending a mental health service in person and who have access to a private and secure space to receive therapy remotely (C), being provided with the option of telemental health as an alternative means there is an option to receive care with more anonymity (M), which helps ensure their privacy and safety (O1), thereby increasing the accessibility of services (O2). | Some groups may be more likely to feel there is a stigma associated with attending mental health premises or reluctant to have contact with others doing so, for example, young people not previously in contact with services. | Offering telemental health (or home visits) to avoid missed appointments to people who are reluctant to attend mental health premises because of perceived stigma or because they find them intimidating. |
| CMO 3.3:<br>Managing risk | [24, 53, 95, 101, 130, 131, 138, 144, 151-155] | When services incorporate tailored risk management procedures in delivery of remote care (C), this encourages consideration of the risks associated with remote care specific to each individual, including risk of self-harm or suicide as well as risk from others in situations of domestic abuse, and ensures staff are aware of the procedures to try to assess and respond to risk or safeguarding concerns despite challenges associated with remote care (M), which has the potential to improve the safety and wellbeing of service users and others (O1). However, a disadvantage of telemental health is that real-time risk assessment limits an immediate response to be organised when someone is at imminent risk of harm and some distance away (O2). | 1) People who are currently unwell or in a crisis; 2) situations where someone is remote from the assessing clinician or at a location unknown to them; 3) situations where technological difficulties occur during an assessment of someone who is at high risk; 4) people with eating disorders or who are physically unwell, where there are practical impediments to assessing risk remotely; 5) when a service user suddenly exits during a telemental health consultation and it is not clear why; 6) in substance misuse services it may be harder to detect if someone is under the influence of drugs or alcohol. | 1) Establishing a call-back number before commencement of the session in the case of disconnection when discussing distressing or sensitive topics; 2) increased coordination with service users/families to facilitate safe transport to emergency departments if needed; 3) setting clear protocols regarding when staff can be contacted via digital means, including who to contact instead in the case of emergency; 4) identification of where the service user is located at the start of the session to enable a faster response of in-person support if needed; 5) development of a "telehealth manual" containing information on what to do in the event of sudden ending of the call and who to contact; 6) co-development of a crisis plan with the service user; 7) offering 24/7 helplines and continued availability of face-to-face crisis response, including capacity for home visits; 8) ensuring adequate device battery or connecting to charger at the start of the session to reduce the risk of disconnection. |
| CMO 3.4:<br>Technological support and information security | [95] | When services provide technology support, software with appropriate security and devices (including mobile phones and headphones) to staff specifically for work use (C), this helps ensure privacy and confidentiality for both service users and staff (O), | 1) In services where staff share office space and devices, or where shortage of devices may lead to use of personal devices, for example for home working; 2) when balancing service user preference with risk from using less secure software, or software with which staff are less | 1) Providing data safeguarding and other technology-based training to all staff (as knowledge cannot be assumed); 2) providing information on which software is encrypted/secure; 3) providing funding to staff for the purchase of equipment; 4) setting recommended boundaries for both service |
|  |  | because staff can store information securely on devices that are not shared with others (M1) and are able to ensure that service users are aware of when they will have access to their work devices (M2). | familiar. 3) where software has a particular set of settings which must be enabled to ensure secure, private connections. | users and clinicians in relation to privacy of personal life and maintaining a work-life balance, for example, by not being contacted outside working hours or using a personal phone. |

With the most supporting literature in this domain, CMO 3.1 highlights the need for appropriate private space to receive telemental health, and that many service users may not have consistent access to such a space. As a lack of privacy can risk breaches in confidentiality and safety for some, a key message was that alternatives such as face-to-face, or alternative times/locations to receive telemental health should be provided. The importance of privacy for effective mental health care has been frequently cited in the literature and is likely to be especially important in ensuring high quality telemental health [101]. Although some literature indicated that some service users feel an increased sense of privacy and a reduction in stigma when not having to attend mental health clinics in person (3.2), a key message lies in providing choice so that each individual can work with their clinicians to find ways of receiving care that they are happy with, a message highlighted in CMOs throughout this paper.

CMOs in this domain also make it clear that telemental health can result in greater risks, both directly because it may be more difficult for clinicians to assess and respond to risks (3.3) and indirectly if data security is impaired (3.4). In both cases, proactive steps to assess and limit risk prior to use of telemental health, as well as pre-planned strategies to respond to events that threaten safety, are important. Data security knowledge should not be assumed and training to help staff keep service users’ personal information secure will also mitigate telemental health-specific risks. However, evidence from both the literature and the stakeholder consultations made it clear that it is difficult to fully overcome the obstacles to effective risk assessment and management that result from staff and service users being in different places, meaning that the continuing availability of an in-person community crisis response is also important.

### Domain 4. Therapeutic quality and relationships

Table 5 displays the overarching CMOs, key contexts, strategies and solutions for the final domain, therapeutic quality and relationships. Therapeutic relationships have been identified as pivotal for the successful delivery of telemental health across the literature and stakeholder consultations. The domain addresses barriers (4.1, 4.2) to and facilitators (4.3, 4.4a-c, 4.5, 4.6) of the development of therapeutic relationships and delivery of quality care and discusses the impact of telemental health on staff wellbeing (4.7).

**Table 5.** Domain 4. Therapeutic quality and relationship.

| CMO title | References | Overarching CMO | Key Contexts | Example strategies and solutions |
| --- | --- | --- | --- | --- |
| CMO 4.1: Change in non-verbal cues and informal chat impacting the therapeutic relationship | [14, 19, 24, 38, 41, 50, 55, 59, 64, 65, 70, 77, 86, 87, 89, 91, 100-102, 113, 115, 122, 164-169] | When switching from face-to-face to telemental health care (C), staff and some service users perceived the relationship between staff and service users (and/or other service user group members) to be negatively affected or found it more difficult to develop a therapeutic relationship (O1) and were thus less willing to take up or use telemental health (O2), more likely to be dissatisfied (O3), and viewed care as less effective compared to previously received face-to-face care (O4). This was because they perceived telemental health to be impersonal and found it more difficult to discuss personal information due to a lack of non-verbal feedback, eye contact, and social cues as well as informal chat before, after, and during sessions (M). | 1) During rapid switches to telemental health due to an emergency situation such as COVID-19 in which staff training and structured telemental health implementation is limited due to time constraints; 2) staff with limited training and experience generally, and those with limited experience of using telemental health specifically may lack the confidence to navigate the change in visual cues which in turn can impact the therapeutic relationship; 3) staff and service users who are new to a specific service, staff/service user, or to mental health care generally; 4) services users who are apprehensive of technology use or who are concerned about the violation of their privacy; 5) group telemental health sessions in which the flow of conversations is impacted, or people may find it less easy to establish relationships and be at ease with the whole group. | 1) Offering new service users the option to receive their first appointment face-to-face when starting telemental health depending on their preference; 2) under pandemic conditions, exploring whether service users prefer telemental health sessions over face-to-face sessions which require wearing masks; 3) checking in with service users about their experiences and preferences regularly while trying to use a particular telemental health platform consistently; 4) allocating additional time to address service user concerns about technology use and privacy; 5) using high quality equipment and ensuring good camera placement during video calls; 6) making greater efforts to communicate clearly, enhance gestures, and to provide verbal and non-verbal reinforcement, such as active listening and backchanneling, i.e. non-verbal or verbal responses; 7) focusing on service user-centred communication, such as being reassuring and supportive; 8) taking more time to informally chat and get to know new service users 1:1 when delivering the initial appointment via telemental health; 9) providing training to staff to increase their comfort with technology and training to interpret social cues when using telemental health; 10) providing reassurance to staff that service users often perceive the therapeutic relationship to be less affected by telemental health than staff believe; 11) facilitating relationships between service user group members by keeping the video call open after the main session to allow follow-up conversations. |
| CMO 4.2:<br>Assessment via<br>telemental health<br>vs face-to-face | [50, 92,<br>102, 130,<br>140, 152,<br>164, 165,<br>170-172] | When using telemental health for assessments (C), staff report finding it more difficult to assess mental health problems, care needs and/or risk, and make diagnoses (O), as they are less able to observe non-verbal and visual cues (depending on the telemental health modality used) and some service users may find it more difficult to have in-depth conversations about their problems and experiences (M). | Cues can include extrapyramidal symptoms from antipsychotics, hygiene, gait, direct eye contact, mannerism, and linguistic nuances.<br>Conducting assessments might be particularly difficult 1) over the phone due to the lack of visual cues; 2) with service users who experience domestic violence and abuse (DVA) and thus cannot be honest about their wellbeing and current situation in the presence of their abuser; 3) with young children; 4) with service users who find it difficult to speak directly about their difficulties and experiences; 5) if staff make incorrect assumptions about service users' mental states based on behavioural indicators and without considering service user reports, especially of neurodivergent service users. | 1) Offering service users the option to receive care face-to-face for first assessments and in crisis situations; 2) taking both service user reports and non-verbal and visual cues into account for assessments; 3) offering service users experiencing DVA the option to use text-based communication in addition to face-to-face care or other telemental health modalities to avoid being overheard; 4) providing training to staff in conducting assessments using telemental health. |
| CMO 4.3: Staff<br>support and<br>training | [59, 73,<br>121, 173] | When staff receive specific instructions as well as training, for example, on how to build rapport using telemental health and support from colleagues with prior telemental health experience (C), this facilitates quality of care (O1), building therapeutic relationships (O2), and increased engagement (O3), as staff are able to ask questions and acquire new skills and knowledge about the interventions and thus build confidence in delivering telemental health (M). | This is likely to be especially relevant to staff who have little or no previous experience of delivering telemental health. | Offering staff training on aspects of good care that go beyond technical skills and issues. Staff training should ideally be co-designed and co-delivered with service users. |
| CMO 4.4a:<br>Service users finding it easier to establish a therapeutic relationship online | [49, 70, 89, 115, 118, 138, 155, 164, 174, 175] | When delivering telemental health to some services users who feel uncomfortable in clinical settings and social situations (C), these service users find it easier to build a therapeutic relationship and are more willing to use telemental health (O), as they feel safer, are more relaxed and less anxious being in their own environment and/or outside of clinical settings and in-person social situations and thus feel more empowered and comfortable to open up and speak freely (M). | This may be especially relevant for: 1) some children and young people, including those with special needs and neurodivergent children, who find clinical settings and having to travel upsetting; 2) some service users with social anxiety. However, it is important that using telemental health does not reinforce potentially detrimental safety behaviours that may maintain and potentially exacerbate their social anxiety. | Offering service users the option of receiving care by telemental health rather than face-to-face, especially if they neither wish to attend clinical settings nor to be visited by professionals at home |
| CMO 4.4b:<br>Service users finding it easier to establish a therapeutic relationship via video vs phone | [24, 38, 60, 62, 65, 88, 113, 124] | When service users and staff who prefer video calls use them (instead of telephone calls or text-based chats) for telemental health (C), this can facilitate a stronger therapeutic relationship (O1), satisfaction (O2) and engagement (O3), as it is easier to see visual and non-verbal cues, gauge the therapist's reaction, and connect with the service user/staff member compared to other telemental health modalities (M). | This applies to service users across age groups and may be especially the case for new service users. | Encouraging clinicians to offer video calls rather than relying on phone calls and text messaging and providing the relevant infrastructure and guidance to support this. |
| CMO 4.4c:<br>Service users finding it easier to establish a therapeutic relationship via the phone vs face-to-face or video-calls | [60, 75, 90, 130] | When services offer phone calls and text messages instead of video calls (C), some service users are more satisfied with their care (O), as they do not have to sit still and see themselves on screen, are less conscious of their body language and facial gestures, are less distracted by the clinician's non-verbal cues, are able to move around freely, and are thus less inhibited and able to open up more quickly (M). | This might be especially relevant for service users who are: 1) neurodivergent; 2) socially anxious; 3) self-conscious about their appearance. | 1) Informing service users about the option to turn off their camera during video calls or using the phone if they are uncomfortable; 2) Offering a telephone call or text service instead if the service user prefers this. |
| CMO 4.5: More frequent telemental health | [49, 50, 118, 123, 176] | When services adapt flexibly to service users' preferences regarding the pattern and frequency of telemental health | 1) Lack of a need to travel means that more frequent shorter sessions may be particularly feasible with | Considering offering shorter and more frequent sessions when telemental health is a primary modality for delivering care. |
| sessions plus text messages |  | sessions, including offering more frequent, shorter rather than infrequent, long sessions, and/or additional asynchronous text messages and calls to check in between sessions (C), this may lead to stronger therapeutic relationships (O1), increased engagement (O2), and improved quality of care (O3), as service users receive regular and more frequent support depending on their preference (M). | telemental health: they are potentially less tiring and thus might better maintain concentration and engagement, especially for children; 2) frequent sessions might help new service users to build trust and reduce anxiety around the treatment; 3) frequent sessions may also help support and monitor less stable service users, for example, following a crisis. |  |
| CMO 4.6: Enhancing quality of care through use of telemental health enhancements | [24, 84, 120, 177-179] | When clinicians make appropriate and personalised use of enhancements and extensions of telemental health (such as using chat, voice activation to instruct phones, SMS and other text-based messaging, online appointment schedules, screen sharing and apps accessed during sessions) (C), this can lead to success engaging in telemental health (O1) and broadening the range of strategies and interventions available during clinical meetings (O2), as these features made engaging with services easier, and provided a functional method useful for exchanging practical information, such as reminding service users about the date and purpose of an appointment, with less room for ambiguity and more creative methods of engagement (M). | Additional telemental health features might be particularly helpful for: 1) young children (who overall find it difficult to engage online); 2) adolescents who experience social anxiety and/or are autistic may benefit from and prefer the chat function. | 1) Using text messaging (including apps) to maintain communication in a flexible way between appointments, especially for younger people for whom this may be a preferred method of communication; 2) using screen sharing to facilitate psychoeducation or working together on assessment or therapeutic tools; 3) using telemental health sessions to introduce apps and websites that support self-management or therapy, or to collaboratively complete measures and questionnaires. |
| CMO 4.7: Staff wellbeing and quality of care | [61-63, 90, 98, 101, 120, 126, 131, 180] | When staff utilise the time saved on travel to take breaks in between telemental health sessions (C), this may increase staff wellbeing (O1) and improve quality of care (O2), as it provides the opportunity to reflect and recharge after telemental health | 1) Clinicians who can work wholly or partly at home; 2) teams working across different sites or who visit service users at home or in other community settings. | 1) Supporting clinicians to plan their time so that they can work from home and save travel time on some days; 2) Considering appointing some interested professionals to fully remote roles in which they can develop skills and make efficient use of time; 3) Ensuring that |
|  |  | sessions, which are often experienced as tiring, and thus reduces fatigue, tension, and anxiety among staff (M1), and staff can use some of the time on clinical work, catch up on administrative tasks, or engage in professional developmental activities (M2). |  | when time is saved because travel is not needed, clinicians still have suitable breaks between on-screen appointments and are able to dedicate some of the time saved to their own professional development; 4) Fostering a working culture in which staff are encouraged to take breaks between telemental health sessions in order to reflect and recharge. |

Trust and therapeutic relationships are important across health care, and relational aspects of care are especially crucial in mental health [156–161]. However, the reliance on telemental health platforms, particularly telephone and text-based communication, may affect communication and subsequently therapeutic relationships (4.1). Our CMOs, particularly their mechanisms, are informed by general theories regarding the role and development of therapeutic relationships in mental health care.

CMO 4.1. highlights that telemental health is likely to lead to a change or reduction in visual and non-verbal cues, including active listening and back channels, facial expressions, gestures, posture, and eye contact, which makes aspects of communication, such as pauses, difficult to interpret. Additionally, time delays in video calls may create silences, and lead to talking over each other and delayed visual responses which negatively impact communication and non-verbal synchrony [162, 163]. As a result, not only therapeutic relationships but also staff’s ability to conduct accurate assessments are compromised (4.1, 4.2). Those making first contact with mental health services appear to be particularly impacted by the potentially impersonal nature of telemental health and thus benefit not only from an initial face-to-face session but also more frequent subsequent telemental health sessions to establish stability and trust (4.1, 4.5). Additionally, our findings indicate that staff confidence and ability to deliver good quality care and develop therapeutic relationships via telemental health can be fostered through training sessions provided by services (4.3).

The literature and stakeholder consultations identified no telemental health modality that is consistently superior for developing therapeutic relationships (4.4a-c). Rather, whether telephone calls, video calls, or face-to-face meetings are most appropriate seems to depend on the purpose of sessions and on an individual’s preferences, based on their personal experiences and circumstances, whether they are new to the service, as well as the nature of their mental or physical health problems. Video-calls seem to be often preferred for more substantial and in-depth sessions compared to other telemental health modalities [24]. Providing service users with choice regarding the frequency, duration, and telemental health modality is crucial for therapeutic relationships and quality of care.

Despite its limitations, flexible use of different telemental health modalities can provide significant opportunities to foster therapeutic relationships and increase quality of care, such as checking in and sending reminders via text messages and using features such as chat functions to increase engagement among service users (4.6).

Lastly, taking breaks in between telemental health sessions and fostering positive telemental health working environments is key for staff wellbeing and the delivery of high-quality care (4.7).

## Discussion

### Key findings

Our RRR identified CMOs within four key domains, each with a range of practical implications regarding what works for whom in telemental health: connecting effectively; flexibility and personalisation; safety, privacy and confidentiality; and therapeutic quality and relationship. Potentially the most important finding of this realist review is the significance of personal choice, and that one size does not fit all for telemental health. This includes choice of modality (for example, video, telephone, text-based chat functions), platform, frequency or duration of sessions, and the option to revert to face-to-face sessions if preferred or required by the service user based on their current context, or to vary modality from contact to contact. This review has highlighted that there are many contexts where face-to-face care is preferred or needed by service users, and this should be accessible and available to them, and should be of equivalent timeliness to remote care (especially when delivered as part of routine care rather than as a response to a national emergency). However, use of telemental health is a convenient and potentially advantageous option for some people in many contexts, so it is beneficial for mental health clinicians to have the skills and resources to offer telemental health as an option. When service users’ choice about what works for them is respected and decisions about care planning are made collaboratively, this is likely to be conducive to a stronger therapeutic relationship where the service user feels heard and respected [161].

Access to a device with stable internet connection, and the confidence and ability to use a device to access telemental health, were identified as minimum requirements for both staff and service users to access telemental health, without which face-to-face appointments would be necessary. The devices and platforms used for delivering telemental health needed to be user-friendly [47, 48], and preferably familiar, to easily facilitate sessions. Telemental health seemed to reduce some barriers to receiving mental health support experienced by some service users, for example, those who were unable to travel or on inpatient wards, making it an acceptable alternative to face-to-face sessions for some people under these circumstances. It may also potentially allow service users greater access to out-of-area specialist services and to support focused on specific groups (for example cultural or LGBTQ+ groups). Issues of privacy, including data protection and confidentiality, or staff and service user access to a private space were emphasised throughout the literature and our consultations; this is likely to disproportionately affect disadvantaged groups of service users, such as those experiencing poverty, in multi-occupancy households, children and young people, or people living with controlling or abusive partners or other family members. Already disadvantaged groups are likewise at particularly high risk of inequalities being exacerbated through digital exclusion. The “inverse digital care law”, that use of digital technologies can make health inequalities worse [35], may well apply to widespread implementation of telemental health [17]. Service planning and delivery must be based on a strong awareness of these risks and the need to overcome these barriers. There is also a duty to ensure in-person care of equivalent quality remains readily available.

The impact on therapeutic relationships, for both staff and service users, has also been highlighted, with difficulties interpreting visual or non-verbal cues cited as a barrier to establishing a good therapeutic relationship that enables service users to disclose sensitive information and staff being able to conduct valid clinical assessments. Adapting to service user preferences flexibly and giving weight to self-reports during assessments is likely to increase quality of care and foster strong therapeutic relationships.

### Strengths and limitations

The use of RRR methodology to rapidly establish a set of theories about what works for whom in which circumstances in telemental health has several strengths. The breadth of written evidence screened extends beyond published academic literature to non-academic (including policy, third sector and lived experience) sources. The targeted call for evidence, sent directly to expert stakeholders from research, policy and clinical settings (nationally and internationally), the voluntary sector, lived experience groups, minority groups, representatives from health tech initiatives, identified resources that would otherwise have been missed. Through these procedures, we rapidly identified literature from a wide range of key perspectives to contribute to the development of the CMOs.

The analysis process was rigorous and valued both published literature and stakeholder views, with the use of rapid realist methods allowing a range of stakeholder perspectives to be incorporated beyond what is normally possible in reviews. Our expert reference group (including clinical, academic, and lived experience experts) fed into the review process and theory development throughout, iteratively reviewing the plausibility, relevance, and usefulness of our individual and overarching CMOs. A wider group of expert stakeholders provided further input to identifying sources and reviewing overarching CMOs, especially regarding our priority groups: children and young people, users of inpatient and crisis care services and digitally excluded populations. Continuous detailed feedback from the lived experience researchers and frontline clinicians helped to reduce bias towards academic perspectives, ensured the inclusion of a breadth of real-life experiences and supported the iterative development of our methods, results and interpretation of findings.

A final key benefit of the RRR methodology is that we could rapidly investigate not only outcomes of telemental health use, but the mechanisms underlying what works for whom, which most methodologies do not allow. We could also explore the contexts in which telemental health was implemented, as well as the telemental health resources that are available. This approach should be considered for future evaluation of telemental health.

Some limitations should be noted. The first relates to generalisations made in the process of developing overarching CMOs. These tended to combine underlying CMOs that related to a range of service user and clinician groups, service settings and types, and social and national contexts. We looked for important themes that appeared of general relevance and were validated through stakeholder consultation. However, it is likely that in some areas we have lost a more nuanced understanding of the relationship between particular CMOs and particular contexts. We also utilised a broad definition of telemental health to capture as much richness (and data) as possible from the available sources. However, we have merged heterogenous forms of telemental health within most of our overarching CMOs, which may differ in their effectiveness and underlying mechanisms. Therefore, conclusions regarding mechanisms, outcomes for specific types of telemental health and the impact on service users, staff and carers are limited. We also included literature that draws on experiences of service users and clinicians both pre- and during the pandemic. However, technologies and approaches to implementation have changed substantially, with pre-pandemic evidence tending to focus on planned and relatively small-scale implementation of tools specifically designed for mental health. Studies from the pandemic tend to relate to a range of phone and video call technologies implemented at scale with limited strategic planning. During the pandemic, staff and service users may also have been more willing to trial telemental health given the extraordinary circumstances. The available technologies, and clinicians’ and service users’ skills in applying them, are also likely to have changed over time, and to continue to change.

The nature and strength of evidence drawn on for the review also needs to be noted. Most sources were qualitative studies, service evaluations or cross-sectional studies of associations; we found few relevant trials or longitudinal studies. We have tried to maximise the value of this body of evidence by combining findings from multiple studies with expert stakeholder input to obtain theories with multiple sources of support about what works for whom, illustrating them with example contexts and strategies. However, lack of testing through traditionally robust methods in testing intervention strategies, such as trials and other longitudinal forms of evaluation, still needs to be noted, as discussed further in the implications for research below.

Despite the inclusive search strategy and specific efforts to gain a wide range of perspectives, digitally excluded groups remain underrepresented in this study. This is partly due to lack of literature focused on digital exclusion and to the online methods used to conduct our review during the COVID-19 pandemic. Efforts were made to gain perspectives on digitally excluded groups by including charities, and staff and service user advocates working with people in such groups, including in projects aimed at addressing digital exclusion, in our stakeholder consultation. However, people experiencing severe digital exclusion obviously did not participate in our online consultations, and the extent to which others can advocate for them is limited. Similarly, this study identified a lack of evidence in the literature about how to make telemental health engaging and effective for young children, nor were we able to find many people with relevant expertise to participate in our consultations. Data was also limited on group therapy and the role or experiences of families and other supporters of service users. Most available literature focused exclusively on staff perspectives of telemental health, and crucially neglected to include the views or experiences of service users and their families or other supporters. Therefore, we were unable to incorporate these groups and their perspectives in our analysis and synthesis.

This study was initially planned and commissioned through discussions between policymakers in the Department of Health and Social Care and the MHPRU leads; lived experience researchers did not have the opportunity to contribute during the early stages of formulating research questions and identifying the methodology to be used.

### Implications

#### Implications for clinical practice

A range of implications for clinical practice and service planning can be drawn from our CMOs. The challenge for the future will be to find sustainable ways to implement them in clinical practice and to find an appropriate balance between telemental health and traditional face-to-face care in future service delivery. In the context of a recent emergency (the COVID-19 pandemic), telemental health has been used with some degree of success to maintain care for at least some service users. Evidence and experiences from this widespread emergency implementation are helpful, both to inform future response to such emergencies, and to allow a preliminary assessment of potential opportunities and pitfalls in implementing telemental health beyond an emergency context.

Some clear principles to guide practice emerge from our CMOs. Offering choice, planning care collaboratively and listening to personal preference regarding whether to use telemental health need to be embedded within services in which there is continuing use of telemental health as we move through and out of the COVID-19 pandemic. How choice is negotiated, enabled and communicated is crucial. For choice to be real, options need to be clearly explained and discussed at every stage, face-to-face care of equal quality should be delivered as promptly as telemental health, and choice should be seen as dynamic, especially when a service user is in crisis. Preferences should be reconfirmed regularly, and hybrid forms of care made available if appropriate. Choices may also be different following the COVID-19 pandemic, when risk of infection travelling to and at appointments may no longer be a concern and consultations are no longer masked; mask-wearing at most face-to-face appointments during the pandemic may undermine some advantages of in-person care. Ideally, service user and clinician choice and resources should be balanced through shared decision making. Use of telemental health cannot be assumed to be a permanent switch, so preferences should be revisited regularly. In planning services, it may be easier to switch from in-person appointments to digital than vice-versa, and this needs to be considered in staffing and working space arrangements. Traditional inpatient and community services are limited in their ability to collaborate and provide choice in their established processes, such as care planning and risk assessment or management [160, 161, 181–183]. It may, therefore, be unrealistic to expect improvements in these areas when delivering telemental health.

Lack of access to digital devices or data, or of expertise in connecting to telemental health services, is a problem that service providers may be able to address for some people. For example, opportunities to develop skills and clear guidance and opportunities to practice may be relatively straightforward ways to alleviate problems with connecting effectively for some people who may find telemental health a convenient way to receive some care if they are supported to engage. At best, getting access to telemental health may be a skill acquired along with developing the skills to access a variety of other significant parts of the digital world. In other instances, clinicians and service providers should be aware that digital exclusion tends to be rooted in other forms of disadvantage, and that they can most readily avoid exacerbating such disadvantage by offering face-to-face care. Persevering with telemental health when service users do not want to receive care by this means and are not in the habit of using digital technologies may prove futile in every day clinical care. There is also likely to be scope for improving the extent to which service providers have the capacity to connect effectively, for example, through better infrastructure, clear guidance and training for staff, and clarity and flexibility regarding platforms.

Developing a therapeutic relationship is key for the quality and success of care, and offering initial appointments as face-to-face may facilitate this, subject to service user choice. Additionally, services and staff may need to consider how to adapt telemental health care to account for the change in visual cues, including body language and facial expressions (although visual cues in face-to-face sessions may in any case be compromised while infection control considerations mean most sessions are masked).

The privacy, safety and confidentiality domain also has implications for clinical practice. Clinician awareness of potential risks associated with using telemental health is important and may steer them away from conducting some consultations in this way. Maintaining privacy and safety, for example for people at risk within their homes, is also a significant reason to prioritise service user choice, especially choices not to accept telemental health appointments, as they may not readily be able to explain the basis for their choice. Clinicians need also to be aware of the challenges of assessing and responding to risk when using telemental health, including the advantages of face-to-face meetings, the need to give weight to service user reports where visual or verbal cues may be obscured, and the importance of back-up plans, such as for disconnection or when an urgent response is needed. Clinicians and care coordinators could also helpfully ensure that service users have access to and can use telemental health care adequately before the agreed online sessions begin, although this may be affected by staffing issues and limited resources [184, 185].

#### Implications for policy

Digital poverty does not exist in isolation, and the experience of poverty may be the root cause of their digital exclusion. Providing service users with access to devices and internet, for example, serves as an adequate short-term fix but does not address the systemic welfare issues experienced by many service users [186]. Strategies to mitigate digital exclusion could include the provision of good national Wi-Fi coverage, free broadband [187], and investment in accessible, connected community hubs; implementing these would require action from the government, rather than health services.

Telemental health services seem to be a viable alternative to face-to-face care for some service users, including in emergency situations, such as COVID-19. In order to provide good telemental health services, investment is needed, for example, in providing telemental health specific training and guidance, high quality infrastructure and potentially technological devices to staff and service users. Pre-registration education and training should include skills in telemental health. Further investment is likely to be needed in updating this as evidence and technologies change (for example, to cover the ongoing costs of keeping hardware and software up to date). This may need to be balanced against any savings anticipated from implementing telemental health. This study has also highlighted the importance of service user and frontline staff involvement in the planning of all telemental health services and provision.

#### Implications for research

Much of the research included was based on explorations of views and experiences of people participating in telemental health in various settings. We found few studies involving systematic evaluation of planned strategies to achieve high quality implementation of telemental health in routine settings. Primary studies of this form would be valuable, potentially using implementation research and participatory action research models to explore outcomes and experiences of strategies aimed at good quality implementation of telemental health in varying real-world settings during and post-pandemic. Our CMOs have potential to inform such a primary research study: it would be helpful to develop and test co-produced strategies for implementing principles encapsulated in the CMOs in real-world settings. Similarly, our understanding of what works for whom in telemental health would be improved by conducting primary research with specific groups, particularly those excluded from previous studies, such as digitally excluded groups or peer support networks, and in specific contexts, such as in group therapy sessions. Identifying methods of reaching digitally excluded populations in research studies, as well as of identifying groups for whom telemental health is not appropriate, would be helpful. This is likely to be labour and time intensive and needs appropriate funding. Future research could also usefully explore the use of different telemental health modalities individually and in more depth.

Choice has been emphasised as crucial in the use of telemental health. The mechanisms behind choice and collaborative decision making would benefit from further investigation, potentially using realist methods and drawing on principles from shared decision-making research. Investigation is warranted of the best approaches to providing the information needed to make an informed choice, holding collaborative discussions about how to personalise care for each individual, and providing staff and service users with guidance and training needed to participate effectively in telemental health. At a provider level, evidence is needed about what makes a good telemental health platform, how to balance data security with the flexibility that service users and clinicians may value in choosing platforms and using familiar tools if possible, and how to adapt risk management to a telemental health context. However, Trusts may compromise their ability to offer choice and flexibility to service users when they specify the platforms that can and cannot be used to deliver telemental health services (although this may have advantages, including increasing staff familiarity).

Researchers conducting evaluations of telemental health should consider that ‘satisfaction’ tends to be evaluated as one component. However, satisfaction with telemental health consists of several components which need to be individually considered. For example, the skills of therapists may be rated highly, while telemental health platforms themselves may cause significant frustration and if they were scored separately would be poorly rated. Telemental health therefore needs to be evaluated as several elements rather than as a singularity.

Impacts of telemental health delivery on staff is a further key area of investigation. Some staff in research studies and in our stakeholder consultations reported finding prolonged screen use draining and perceived it as a contributor to burnout. The impact of telemental health on staff and ways of ensuring that it does not increase burnout, or physical or psychological stress requires investigation.

It may be pertinent to investigate whether services function better, and staff and service users are more satisfied, when certain staff become telemental health specialists, as opposed to asking all to engage with it for some appointments.

A final key consideration is that any future research into telemental health should include lived experience knowledge, expertise and views, including those from digitally excluded groups; there is a need to fund research designed and led by people with lived experience of mental health service use.

## Lived Experience Commentary

Written by Rachel Rowan Olive, Karen Machin and Prisha Shah

We welcome the question “what works for whom, in what circumstances, and how?”. At its heart, a realist review understands that each person has different needs from services, including telemental health services. The challenge is the reliance on existing knowledge, and the potential to overlook gaps, especially where the world has changed rapidly because of COVID-19.

The digital methods used to consult a wider audience also further marginalise everyone who does not have, or want, such access. Including people who do not use telemental health would produce different research questions and answers. Similarly, including technology experts might provide some reassurance, for example, about regulation, risk and ethics raised by practices such as the recent sale of data from a US crisis text line to a for-profit artificial intelligence company [188].

Digital technology has increased restrictive practice in mental health via surveillance [189], sometimes based on poor quality research conducted with financial involvement from manufacturers [190]. Health data has been shared with police in programmes such as Serenity Integrated Monitoring (SIM) on shaky legal and ethical grounds [191]. While these are not telemental health per se, they provide a context. In that context, we would have liked the question “What works for whom?” to consider political and financial interests.

This study’s methods encouraged a discussion of choice, personalisation, and flexibility, which we welcome. We highlight two reflections.

Firstly, choice is not only about preferring one option over another: it can be life-or-death. Within mental health, service users are often expected to bare our souls to get our choices respected. With telemental health, this is dangerous. If the criteria for accessing a face-to-face service are harm-based, we might be forced to put ourselves at risk to get what we need. Where someone is being abused by their partner, they may need face-to-face services, but not explain why at a first assessment. We must be taken at our word without explaining ourselves to clinicians who have not yet earned our trust.

Secondly, choice is limited by the available options, which are constrained by material circumstances and power. Service users generally have relatively little power in their relationships with an overstretched system. If a wheelchair user’s choice is to travel to a building with an unreliable lift, versus telemental health - that is not a meaningful choice. If you have to wait six months for a face-to-face appointment, but you can have telemental health next week - that is not a meaningful choice. If you cannot afford to connect to the internet, you do not have a meaningful choice. The option of telemental health must not become an excuse to allow face-to-face services to become harder to access.

Many of the actions for telemental health implementation are specific applications of general principles of good care: informed consent to make meaningful choices; clarity about our health data breeding trust; understanding and responding to the contexts in which we live our lives.

Within such contexts, we welcome a focus on digital poverty as poverty. The policy solutions to poverty lie well beyond mental health: a broader overhaul of the punitive welfare system and a society in which workers are empowered to negotiate liveable wages.

## Data Availability

All data produced in the present work are contained in the manuscript and appendices.

## Notes

### Competing Interest Statement

The authors have declared no competing interest.

### Funding Statement

This paper presents independent research commissioned and funded by the National Institute for Health Research (NIHR) Policy Research Programme, conducted by the NIHR Policy Research Unit (PRU) in Mental Health (grant no. PR-PRU-0916-22003). The views expressed are those of the authors and not necessarily those of the NIHR, the Department of Health and Social Care or its arm's 591 length bodies, or other government departments. The funders had no role in study design, data collection and analysis, decision to publish, or preparation of the manuscript.

## Reference List

1. Barnett P, Goulding L, Casetta C, Jordan H, Sheridan-Rains L, Steare T, et al. Implementation of Telemental Health Services Before COVID-19: Rapid umbrella review of systematic reviews. Journal of Medical Internet Research. 2021;23(7):e26492.

2. Appleton R, Williams J, San Juan NV, Needle JJ, Schlief M, Jordan H, et al. Implementation, Adoption, and Perceptions of Telemental Health During the COVID-19 Pandemic: Systematic Review. Journal of Medical Internet Research. 2021;23(12):e31746.

3. Agency for Healthcare Research and Quality. Telehealth. [23 September 2020]; Available from: https://digital.ahrq.gov/key-topics/telehealth.

4. Hensel J, Graham R, Isaak C, Ahmed N, Sareen J, Bolton J. A Novel Emergency Telepsychiatry Program in a Canadian Urban Setting: Identifying and Addressing Perceived Barriers for Successful Implementation. The Canadian Journal of Psychiatry. 2020;65(8):559–67.

5. Bashshur RL, Shannon GW, Bashshur N, Yellowlees PM. The empirical evidence for telemedicine interventions in mental disorders. Telemedicine and e-Health. 2016;22(2):87–113.

6. Salmoiraghi A, Hussain S. A systematic review of the use of telepsychiatry in acute settings. Journal of Psychiatric Practice. 2015;21(5):389–93.

7. Telebehavioural Health Institute. Telehealth bibliography. [23 September 2020]; Available from: https://telehealth.org/bibliography/.

8. Sheridan Rains L, Johnson S, Barnett P, Steare T, Needle JJ, Carr S, et al. Early impacts of the COVID-19 pandemic on mental health care and on people with mental health conditions: Framework synthesis of international experiences and responses. Social Psychiatry and Psychiatric Epidemiology. 2021;56(1):13–24.

9. Seritan AL, Heiry M, Iosif A-M, Dodge M, Ostrem JL. Telepsychiatry for patients with movement disorders: A feasibility and patient satisfaction study. Journal of Clinical Movement Disorders. 2019;6(1):1–8.

10. APA Work Group. Telepsychiatry Toolkit. [23 September 2020]; Available from: https://www.psychiatry.org/psychiatrists/practice/telepsychiatry/toolkit.

11. Hilty DM, Ferrer DC, Parish MB, Johnston B, Callahan EJ, Yellowlees PM. The effectiveness of telemental health: A 2013 review. Telemedicine and e-Health. 2013;19(6):444–54.

12. Varker T, Brand RM, Ward J, Terhaag S, Phelps A. Efficacy of synchronous telepsychology interventions for people with anxiety, depression, posttraumatic stress disorder, and adjustment disorder: A rapid evidence assessment. Psychological Services. 2019;16(4):621–35.

13. Gloff NE, LeNoue SR, Novins DK, Myers K. Telemental health for children and adolescents. International Review of Psychiatry. 2015;27(6):513–24.

14. Hubley S, Lynch SB, Schneck C, Thomas M, Shore J. Review of key telepsychiatry outcomes. World Journal of Psychiatry. 2016;6(2):269–82.

15. Fletcher TL, Hogan JB, Keegan F, Davis ML, Wassef M, Day S, et al. Recent advances in delivering mental health treatment via video to home. Current Psychiatry Reports. 2018;20(8):1–9.

16. Santesteban-Echarri O, Piskulic D, Nyman RK, Addington J. Telehealth interventions for schizophrenia-spectrum disorders and clinical high-risk for psychosis individuals: A scoping review. Journal of Telemedicine and Telecare. 2020;26(1-2):14–20.

17. Davies AR, Honeyman M, Gann B. Addressing the digital inverse care law in the time of COVID-19: Potential for digital technology to exacerbate or mitigate health inequalities. Journal of Medical Internet Research. 2021;23(4):e21726.

18. Spanakis P, Peckham E, Mathers A, Shiers D, Gilbody S. The digital divide: Amplifying health inequalities for people with severe mental illness in the time of COVID-19. The British Journal of Psychiatry. 2021;219(4):529–31.

19. Johnson S, Dalton-Locke C, San Juan NV, Foye U, Oram S, Papamichail A, et al. Impact on mental health care and on mental health service users of the COVID-19 pandemic: A mixed methods survey of UK mental health care staff. Social Psychiatry and Psychiatric Epidemiology. 2021;56(1):25–37.

20. Hutchings R. The impact of Covid-19 on the use of digital technology in the NHS. Nuffield Trust. 2020 (23 Sep 2020):1–23.

21. Watkins S. Twitter2020 [cited 21 January 2022]; Available from: https://twitter.com/SteveWatkinsNHS/status/1304467986556891136.

22. Fisk M, Livingstone A, Pit SW. Telehealth in the context of COVID-19: Changing perspectives in Australia, the United Kingdom, and the United States. Journal of Medical Internet Research. 2020;22(6):e19264.

23. Whaibeh E, Mahmoud H, Naal H. Telemental health in the context of a pandemic: The COVID-19 experience. Current Treatment Options in Psychiatry. 2020;7(2):198–202.

24. Vera San Juan N, Shah P, Schlief M, Appleton R, Nyikavaranda P, Birken M, et al. Service user experiences and views regarding telemental health during the COVID-19 pandemic: A co-produced framework analysis. PloS one. 2021;16(9):e0257270.

25. Madigan S, Racine N, Cooke JE, Korczak DJ. COVID-19 and telemental health: Benefits, challenges, and future directions. Canadian Psychology/Psychologie Canadienne. 2021;62(1):5.

26. Saul JE, Willis CD, Bitz J, Best A. A time-responsive tool for informing policy making: Rapid realist review. Implementation Science. 2013;8(1):1–15.

27. Pawson R, Tilley N, Tilley N. Realistic evaluation: Sage; 1997. ISBN: 0761950095.

28. Lemire S, Kwako A, Nielsen SB, Christie CA, Donaldson SI, Leeuw FL. What is this thing called a mechanism? Findings from a review of realist evaluations. New Directions for Evaluation. 2020;2020(167):73–86.

29. Khangura S, Konnyu K, Cushman R, Grimshaw J, Moher D. Evidence summaries: The evolution of a rapid review approach. Systematic Reviews. 2012;1(1):1–9.

30. Stolee P, Elliott J, McNeil H, Boscart V, Heckman GA, Hutchinson R, et al. Choosing Healthcare Options by Involving Canada’s Elderly: A protocol for the CHOICE realist synthesis project on engaging older persons in healthcare decision-making. BMJ Open. 2015;5(11):e008190.

31. Willis C, Saul J, Bitz J, Pompu K, Best A, Jackson B. Improving organizational capacity to address health literacy in public health: A rapid realist review. Public Health. 2014;128(6):515–24.

32. Gillard S, Dare C, Hardy J, Nyikavaranda P, Rowan Olive R, Shah P, et al. Experiences of living with mental health problems during the COVID-19 pandemic in the UK: A coproduced, participatory qualitative interview study. Social Psychiatry and Psychiatric Epidemiology. 2021;56(8):1447–57.

33. Page MJ, McKenzie JE, Bossuyt PM, Boutron I, Hoffmann TC, Mulrow CD, et al. The PRISMA 2020 statement: An updated guideline for reporting systematic reviews. International Journal of Surgery. 2021;88:105906.

34. Thomas J, Brunton J, Graziosi S. EPPI-Reviewer 4.0: Software for research synthesis. 2010.

35. Watts G. COVID-19 and the digital divide in the UK. The Lancet Digital Health. 2020;2(8):e395–e6.

36. Kim KJ, Sundar SS. Mobile persuasion: Can screen size and presentation mode make a difference to trust? Human Communication Research. 2016;42(1):45–70.

37. Kortum P, Johnson M, editors. The relationship between levels of user experience with a product and perceived system usability. Proceedings of the Human Factors and Ergonomics Society Annual Meeting; 2013: SAGE Publications Sage CA: Los Angeles, CA.

38. Severe J, Tang R, Horbatch F, Onishchenko R, Naini V, Blazek MC. Factors influencing patients’ initial decisions regarding telepsychiatry participation during the COVID-19 pandemic: Telephone-based survey. JMIR Formative Research. 2020;4(12):e25469.

39. Shore JH, Yellowlees P, Caudill R, Johnston B, Turvey C, Mishkind M, et al. Best practices in videoconferencing-based telemental health April 2018. Telemedicine and e-Health. 2018;24(11):827–32.

40. Nelson E-L, Bui T, Sharp S. Telemental health competencies: Training examples from a youth depression telemedicine clinic. Technology Innovations for Behavioral Education: Springer; 2011. p. 41–7.

41. Lopez A, Schwenk S, Schneck CD, Griffin RJ, Mishkind MC. Technology-based mental health treatment and the impact on the therapeutic alliance. Current Psychiatry Reports. 2019;21(8):1–7.

42. MacIntyre G, Cogan NA, Stewart AE, Quinn N, Rowe M, O’Connell M. What’s citizenship got to do with mental health? Rationale for inclusion of citizenship as part of a mental health strategy. Journal of Public Mental Health. 2019.

43. Eiroa-Orosa FJ, Rowe M. Taking the concept of citizenship in mental health across countries. Reflections on transferring principles and practice to different sociocultural contexts. Frontiers in Psychology. 2017;8:1020.

44. Cogan NA, MacIntyre G, Stewart A, Tofts A, Quinn N, Johnston G, et al. “The biggest barrier is to inclusion itself”: The experience of citizenship for adults with mental health problems. Journal of Mental Health. 2021;30(3):358–65.

45. Shore P, Goranson A, Ward MF, Lu MW. Meeting veterans where they’re@: a VA Home-Based Telemental Health (HBTMH) pilot program. The International Journal of Psychiatry in Medicine. 2014;48(1):5–17.

46. Greener S, Wakefield C. Developing confidence in the use of digital tools in teaching. Electronic Journal of E-Learning. 2015;13(4):260–7.

47. Harst L, Lantzsch H, Scheibe M. Theories predicting end-user acceptance of telemedicine use: systematic review. Journal of Medical Internet Research. 2019;21(5):e13117.

48. Nielsen J, Landauer TK, editors. A mathematical model of the finding of usability problems. Proceedings of the INTERACT’93 and CHI’93 conference on Human factors in computing systems; 1993.

49. Bierbooms JJ, van Haaren M, IJsselsteijn WA, de Kort YA, Feijt M, Bongersi IM. Integration of online treatment into the “new normal” in mental health care in post–COVID-19 times: Exploratory qualitative study. JMIR Formative Research. 2020;4(10):e21344.

50. Chen JA, Chung W-J, Young SK, Tuttle MC, Collins MB, Darghouth SL, et al. COVID-19 and telepsychiatry: Early outpatient experiences and implications for the future. General Hospital Psychiatry. 2020;66:89–95.

51. Ghaneirad E, Groba S, Bleich S, Szycik GR. Use of outpatient psychotherapy via video consultation. Psychotherapeut. 2021:1–6.

52. Ghosh A, Mahintamani T, Subodh B, Pillai RR, Mattoo S, Basu D. Telemedicine-assisted stepwise approach of service delivery for substance use disorders in India. Asian Journal of Psychiatry. 2021;58:102582.

53. Kanellopoulos D, Castellano CB, McGlynn L, Gerber S, Francois D, Rosenblum L, et al. Implementation of telehealth services for inpatient psychiatric Covid-19 positive patients: A blueprint for adapting the milieu. General Hospital Psychiatry. 2021;68:113–4.

54. Lakeman R, Crighton J. The impact of social distancing on people with borderline personality disorder: The views of dialectical behavioural therapists. Issues in Mental Health Nursing. 2021;42(5):410–6.

55. Mind. Trying to connect: The importance of choice in remote mental health services. 2021.

56. Mental Health Network NHS Confederation. Digital inclusion in mental health. 2020.

57. Simpson J, Doze S, Urness D, Hailey D, Jacobs P. Evaluation of a routine telepsychiatry service. Journal of Telemedicine and Telecare. 2001;7(2):90–8.

58. Watson A, Mellotte H, Hardy A, Peters E, Keen N, Kane F. The digital divide: factors impacting on uptake of remote therapy in a South London psychological therapy service for people with psychosis. Journal of Mental Health. 2021:1–8.

59. Disney L, Mowbray O, Evans D. Telemental health use and refugee mental health providers following COVID-19 pandemic. Clinical Social Work Journal. 2021;49(4):463–70.

60. Costa M, Reis G, Pavlo A, Bellamy C, Ponte K, Davidson L. Tele-mental health utilization among people with mental illness to access care during the CoViD-19 pandemic. Community Mental Health Journal. 2021;57(4):720–6.

61. Bommersbach T, Dube L, Li L. Mental health staff perceptions of improvement opportunities around COVID-19: A mixed-methods analysis. Psychiatric Quarterly. 2021;92(3):1079–92.

62. Buckman JE, Saunders R, Leibowitz J, Minton R. The barriers, benefits and training needs of clinicians delivering psychological therapy via video. Behavioural and Cognitive Psychotherapy. 2021;49(6):696–720.

63. Foye U, Dalton-Locke C, Harju-Seppänen J, Lane R, Beames L, Vera San Juan N, et al. How has COVID-19 affected mental health nurses and the delivery of mental health nursing care in the UK? Results of a mixed-methods study. Journal of Psychiatric and Mental Health Nursing. 2021;28(2):126–37.

64. Frayn M, Fojtu C, Juarascio A. COVID-19 and binge eating: Patient perceptions of eating disorder symptoms, tele-therapy, and treatment implications. Current Psychology. 2021;40(12):6249–58.

65. Hawke LD, Sheikhan NY, MacCon K, Henderson J. Going virtual: Youth attitudes toward and experiences of virtual mental health and substance use services during the COVID-19 pandemic. BMC Health Services Research. 2021;21(1):1–10.

66. He Z, Chen J, Pan K, Yue Y, Cheung T, Yuan Y, et al. The development of the “COVID-19 Psychological Resilience Model” and its efficacy during the COVID-19 pandemic in China. International Journal of Biological Sciences. 2020;16(15):2828.

67. Lecomte T, Abdel-Baki A, Francoeur A, Cloutier B, Leboeuf A, Abadie P, et al. Group therapy via videoconferencing for individuals with early psychosis: A pilot study. Early Intervention in Psychiatry. 2021;15(6):1595–601.

68. Medalia A, Lynch DA, Herlands T. Telehealth conversion of serious mental illness recovery services during the COVID-19 crisis. Psychiatric Services. 2020;71(8):872.

69. The Mental Health Innovation Network. Inclusive messaging: Supporting refugee mental health in the United States during COVID-19. 2020 [31 January 2022]; Available from: https://www.mhinnovation.net/blog/2020/apr/19/inclusive-messaging-supporting-refugee-mental-health-united-states-during-covid-19

70. Ogueji IA, Amusa AO, Olofe OJ, Omotoso EB. Willingness and barriers to utilizing e-therapy services: A Nigerian general population qualitative study. Journal of Human Behavior in the Social Environment. 2021;32(2):214–28.

71. Arighi A, Fumagalli GG, Carandini T, Pietroboni AM, De Riz MA, Galimberti D, et al. Facing the digital divide into a dementia clinic during COVID-19 pandemic: Caregiver age matters. Neurological Sciences. 2021;42(4):1247–51.

72. Castillo M, Conte B, Hinkes S, Mathew M, Na C, Norindr A, et al. Implementation of a medical student-run telemedicine program for medications for opioid use disorder during the COVID-19 pandemic. Harm Reduction Journal. 2020;17(1):1–6.

73. Conn DK, Madan R, Lam J, Patterson T, Skirten S. Program evaluation of a telepsychiatry service for older adults connecting a university-affiliated geriatric center to a rural psychogeriatric outreach service in Northwest Ontario, Canada. International Psychogeriatrics. 2013;25(11):1795–800.

74. Godleski L, Cervone D, Vogel D, Rooney M. Home telemental health implementation and outcomes using electronic messaging. Journal of Telemedicine and Telecare. 2012;18(1):17–9.

75. Greenwood J, Chamberlain C, Parker G. Evaluation of a rural telepsychiatry service. Australasian Psychiatry. 2004;12(3):268–72.

76. Humer E, Stippl P, Pieh C, Schimböck W, Probst T. Psychotherapy via the Internet: What programs do psychotherapists use, how well-informed do they feel, and what are their wishes for continuous education? International Journal of Environmental Research and Public Health. 2020;17(21):8182.

77. Juarez-Reyes M, Mui HZ, Kling SM, Brown-Johnson C. Accessing behavioral health care during COVID: rapid transition from in-person to teleconferencing medical group visits. Therapeutic Advances in Chronic Disease. 2021;12:1–11.

78. Lin X, Swift J, Cheng Y, An Q, Liang H, Wang Y, et al. The psychological hotline services quality survey during the pandemic of COVID-19 in mainland China. International Journal of Mental Health Promotion. 2020;22(3):109–13.

79. Mental Health Commission of Canada. E-Mental health in Canada: Transforming the mental health system using technology. Ottawa, ON: Mental Health Commission of Canada; 2014.

80. Open Excellence. Hearing Voices USA groups go virtual in response to COVID-19 2020 [31 January 2022]; Available from: https://openexcellence.org/hearing-voices-usa-groups-go-virtual-in-response-to-covid-19/.

81. Pote H, Rees A, Holloway-Biddle C, Griffith E. Workforce challenges in digital health implementation: How are clinical psychology training programmes developing digital competences? Digital Health. 2021;7:1–11.

82. Rabinowitz T, Murphy KM, Amour JL, Ricci MA, Caputo MP, Newhouse PA. Benefits of a telepsychiatry consultation service for rural nursing home residents. Telemedicine and e-Health. 2010;16(1):34–40.

83. Scharff A, Breiner CE, Ueno LF, Underwood SB, Merritt EC, Welch LM, et al. Shifting a training clinic to teletherapy during the COVID-19 pandemic: A trainee perspective. Counselling Psychology Quarterly. 2021;34(3-4):676–86.

84. Williams H, Whelan A, Team R. An investigation into access to digital inclusion for healthcare for the homeless population. Seaview; 2017.

85. Yellowlees P, Nakagawa K, Pakyurek M, Hanson A, Elder J, Kales HC. Rapid conversion of an outpatient psychiatric clinic to a 100% virtual telepsychiatry clinic in response to COVID-19. Psychiatric Services. 2020;71(7):749–52.

86. Fogler JM, Normand S, O’Dea N, Mautone JA, Featherston M, Power TJ, et al. Implementing group parent training in telepsychology: Lessons learned during the COVID-19 pandemic. Journal of Pediatric Psychology. 2020;45(9):983–9.

87. Healthwatch. Ready set connect: Exploring young people’s perceived barriers to accessing video/phone mental health appointments - a solution-focused consultation. 2021.

88. Lichstein KL, Scogin F, Thomas SJ, DiNapoli EA, Dillon HR, McFadden A. Telehealth cognitive behavior therapy for co-occurring insomnia and depression symptoms in older adults. Journal of Clinical Psychology. 2013;69(10):1056–65.

89. Lodder A, Papadopoulos C, Randhawa G. Using a blended format (videoconference and face to face) to deliver a group psychosocial intervention to parents of autistic children. Internet Interventions. 2020;21:100336.

90. McBeath AG, Du Plock S, Bager-Charleson S. The challenges and experiences of psychotherapists working remotely during the coronavirus* pandemic. Counselling and Psychotherapy Research. 2020;20(3):394–405.

91. Pugh M, Bell T, Dixon A. Delivering tele-chairwork: A qualitative survey of expert therapists. Psychotherapy Research. 2021;31(7):843–58.

92. Uscher-Pines L, Sousa J, Raja P, Mehrotra A, Barnett M, Huskamp HA. Treatment of opioid use disorder during COVID-19: Experiences of clinicians transitioning to telemedicine. Journal of Substance Abuse Treatment. 2020;118:108124.

93. Aafjes-van Doorn K, Békés V, Prout TA. Grappling with our therapeutic relationship and professional self-doubt during COVID-19: Will we use video therapy again? Counselling Psychology Quarterly. 2021;34(3-4):473–84.

94. Newbronner E, Spanakis P, Wadman R, Crosland S, Heron P, Johnston G, et al. Business as Un-usual: Access to mental health and primary care services for people with severe mental illness during the COVID-19 restrictions. Frontiers in Psychiatry. 2021;12:799885.

95. O’Dell SM, Hosterman SJ, Parikh MR, Winnick JB, Meadows TJ. Chasing the curve: Program description of the Geisinger primary care behavioral health virtual first response to COVID-19. Journal of Rural Mental Health. 2021;45(2):95–106.

96. Barney A, Buckelew S, Mesheriakova V, Raymond-Flesch M. The COVID-19 pandemic and rapid implementation of adolescent and young adult telemedicine: Challenges and opportunities for innovation. Journal of Adolescent Health. 2020;67(2):164–71.

97. Connolly SL, Stolzmann KL, Heyworth L, Weaver KR, Bauer MS, Miller CJ. Rapid increase in telemental health within the Department of Veterans Affairs during the COVID-19 pandemic. Telemedicine and e-Health. 2021;27(4):454–8.

98. Hopkins L, Pedwell G. The COVID PIVOT - Re-orienting child and youth mental health care in the light of pandemic restrictions. Psychiatric Quarterly. 2021;92(3):1259–70.

99. Peralta EA, Taveras M. Effectiveness of teleconsultation use in access to mental health services during the coronavirus disease 2019 pandemic in the Dominican Republic. Indian Journal of Psychiatry. 2020;62(Suppl 3):S492–S4.

100. Hensel JM, Yang R, Vigod SN, Desveaux L. Videoconferencing at home for psychotherapy in the postpartum period: Identifying drivers of successful engagement and important therapeutic conditions for meaningful use. Counselling and Psychotherapy Research. 2021;21(3):535–44.

101. Liberati E, Richards N, Parker J, Willars J, Scott D, Boydell N, et al. Remote care for mental health: qualitative study with service users, carers and staff during the COVID-19 pandemic. BMJ Open. 2021;11(4):e049210.

102. Sehlo MG, Youssef UM, Elshami MI, Elrafey DS, Elgohari HM. Telepsychiatry versus face to face consultation in COVID-19 Era from the patients’ perspective. Asian Journal of Psychiatry. 2021;59:102641.

103. Alguera-Lara V, Dowsey MM, Ride J, Kinder S, Castle D. Shared decision making in mental health: the importance for current clinical practice. Australasian Psychiatry. 2017;25(6):578–82.

104. Carr S. Personalisation: a rough guide. Social Care Institute for Excellence; 2008.

105. Rodenburg-Vandenbussche S, Carlier I, van Vliet I, van Hemert A, Stiggelbout A, Zitman F. Patients’ and clinicians’ perspectives on shared decision-making regarding treatment decisions for depression, anxiety disorders, and obsessive-compulsive disorder in specialized psychiatric care. Journal of Evaluation in Clinical Practice. 2020;26(2):645–58.

106. Wallcraft J, Amering M, Freidin J, Davar B, Froggatt D, Jafri H, et al. Partnerships for better mental health worldwide: WPA recommendations on best practices in working with service users and family carers. World Psychiatry. 2011;10(3):229–36.

107. Archer J, Bower P, Gilbody S, Lovell K, Richards D, Gask L, et al. Collaborative care for depression and anxiety problems. Cochrane Database of Systematic Reviews. 2012 (10).

108. Vinokur-Kaplan D. Treatment teams that work (and those that don’t): An application of Hackman’s group effectiveness model to interdisciplinary teams in psychiatric hospitals. The Journal of Applied Behavioral Science. 1995;31(3):303–27.

109. Kutash K, Acri M, Pollock M, Armusewicz K, Serene Olin S-c, Hoagwood KE. Quality indicators for multidisciplinary team functioning in community-based children’s mental health services. Administration and Policy in Mental Health and Mental Health Services Research. 2014;41(1):55–68.

110. Miles P. Family partners and the wraparound process. Portland, OR: National Wraparound Initiative, Research and Training Center for Family Support and Children’s Mental Health; 2008. 3 p.

111. Adamou M, Jones SL, Fullen T, Galab N, Abbott K, Yasmeen S. Remote assessment in adults with Autism or ADHD: A service user satisfaction survey. Plos one. 2021;16(3):e0249237.

112. Schueller SM, Hunter JF, Figueroa C, Aguilera A. Use of digital mental health for marginalized and underserved populations. Current Treatment Options in Psychiatry. 2019;6(3):243–55.

113. Eagle. Service user experience of remote therapy during the COVID-19 pandemic and service improvement points 2020.

114. Guinart D, Marcy P, Hauser M, Dwyer M, Kane JM. Patient attitudes toward telepsychiatry during the COVID-19 pandemic: A nationwide, multisite survey. JMIR Mental Health. 2020;7(12):e24761.

115. King R, Bambling M, Lloyd C, Gomurra R, Smith S, Reid W, et al. Online counselling: The motives and experiences of young people who choose the Internet instead of face to face or telephone counselling. Counselling and Psychotherapy Research. 2006;6(3):169–74.

116. Mind. Trying to Connect: The importance of choice in remote mental health services (Young people data cut). 2021.

117. National Health Service England. 24/7 urgent mental health helplines available across the country. [31 January 2022]; Available from: https://www.england.nhs.uk/mental-health/case-studies/24-7-urgent-mental-health-helplines-available-across-the-country/

118. Nissling L, Fahlke C, Lilja JL, Skoglund I, Weineland S. Primary care peer-supported internet-mediated psychological treatment for adults with anxiety disorders: Mixed methods study. JMIR Formative Research. 2020;4(8):e19226.

119. Orlowski S, Lawn S, Matthews B, Venning A, Wyld K, Jones G, et al. The promise and the reality: a mental health workforce perspective on technology-enhanced youth mental health service delivery. BMC Health Services Research. 2016;16(1):1–12.

120. Shklarski L, Abrams A, Bakst E. Navigating changes in the physical and psychological spaces of psychotherapists during Covid-19: When home becomes the office. Practice Innovations. 2021;6(1):55–66.

121. Simon N, Ploszajski M, Lewis C, Smallman K, Roberts NP, Kitchiner NJ, et al. Internet-based psychological therapies: A qualitative study of National Health Service commissioners and managers views. Psychology and Psychotherapy: Theory, Research and Practice. 2021;94(11):994–1014.

122. YoungMinds. YoungMinds submission to the COVID Committee’s inquiry into ‘Living Online: The Long Term Impact of Wellbeing’. 2020.

123. The British Psychological Society. Considerations for psychologists working with children and young people using online video platforms. 2020.

124. Choi NG, Hegel MT, Marti CN, Marinucci ML, Sirrianni L, Bruce ML. Telehealth problem-solving therapy for depressed low-income homebound older adults. The American Journal of Geriatric Psychiatry. 2014;22(3):263–71.

125. Gaddy S, Gallardo R, McCluskey S, Moore L, Peuser A, Rotert R, et al. COVID-19 and music therapists’ employment, service delivery, perceived stress, and hope: A descriptive study. Music Therapy Perspectives. 2020;38(2):157–66.

126. The Health Foundation. CWTCH: Connecting With Telehealth to Children in Hospital. 2020.

127. Hernandez-Tejada MA, Zoller JS, Ruggiero KJ, Kazley AS, Acierno R. Early treatment withdrawal from evidence-based psychotherapy for PTSD: Telemedicine and in-person parameters. The International Journal of Psychiatry in Medicine. 2014;48(1):33–55.

128. Sheehan R, Dalton-Locke C, Ali A, Totsika V, San Juan NV, Hassiotis A. Mental healthcare and service user impact of the COVID-19 pandemic: Results of a UK survey of staff working with people with intellectual disability and developmental disorders. medRxiv. 2020.

129. Simpson J, Doze S, Urness D, Hailey D, Jacobs P. Telepsychiatry as a routine service - the perspective of the patient. Journal of Telemedicine and Telecare. 2001;7(3):155–60.

130. Uscher-Pines L, Sousa J, Raja P, Mehrotra A, Barnett ML, Huskamp HA. Suddenly becoming a “virtual doctor”: Experiences of psychiatrists transitioning to telemedicine during the COVID-19 pandemic. Psychiatric Services. 2020;71(11):1143–50.

131. Wilson CA, Dalton-Locke C, Johnson S, Simpson A, Oram S, Howard LM. Challenges and opportunities of the COVID-19 pandemic for perinatal mental health care: A mixed-methods study of mental health care staff. Archives of Women’s Mental Health. 2021;24(5):749–57.

132. Brooks E, Novins DK, Noe T, Bair B, Dailey N, Lowe J, et al. Reaching rural communities with culturally appropriate care: a model for adapting remote monitoring to American Indian veterans with posttraumatic stress disorder. Telemedicine and e-Health. 2013;19(4):272–7.

133. Moslehi S, Aubrey-Jones D, Knowles M, Obeney-Williams J, Leveson S, Aref-Adib G. Cyberpsychiatry versus COVID-19: Using video consultation to improve clinical care in an in-patient psychiatric unit. BJPsych International. 2021;18(3):1–4.

134. Whistance B, Wright P, Brind J, Johns G, Khalil S, Ahuja A. ’Ask Us About Dementia’ Pilot Support Service Independent Evaluation. Technology Enabled Care (TEC) CYMRU. 2021.

135. Colle R, Ait Tayeb AEK, de Larminat D, Commery L, Boniface B, Lasica PA, et al. Short-term acceptability by patients and psychiatrists of the turn to psychiatric teleconsultation in the context of the COVID-19 pandemic. Psychiatry and Clinical Neurosciences. 2020;74(8):443–4.

136. Crowe T, Jani S, Jani S, Jani N, Jani R. A pilot program in rural telepsychiatry for deaf and hard of hearing populations. Heliyon. 2016;2(3):e00077.

137. Zheng P, Gray MJ. Telehealth-based therapy connecting rural Mandarin-speaking traumatized clients with a Mandarin-speaking therapist. Clinical Case Studies. 2014;13(6):514–27.

138. Holland M, Smith V, Robertson N, Heafield G. Staying connected decision making tools, learning and case studies. Health Innovation Network. 2020.

139. Dores AR, Geraldo A, Carvalho IP, Barbosa F. The use of new digital information and communication technologies in psychological counseling during the COVID-19 pandemic. International Journal of Environmental Research and Public Health. 2020;17(20):7663.

140. Khanna R, Murnane T, Kumar S, Rolfe T, Dimitrieski S, McKeown M, et al. Making working from home work: Reflections on adapting to change. Australasian Psychiatry. 2020;28(5):504–7.

141. Boldrini T, Schiano Lomoriello A, Del Corno F, Lingiardi V, Salcuni S. Psychotherapy during COVID-19: How the clinical practice of Italian psychotherapists changed during the pandemic. Frontiers in Psychology. 2020;11:2716.

142. Mad Covid, Edwards B, Grundy A, Bradford E, Aitch N. COVID-19: Recommendations for Mental Health Services 2020 [18 March 2022]; Available from: http://www.infocop.es/pdf/Recommendations%20for%20Mental%20Health%20Services.pdf.

143. Moore J. Severe mental illness and Covid-19: Service support and digital solutions 2020.

144. Mental Health Network NHS Confederation. Delivering mental gealth services digitally: Briefing 2020.

145. Barratt C. Mental health and houses in multiple occupation: The challenge of turning research into practice. [31 January 2022]; Available from: https://www.emcouncils.gov.uk/write/Mental_Health_and_HMOs_Caroline__Barratt.pdf.

146. De Witte NA, Carlbring P, Etzelmueller A, Nordgreen T, Karekla M, Haddouk L, et al. Online consultations in mental healthcare during the COVID-19 outbreak: An international survey study on professionals’ motivations and perceived barriers. Internet Interventions. 2021;25:100405.

147. National Health Service England. Mental health COVID-19 children and young people case studies. 2021 [31 January 2022]; Available from: https://www.england.nhs.uk/mental-health/case-studies/children-and-young-people-cyp-case-studies/mental-health-covid-19-children-and-young-people-case-studies/.

148. Healthwatch. Remote mental health survey. 2021.

149. James K. Remote mental health interventions for young people: A rapid review of the evidence. Youth Access, 2020.

150. Lannin DG, Vogel DL, Brenner RE, Abraham WT, Heath PJ. Does self-stigma reduce the probability of seeking mental health information? Journal of Counseling Psychology. 2016;63(3):351–8.

151. Benaque A, Gurruchaga MJ, Abdelnour C, Hernández I, Cañabate P, Alegret M, et al. Dementia care in times of COVID-19: Experience at Fundació ACE in Barcelona, Spain. Journal of Alzheimer’s Disease. 2020;76(1):33–40.

152. Martin R, Sturt J, Griffiths F. The impact of digital communication on adolescent to adult mental health service transitions. Journal of Research in Nursing. 2020;25(3):277–88.

153. Pierce BS, Perrin PB, Tyler CM, McKee GB, Watson JD. The COVID-19 telepsychology revolution: A national study of pandemic-based changes in US mental health care delivery. American Psychologist. 2021;76(1):14–25.

154. Rosen CS, Morland LA, Glassman LH, Marx BP, Weaver K, Smith CA, et al. Virtual mental health care in the Veterans Health Administration’s immediate response to coronavirus disease-19. American Psychologist. 2021;76(1):26–38.

155. Smith J, Gillon E. Therapists’ experiences of providing online counselling: A qualitative study. Counselling and Psychotherapy Research. 2021;21(3):545–54.

156. Birkhäuer J, Gaab J, Kossowsky J, Hasler S, Krummenacher P, Werner C, et al. Trust in the health care professional and health outcome: A meta-analysis. PloS one. 2017;12(2):e0170988.

157. Flückiger C, Del Re A, Wampold BE, Symonds D, Horvath AO. How central is the alliance in psychotherapy? A multilevel longitudinal meta-analysis. Journal of Counseling Psychology. 2012;59(1):10–7.

158. Flückiger C, Del Re AC, Wampold BE, Horvath AO. The alliance in adult psychotherapy: A meta-analytic synthesis. Psychotherapy. 2018;55(4):316–40.

159. Kaiser J, Hanschmidt F, Kersting A. The association between therapeutic alliance and outcome in internet-based psychological interventions: A meta-analysis. Computers in Human Behavior. 2021;114:106512.

160. Coffey M, Cohen R, Faulkner A, Hannigan B, Simpson A, Barlow S. Ordinary risks and accepted fictions: How contrasting and competing priorities work in risk assessment and mental health care planning. Health Expectations. 2017;20(3):471–83.

161. Simpson A, Hannigan B, Coffey M, Barlow S, Cohen R, Jones A, et al. Recovery-focused care planning and coordination in England and Wales: A cross-national mixed methods comparative case study. BMC Psychiatry. 2016;16(1):1–18.

162. Paulick J, Deisenhofer A-K, Ramseyer F, Tschacher W, Boyle K, Rubel J, et al. Nonverbal synchrony: A new approach to better understand psychotherapeutic processes and drop-out. Journal of Psychotherapy Integration. 2018;28(3):367–84.

163. Seuren LM, Wherton J, Greenhalgh T, Shaw SE. Whose turn is it anyway? Latency and the organization of turn-taking in video-mediated interaction. Journal of Pragmatics. 2021;172:63–78.

164. Bambling M, King R, Reid W, Wegner K. Online counselling: The experience of counsellors providing synchronous single-session counselling to young people. Counselling and Psychotherapy Research. 2008;8(2):110–6.

165. Grover S, Mehra A, Sahoo S, Avasthi A, Tripathi A, D’Souza A, et al. State of mental health services in various training centers in India during the lockdown and COVID-19 pandemic. Indian Journal of Psychiatry. 2020;62(4):363–9.

166. Olwill C, Mc Nally D, Douglas L. Psychiatrist experience of remote consultations by telephone in an outpatient psychiatric department during the COVID-19 pandemic. Irish Journal of Psychological Medicine. 2021;38(2):132–9.

167. Sasangohar F, Bradshaw MR, Carlson MM, Flack JN, Fowler JC, Freeland D, et al. Adapting an outpatient psychiatric clinic to telehealth during the COVID-19 pandemic: A practice perspective. Journal of Medical Internet Research. 2020;22(10):e22523.

168. LeRouge CM, Garfield MJ, Hevner AR. Patient perspectives of telemedicine quality. Patient Preference and Adherence. 2015;9:25–40.

169. Pinto RZ, Ferreira ML, Oliveira VC, Franco MR, Adams R, Maher CG, et al. Patient-centred communication is associated with positive therapeutic alliance: A systematic review. Journal of Physiotherapy. 2012;58(2):77–87.

170. Wyler H, Liebrenz M, Ajdacic-Gross V, Seifritz E, Young S, Burger P, et al. Treatment provision for adults with ADHD during the COVID-19 pandemic: An exploratory study on patient and therapist experience with on-site sessions using face masks vs. telepsychiatric sessions. BMC Psychiatry. 2021;21(1):1–16.

171. Milton DE. On the ontological status of autism: the ‘double empathy problem’. Disability & Society. 2012;27(6):883–7.

172. Grover S, Mehra A, Sahoo S, Avasthi A, Tripathi A, D’Souza A, et al. Impact of COVID-19 pandemic and lockdown on the state of mental health services in the private sector in India. Indian Journal of Psychiatry. 2020;62(5):488–93.

173. Lindsay JA, Kauth MR, Hudson S, Martin LA, Ramsey DJ, Daily L, et al. Implementation of video telehealth to improve access to evidence-based psychotherapy for posttraumatic stress disorder. Telemedicine and e-Health. 2015;21(6):467–72.

174. McManus F, Sacadura C, Clark DM. Why social anxiety persists: An experimental investigation of the role of safety behaviours as a maintaining factor. Journal of Behavior Therapy and Experimental Psychiatry. 2008;39(2):147–61.

175. Rapee RM, Heimberg RG. A cognitive-behavioral model of anxiety in social phobia. Behaviour Research and Therapy. 1997;35(8):741–56.

176. Chong J, Moreno F. Feasibility and acceptability of clinic-based telepsychiatry for low-income Hispanic primary care patients. Telemedicine and e-Health. 2012;18(4):297–304.

177. The British Psychological Society. Working therapeutically with parents and their infants during pregnancy and postpartum using remote delivery platforms 2020.

178. Jones DJ, Forehand R, Cuellar J, Parent J, Honeycutt A, Khavjou O, et al. Technology-enhanced program for child disruptive behavior disorders: Development and pilot randomized control trial. Journal of Clinical Child & Adolescent Psychology. 2014;43(1):88–101.

179. Riches S, Azevedo L, Steer N, Nicholson S, Vasile R, Lyles S, et al. Brief videoconference-based dialectical behaviour therapy skills training for COVID-19-related stress in acute and crisis psychiatric staff. Clinical Psychology Forum. 2021:57–62.

180. Mahmoud H, Naal H, Cerda S. Planning and implementing telepsychiatry in a community mental health setting: A case study report. Community Mental Health Journal. 2021;57(1):35–41.

181. Coffey M, Hannigan B, Barlow S, Cartwright M, Cohen R, Faulkner A, et al. Recovery-focused mental health care planning and co-ordination in acute inpatient mental health settings: A cross national comparative mixed methods study. BMC Psychiatry. 2019;19(1):1–18.

182. Ahmed N, Barlow S, Reynolds L, Drey N, Begum F, Tuudah E, et al. Mental health professionals’ perceived barriers and enablers to shared decision-making in risk assessment and risk management: A qualitative systematic review. BMC Psychiatry. 2021;21(1):1–28.

183. Zelle H, Kemp K, Bonnie RJ. Advance directives in mental health care: evidence, challenges and promise. World Psychiatry. 2015;14(3):278–80.

184. The British Medical Association. Measuring progress: Commitments to support and expand the mental health workforce in England. 2019.

185. Gilburt H. Securing money to improve mental health care… but no staff to spend it on The King’s Fund; 2019 [31 January 2022]; Available from: https://www.kingsfund.org.uk/blog/2019/10/mental-health-staff-shortage.

186. The Faculty of Public Health. The impact of the UK recession and welfare reform on mental health. [31 January 2022]; Available from: https://www.fph.org.uk/policy-advocacy/special-interest-groups/special-interest-groups-list/public-mental-health-special-interest-group/better-mental-health-for-all/the-economic-case/the-impact-of-the-uk-recession-and-welfare-reform-on-mental-health/.

187. Corbyn J. Full text of Jeremy Corbyn’s speech on Labour’s British Broadband announcement. 2019 [31 January 2022]; Available from: https://labour.org.uk/press/full-text-of-jeremy-corbyns-speech-on-labours-british-broadband-announcement/.

188. Das S. Mental health helpline funded by royals shared users’ conversations. The Guardian. 2022.

189. National Survivor User Network. Surveillance in mental health settings - NSUN response to JCHR inquiry. 2021 [18 March 2022]; Available from: https://www.nsun.org.uk/news/surveillance-in-mental-health-settings-jchr/.

190. Wilson K, Eaton J, Foye U, Ellis M, Thomas E, Simpson A. What evidence supports the use of Body Worn Cameras in mental health inpatient wards? A systematic review and narrative synthesis of the effects of Body Worn Cameras in public sector services. International Journal of Mental Health Nursing. 2022;31(2):260–77.

191. STOPSIM. Concerns regarding privacy and data protection within the High Intensity Network (HIN) and Serenity Integrated Mentoring (SIM). 2021 [18 March 2022]; Available from: https://stopsim.co.uk/2021/04/26/concerns-regarding-privacy-and-data-protection-within-the-high-intensity-network-hin-and-serenity-integrated-mentoring-sim/.

