## Supplemental file 1 for "What works for whom with telemental health? A rapid realist review"

### Appendix 1. Study Characteristics

| Author, year and country | Study aim | Type of report and study design | Type of service | Nature of mental health problem/diagnosis | Participants (n) | Telemental health modalities |
|---|---|---|---|---|---|---|
| Aafjes-van Doorn et al. (2020) USA | Exploring therapists' experiences of switching to video therapy during the pandemic. | Quantitative cross-sectional study | Psychology/psychotherapy/counselling service | Not applicable (NA) | Counsellors and Psychologists (n=141) | Video call |
| Adamou et al. (2021) UK | Investigating if tele-communication methods are preferable to in-person consultations at an Autism and ADHD service during the pandemic. | Quantitative cross-sectional and qualitative survey | Community mental health teams and outpatient services | ADHD and/or autism | Service users (n=117) | Video call, Phone call, Other (combination) |
| Arighi et al. (2021) Italy | Describing the digital divide of patients with dementia contacted by telemedicine during the COVID-19 pandemic. | Quantitative cross-sectional study | General hospital/physical health service | Dementia | Service users (n=108) | Video call |
| Bambling et al. (2008) Australia | Exploring the experience of online counsellors, particularly of text-based communication. | Qualitative focus group study | Helplines | General population | Counsellor (n=26) | Video call, Text message/SMS/Whats App |
| Barney et al. (2020) USA | Reporting on a clinic's rapid transition to audiovisual telemedicine in response to COVID-19. | Service evaluation/audit: Quantitative cross-sectional study | Community mental health teams and outpatient services | Medical conditions, attention and mood disorders, sexual and reproductive health care, eating disorder care, addiction treatment | Mixed groups of mental health professionals (n not stated/unclear) | Video call |
| Benaque et al. (2020) Spain | Describing the experiences of a service in dealing with the COVID-19 pandemic. | Service evaluation/audit | Voluntary sector/non-profit organisation | | NA | Video call, Phone call, Text |

| | | | | | | |
|---|---|---|---|---|---|---|
| | | | | | | message/SMS/Whats App |
| Bierbooms et al. (2020) The Netherlands | Discussing the expectations regarding the sustainability of online treatment post-COVID-19 regulations. | Qualitative interview study | Community mental health teams and outpatient services | Light to complex psychological and psychiatric problems | Mental healthcare staff (n=9), support staff (n=2) | Video call, online modules, Text message/SMS/Whats App |
| Boldrini et al. (2020) Italy | Investigating factors influencing the rate of interrupted treatments during lockdown and psychotherapists' satisfaction with telepsychotherapy. | Quantitative cross-sectional study and qualitative survey | Community mental health teams and outpatient services; Psychology/psychotherapy/counselling service; Private hospital/clinic | Mixed primary care or psychology/psychotherapy service users | Psychotherapist or analyst (n=308) | Video call, Phone call, Mixed F2F and telemental health |
| Bommersbach et al. (2021) USA | Identifying staff perceptions on changes that should be maintained or improved in services. | Quantitative cross-sectional study and qualitative focus group study | General hospital/physical health service | NA | Staff (n=99 in survey; n=25 in focus group) | Video call, online patient portals |
| Brooks et al. (2013) USA | Describing the cultural adaption model. | Service evaluation/audit | Community mental health teams and outpatient services | PTSD | Service users (n not stated) | Video call |
| Buckman et al. (2021) UK | Exploring 1) changes of psychological therapy during the pandemic; 2) clinicians' experience and confidence of delivering video-based psychological therapy; and 3) barriers, benefits, and training needs in respect to video-based psychological therapy delivery. | Quantitative cross-sectional study | Community mental health teams and outpatient services | NA | Therapists (n=66) | Video call |
| Castillo et al. (2020) USA | Describing the development of TeleMOUD tool (medications for Opiod Use disorder) to mitigate disruptions in care for people with Opiod Use Disorder. | Quantitative cross-sectional study; descriptive study; Implementation study | Community mental health teams and outpatient services | Opioid dependency and drug injection | Service users and individuals who inject drugs (n=22) | Video call, Phone call |

| | | | | | | |
|---|---|---|---|---|---|---|
| Chen et al. (2020) USA | Reviewing payment and regulatory changes in telepsychiatry in March and April 2020. | Commentary/editorial (with data) | General hospital/physical health service. | Mixed secondary mental health service users | NA (description of service change) | Video call, Phone call |
| Choi et al. (2014) USA | Evaluating the acceptance and preliminary efficacy of in-home telehealth delivery of problem-solving therapy (tele-PST) among low-income homebound older adults | Quantitative randomised controlled trial (RCT) | Aging service agencies | Depression | Service users (n=121) | Video call, Phone call |
| Chong and Moreno (2012) USA | Comparing telepsychiatry services through the Internet in primary care settings. | Quantitative randomised controlled trial (RCT) | Community mental health teams and outpatient services | Depression (n=167) | Service users (n=167) | Video call |
| Colle et al. (2020) France | Assessing the short-term acceptability of teleconsultation during the COVID-19 pandemic. | Letter: Quantitative cross-sectional study | Community mental health teams and outpatient services | Psychosis and bipolar, depression, anxiety, obsessive-compulsive disorder, stress, and substance misuse | Psychiatrists (n=17) | Video call, Phone call |
| Conn et al. (2013) Canada | Evaluating a telepsychiatry programme for older adults. | Service evaluation/audit: Quantitative retrospective and cross-sectional cohort study and qualitative focus group study | Community mental health teams and outpatient services | Mixed secondary mental health service users | Physicians (n=29, in survey), mixed groups of mental health professionals (n in focus groups unclear), service users (n=100 in retrospective chart review; n=253 sessional evaluation forms of patient assessments; n=41 case consultations) | Video call |
| Connolly et al. (2021) USA | Describing Veterans Affairs' rapid increase in telemental health due to COVID-19. | Service evaluation/audit | Community mental health teams and outpatient services | NA | NA (description of service change) | Video call, Phone call |

| | | | | | | |
|---|---|---|---|---|---|---|
| Costa et al. (2021) USA | Assessing resilience and mental health care during the COVID-19 pandemic, with a focus on telemental health. | Quantitative cross-sectional study and qualitative survey | Service user-led/peer support | Mixed primary care, psychology/psychotherapy, or secondary care service users | Service users (n=435 of which n=381 had self-reported mental illness) | Video call, Phone call |
| Crowe et al. (2016) USA | Comparing in person and telepsychiatry for deaf or hard of hearing patients. | Quantitative non-randomised controlled trial (nRCT) | Community mental health teams and outpatient services | Mixed secondary mental health service users | Service users (n=24) | Video call |
| Disney et al. (2021) USA | Evaluating the transition of a refugee-serving mental health clinic to telemental health as well as perceived obstacles and helpful resources. | Qualitative survey | Refugee-serving outpatient mental health clinic in a large refugee resettlement area | NA | Mixed groups of mental health professionals (n=17) | Phone call, or phone call in combination with video call (did not use video call alone) |
| Dores et al. (2020) Portugal | Analysing 1) changes in professionals' use of and attitudes towards Information and Communication Technologies during the first COVID-19 lockdown; 2) factors underlying such changes; and 3) possible adoption of guidelines. | Quantitative cross-sectional study | Psychology/psychotherapy/counselling service | Depression, anxiety, neurodevelopmental, "personality disorders", substance use disorders, sleep disorder, neurocognitive disorders | Psychologists (n=108) | Video call, Phone call, Email, Other (audio conferences, online intervention platforms, smartphones and tablet apps, online forums, chats, short-message services, virtual rooms), Text message/SMS/WhatsApp |
| Eagle (2020) UK | Exploring what has been advantageous and disadvantageous about remote therapy. | Service evaluation/audit and Primary research study: Qualitative interview study | Community mental health teams and outpatient services | Mixed secondary mental health service users | Service users (n=8) | Phone call, video call |
| Fogler et al. (2020) USA | Describing how children's hospitals used treatment fidelity to guide adaptations to an ADHD bootcamp for parents. | Service evaluation/audit: Quantitative cross-sectional study and qualitative focus group study and survey | Tertiary care children's hospitals | ADHD | Caregivers of 20 children with ADHD | Video call |

| | | | | | | |
|---|---|---|---|---|---|---|
| Foye et al. (2020) UK | Exploring how Covid-19 has affected the mental health nurse workforce. | Quantitative cross-sectional study and qualitative survey | Community mental health teams and outpatient services; Crisis and emergency mental health services; adult and older adult services; children and young people; intellectual disabilities; forensic services; perinatal; drugs and alcohol services eating disorders | NA | Mental health nurses (n=897) | Video call, Phone call |
| Frayn et al. 2021 USA | Examining the impact of COVID-19 on people with binge eating spectrum disorders and their perception of tele-therapy. | Qualitative interview study | Community mental health teams and outpatient services | Eating disorders | Service users (n=11) | Video call |
| Gaddy et al. (2020) USA | Assessing the impact of the pandemic on the employment and service delivery of music therapy professionals. | Quantitative cross-sectional study and qualitative survey | Music therapy provided at schools, hospice, inpatient/outpatient psychiatry and private practice/contractual | Autism spectrum disorder, developmental disabilities, Alzheimer's disorder | Music therapists (n=1,196) | Video call, Phone call, Other (pre-recorded songs/playlists and video sessions) |
| Ghaneirad et al. (2021) Germany | Examining therapists' and service users' experiences of video consultations during the first COVID-19 lockdown. | Quantitative cross-sectional study | Community mental health teams and outpatient services | Mixed primary care or psychology/psychotherapy service users | Service users (n=338) | Video call |
| Ghosh et al. (2021) India | Describing the stepwise telemedicine model adopted by a Drug De-addiction and Treatment Centre. | Service evaluation/audit | Substance use clinic | Substance use disorder | Service users (n=379) | Video call, Phone call, Mixed F2F and telemental health |
| Godleski et al. (2012) USA | Assessing the feasibility and outcomes of the home electronic messaging programme for psychiatric patients. | Quantitative before-and-after study | Veterans' healthcare community service | Schizophrenia, PTSD, depression and substance-use disorder | Service users (n=76) | Other (Electronic messaging device) |

| | | | | | | |
|---|---|---|---|---|---|---|
| Greenwood et al. (2004) Australia | Evaluating a telepsychiatry clinical service in rural New South Wales. | Service evaluation/audit: Quantitative cross-sectional study | Satellite clinic in a rural site to bring specialized mood disorder | Mood and anxiety disorder | Service users (n=20) | Video call, Phone call, Mixed F2F and telemental health |
| Grover et al. (2020a) India | Evaluating the impact of the COVID-19 pandemic on mental health services. | Quantitative cross-sectional study | Community mental health teams and outpatient services; Inpatient mental health service; state-funded government medical colleges; private medical colleges; central government-funded institutes | Mixed primary or secondary care, or psychology/psychotherapy service users | Professors or heads of department from various medical colleges (n=109) | Video call, Phone call |
| Grover et al. (2020b) India | Evaluating the impact of the Covid-19 pandemic and lockdown on the mental health services in the private sector. | Quantitative cross-sectional study | Mental health services in the private sector (single chamber outpatient practice; mental health inpatient setting owned by professionals; mental health service provided through the corporate hospitals; multi-specialty hospitals) | General population in quarantine; mixed primary or secondary care, or psychology/psychotherapy service users | Members of the Indian Psychiatric Society working in the private mental health sector (n=396) | Video call, Phone call |
| Guinart et al. (2020) USA | Examining patients' experiences of, and attitudes toward, telepsychiatry. | Quantitative cross-sectional study | Community mental health teams and outpatient services; General hospital/physical health service | NA | Service users (n=3,070) | Video call, Phone call, other (combination) |
| Hawke et al. (2021) Canada | Examining youth's attitudes toward and experiences of virtual mental health services and substance use services. | Quantitative cross-sectional study and qualitative survey | Community mental health teams and outpatient services (CAMH specialty clinics) and no specific setting/general | Mixed primary or secondary care, or psychology/psychotherapy service users | Service users (n=164), general population (n=245) | Video call, Phone call, Email, Text message/SMS/WhatsApp |

| | | | | | | |
|---|---|---|---|---|---|---|
| | | | population | | | |
| He et al. (2020) China | Evaluating the 'The COVID-19 Psychological Resilience Model' intervention. | Quantitative cross-sectional study | No specific setting/general population | NA | NA (intervention description) | Video call, Phone call, Mixed F2F and telemental health |
| Healthwatch (2021) UK | Exploring the barriers to accessing mental health appointments by phone or video call. | Quantitative cross-sectional study and qualitative focus group study and Interview study | No specific setting | Mixed primary care or psychology/psychotherapy service users | Service users (n=98) | Video call, Phone call |
| Hensel et al. (2021) Canada | Exploring "how videoconferencing should be used and for whom" with reference to postpartum women. | Qualitative interview study | Psychology/psychotherapy/counselling service within perinatal mental health service | Postpartum depression and anxiety | Therapists (n=3), service users (n=12) | Video call, Mixed F2F and telemental health |
| Hernandez-Tejada et al. (2014) USA | Investigating parameters and barriers to treatment completion among veterans receiving exposure therapies in-person compared to telemedicine. | Quantitative case–control study using secondary data | Not stated/unclear | PTSD | Veterans (n=47) | Phone call |
| Holland et al. (2020) UK | Providing guidance for remote working in mental health settings | NHS guidance document | No specific setting | NA | Not applicable (guidance document) | Video call, Phone call, Mixed F2F and telemental health |
| Hopkins and Pedwell (2021) Australia | Assessing the nature and impact of changes in child and youth mental health services. | Quantitative cross-sectional study and qualitative survey | Child and youth psychology/psychotherapy/counselling services | NA | Mixed groups of mental health professionals (n=113) | Video call, Phone call |
| Humer et al. (2020) Austria | Evaluating how well psychotherapists feel informed about the use of the Internet in psychotherapy and which software was used during the | Quantitative cross-sectional study | Not stated/unclear | NA | Psychotherapists (n=1,547) | Video call |

| | | | | | | |
|---|---|---|---|---|---|---|
| | COVID-19 lockdown for online psychotherapy. | | | | | |
| Johnson et al. (2021) UK | Describing the perspectives and experiences of staff working in inpatient and community settings. | Quantitative cross-sectional study and qualitative survey | NHS, private healthcare, social care, and voluntary sector services | Mixed primary care or psychology/psychotherapy service users, mixed secondary mental health service users, general population | Staff (n=2,180) | Video call, Phone call, Mixed F2F and telemental health |
| Jones et al. (2014) USA | Evaluating a technology enhanced version of behavioural parent training for children with disruptive behaviour from low-income families. | Quantitative randomised controlled trial (Pilot RCT) | No specific setting | Childhood disruptive behaviour | Families with low income (n=19) | Video call, Text message/SMS/Whats App |
| Juarez-Reyes et al. (2021) USA | Describing procedures, acceptability, and feasibility of converting from in-person to videoconference sessions. | Quantitative before-and-after study and qualitative focus group | University-based primary care clinic | Depression, anxiety, and/or stress | Service users (n=6) | Mixed F2F and telemental health |
| Kanellopoulos et al. (2021) USA | Describing perceived benefits and challenges of an inpatient Telemental Health conversion. | Letter with data on a psychiatric inpatient service | Inpatient mental health service | NA | Inpatients (n=15) | Video call, Phone call |
| Khanna et al. (2020) Australia | Investigating flexible work arrangements. | Service evaluation/audit | Community mental health teams and outpatient services | PTSD | Psychiatrists (n=4), registrars (n=3), psychologists (n=14) | Video call, Phone call, Mixed F2F and telemental health |
| King et al. (2006) Australia | Examining adolescents' motivations and experiences in accessing the Internet for counselling services. | Qualitative focus group study | No specific setting/general population | Mixed primary care or psychology/psychotherapy service users | Service users and general population (n=39) | Text message/SMS/Whats App |
| Lakeman and Crighton (2021) Australia | Exploring the impacts of the cessation of DBT programmes and obstacles and solutions to providing DBT using telehealth technology. | Quantitative cross-sectional study and qualitative survey | 12 hospitals and 21 community health centres | "Borderline personality disorder" diagnosis | Mixed group of mental health staff (n=28) | Video call, Phone call, Mixed F2F and telemental health |

| | | | | | | |
|---|---|---|---|---|---|---|
| Lecomte et al. (2020) Canada | Determining the feasibility and acceptability of CBT for first episode psychosis via videoconferencing. | Quantitative before-and-after study and qualitative interview study | Early intervention for psychosis service | Schizophrenia-spectrum disorders, bipolar disorder | Service users (n=14) | Video call |
| Liberati et al. (2021) UK | Exploring the experiences of service users, carers and staff seeking or providing secondary mental health services during the pandemic. | Qualitative interview study | Community mental health teams and outpatient services (early intervention for psychosis); Inpatient mental health service; Crisis and emergency mental health services; secure forensic services; specialist services | Mixed secondary mental health service users | Mixed mental health staff (n=35), service users (n=20), carers of people with mental health difficulties (n=10) | Video call, Phone call, Email, Text message/SMS/Whats App, Mixed F2F and telemental health |
| Lichstein et al. (2013) USA | Exploring the efficacy of telehealth targeting insomnia and depression in older adults in rural primary care settings. | Quantitative before-and-after study | Primary care CBT counselling service | Depression, comorbid insomnia | Service users (n=5) | Video call, Phone call |
| Lin et al. (2020) China | Evaluating the quality of a psychological hotline services. | Quantitative cross-sectional study | Helplines | NA | Staff (n=414) | Phone call, Text message/SMS/Whats App |
| Lindsay et al. (2015) USA | Investigating the implementation of video telehealth for the delivery of psychotherapy for veterans with PTSD. | Quantitative before-and-after study | Department of Veteran Affairs Medical Clinics | NA | 5 Veteran Affairs Medical Clinics | Video call |
| Lodder et al. (2020) UK | Evaluating procedural and implementation challenges of a blended, group-based psychosocial stigma protection intervention for parents of autistic children. | Qualitative focus group study and survey | Not stated/unclear | Autism | Children (n=9) and their parents (n=10) | Video call |
| Mad Covid (2020) UK | Discussing the impact COVID-19 has had on people with pre-existing mental health conditions and how mental | Service user led report: Qualitative focus group study and participant observation | No specific setting | Mixed primary care or psychology/psychotherapy service users and mixed secondary | Service users, survivors, allies (4 panel members and | Video call, phone call |

| | | | | | | |
|---|---|---|---|---|---|---|
| | health services can support them. | | | mental health service users | 67 symposium attendees) | |
| Mahmoud et al. (2021) USA | Examining approaches implemented by a telepsychiatry organisation to increase satisfaction, engagement and wellbeing among telepsychiatrists. | Quantitative cross-sectional study | Community mental health teams and outpatient services | NA | Telepsychiatrists (n=25 in 2018, n=23 in 2019) | Video call, Phone call, Email |
| Martin et al. (2020) UK | Investigating the role of digital communications in mental health services. | Qualitative interview study (secondary analysis) | Early intervention psychosis clinics | Psychosis and bipolar | Mixed mental health staff (n=19), youth service users (n=5) | Email, Text message/SMS/Whats App |
| McBeath et al. (2020) UK | Exploring the experiences and challenges of psychotherapists working remotely during the COVID-19 pandemic. | Quantitative cross-sectional study and qualitative survey | Not stated/unclear | NA | Psychotherapists (n=335) | Video call, Phone call, Email, Other (letters), Text message/SMS/Whats App |
| Medalia et al. (2020) USA | Describing key factors addressed by a clinic for adults with service mental illness to continue to provide services via video-supported telehealth. | Commentary/editorial with data | Community mental health teams and outpatient services | Severe mental illness | NA (description of service change) | Video call |
| Mental Health Commission of Canada (2014) Canada | Providing an overview of e-Mental health in Canada, including its challenges and barriers. | Briefing document for e-mental health care in Canada | No specific setting | NA | NA (briefing document) | Video call, Phone call, Email, Text message/SMS/Whats App, Other |
| Mental Health Network NHS Confederation (2020a) UK | Describing the experiences of two Mental Health Trusts delivering group sessions. | Briefing paper based on the experiences of two Mental Health Trusts delivering Group sessions | Community mental health teams and outpatient service; Psychology/psychotherapy/counselling service | Mixed secondary mental health service users | NA (briefing paper) | Video call, Phone call |
| Mental Health | Providing guidance for increasing service user choice in | NHS guidance document | No specific setting | NA | Not applicable (guidance document) | Video call, Phone call |

| | | | | | | |
|---|---|---|---|---|---|---|
| Network NHS Confederation (2020b) UK | and access to digital mental health care. | | | | | |
| Mind (2021a) UK | Reporting people's experiences of being offered and using support from the NHS for their mental health by phone or online. | Quantitative cross-sectional survey and qualitative focus group and interview study | Mental health services (No specific setting) | NA | Mixed groups of mental health professionals (n=4 interviews), individuals offered NHS mental health support (n=1,914 survey participants, n=11 interviews); Mind's 'community activist' campaigners (focus group, n not stated) | Video call, Phone call, Text message/SMS/Whats App |
| Mind (2021b) UK | Reporting people's experiences of accessing support from the NHS for their mental health by phone or online. | Quantitative cross-sectional survey and qualitative interviews | Mental health services (No specific setting) | NA | General population accessing NHS mental health services (n=approximately 1,900 survey participants; n=5 interviews) | Phone call, video call |
| Moore (2021) UK | Presenting the challenges of people severely affected by mental illness have faced during the COVID-19 pandemic. | Briefing report: Quantitative cross-sectional study | Early intervention in psychosis & Liaison and diversion services | Severe mental illness | People with mental illness (n=272) | Video call, Phone call, Text message/SMS/Whats App |
| Moslehi et al. (2021) UK | Describing the Attend Anywhere platform, used in an inpatient psychiatric unit in Camden & Islington during the pandemic. | Service description | Inpatient mental health service | Mixed secondary mental health service users | NA (description of service change) | Video call |

| | | | | | | |
|---|---|---|---|---|---|---|
| Newbronner et al. (2021) UK | Exploring the satisfaction with and difficulties accessing the support provided during the pandemic among people with severe mental illnesses | Quantitative cross-sectional study and qualitative survey | Primary care mental health trusts | Schizophrenia or delusional/psychotic illness or bipolar disorder | Service users (n=367) | Phone call, Text message/SMS/Whats App |
| National Health Service England (accessed 2022) UK | Describing the adoption of crisis lines in NHS services. | NHSE case studies of 24/7 crisis lines provided during COVID | 24/7 crisis helplines | NA | Not applicable (description of service change) | Phone call |
| Nissling et al. (2020) Sweden | Assessing the experiences of patients with anxiety disorders as well as the feasibility, safety, accessibility and effectiveness of an internet-CBT program. | Quantitative before-and-after study and qualitative interview study | Primary care | Anxiety disorder | Service users (n=9 for quantitative measures, n=8 for qualitative follow-up) | Phone call, Text message/SMS/Whats App |
| O'Dell et al. (2021) USA | Reporting the acceptability and usability of an InTouch platform used for Tele Behavioural Health | Service evaluation/audit: Quantitative cohort study (retrospective and cross-sectional) | Primary care | NA | NA (description of service change) | Text message/SMS/Whats App, Phone call |
| Ogueji et al. (2021) Nigeria | Exploring the willingness to utilise e-therapy services and identify its limitations in Nigeria. | Qualitative survey | No specific setting | | General population (n=100) | Video call, Phone call, Text message/SMS/Whats App |
| Olwill et al. (2020) Ireland | Reporting Irish psychiatrists' experience of remote consultations at a Department of Old Age Psychiatry. | Quantitative cross-sectional study and qualitative survey | Outpatient psychiatric service, Department of Old Age Psychiatry | Mixed secondary mental health service users | Non-consultant hospital doctor and psychiatric consultants (n=26) | Phone call |
| Open Excellence (2020) USA | Publicising online hearing voices groups. | Publicity / news article | Service user-led/peer support: Hearing voices groups | Psychosis and bipolar | Peer workers (n=2) | Video call |
| Orlowski et al. (2016) Australia | Reporting the receptiveness and readiness of the youth mental health workforce towards the use of mobile and | Qualitative focus group study and Interview study | Youth Mental Health services | NA | Mixed mental health staff (n=40 in focus groups, n=8 interviewed) | Email, Facebook chat/messaging, Text message/SMS/Whats App |

| | | | | | | |
|---|---|---|---|---|---|---|
| | online-based technologies for young people. | | | | | |
| Peralta and Taveras (2020) Dominican Republic | Determining the effectiveness of teleconsultation during the pandemic in the Dominican Republic. | Quantitative cross-sectional study | Helplines | General population | NA (6,800 interventions) | Video call, Phone call, Text message/SMS/Whats App |
| Pierce et al. (2021) USA | Reporting the effect of the COVID-19 pandemic on the extend of telepsychology use. | Quantitative cross-sectional study | Community mental health teams and outpatient services; residential services; general hospital/physical health service; Psychology/psychotherapy/counselling service | Mixed primary care or psychology/psychotherapy service users | Licensed psychologists (n=2,619) | Video call, Phone call |
| Pote et al. (2021) UK | Reporting digital mental health competencies and training needed by UK clinical psychologists including opportunities and barriers. | Quantitative cross-sectional study and qualitative interview study | Psychology/psychotherapy/counselling service | NA | UK-wide Doctoral programmes in Clinical Psychology (n=18) | Video call, Email, Text message/SMS/Whats App |
| Pugh et al. (2020) UK | Investigation how chairwork is best provided in internet-delivered psychotherapy. | Qualitative survey | Not stated/unclear | NA | Mixed groups of mental health professionals (n=41) | Video call, Phone call |
| Rabinowitz et al. (2010) USA | Describing the development, implementation, and benefits of a nursing home Telepsychiatry consultation service. | Quantitative cohort study (retrospective) | Nursing home | Depression, dementia, other mental health disorders | Service users (n=106) | Video call, Phone call |
| Rosen et al. (2021) USA | Describing barriers, facilitators, and challenges of Veteran's Health Administratoin expansion of telemental health care during the COVID-19 pandemic. | Service evaluation/audit | Veteran's Health Administration mental health services in USA | Mixed mental health problems | NA (description of service change) | Video call, Phone call |

| | | | | | | |
|---|---|---|---|---|---|---|
| Sasangohar et al. (2020) USA | Reporting the rapid implementation of telepsychiatry in a psychiatric practice in Texas. | Service evaluation/audit | Community mental health teams and outpatient services | Psychosis, bipolar, depression, anxiety, PTSD, other mental disorders | NA (description of service change) | Video call, Phone call, Text message/SMS/Whats App |
| Scharff et al. (2020) USA | Reporting the steps taken to shift to remote training and practice by a Psychological Services Centre. | Service evaluation/audit | Community based training clinic providing therapy | General population and individuals with anxiety and other mental disorders | NA (description of service change) | Video call |
| Schueller et al. (2019) USA | Review the use of digital mental health interventions in marginalised and underserved populations. | Review | No specific setting | NA | NA (review) | General telemental health |
| Sehlo et al. (2021) Egypt | Comparing patient views on F2F consultation and telepsychiatry in Egypt during the pandemic. | Quantitative cross-sectional study | Online psychological therapy service | Mixed primary care or psychology/psychotherapy service users | Service users (n=183) | Mixed F2F and telemental health |
| Severe et al. (2020) USA | Examining factors influencing patients' decisions to accept or decline telepsychiatry and their choice of telemental health modality. | Quantitative cross-sectional study and qualitative survey | Community mental health teams and outpatient services | Mixed secondary mental health service users | Service users (n=44) | Video call, Phone call, Other (pause/postpone care) |
| Sheehan et al. (2020) UK | Reporting the experiences of staff working across different mental health services for people with intellectual and other developmental disabilities. | Quantitative cross-sectional study and qualitative survey | Not stated/unclear | Intellectual and developmental disabilities | Mental health professionals working with people with intellectual disabilities and/or developmental disabilities (n=648) | NA |
| Shklarski et al. (2021) USA | Assessing psychotherapists' challenges and adaptations to providing remote therapy from home during the Covid-19 pandemic. | Quantitative cross-sectional study and qualitative interview study | No specific setting/general population | NA | Mixed mental health professionals (n=92 in survey, n=19 in interviews) | General telemental health |
| Shore et al. (2014) USA | Describing the Home-Based Telemental program and examining the feasibility of | Quantitative cross-sectional study | Veteran medical centre | Depression, PTSD, psychosis and bipolar disorder, | Veteran service users (n=38) | Video call |

| | | | | | | |
|---|---|---|---|---|---|---|
| | providing online mental healthcare to veterans at home. | | | anxiety, other mental disorders | | |
| Simon et al. (2021) UK | Exploring the views of NHS commissioners and managers on internet-based therapies and their implementation. | Qualitative interview study | No specific setting | | Individuals working in the NHS (n=10) | Video call, Phone call |
| Simpson et al. (2001a) Canada | Evaluating the provision of routine psychiatric care to a rural community via video call. | Service evaluation/audit and primary research study: Quantitative cross-sectional study and qualitative interview and focus group study | Outpatient telepsychiatry consultations delivered from a psychiatric hospital to local general hospitals | Psychosis and bipolar, depression, dementia, other mental health disorders | Mental health clinic staff (n=13 in focus group), psychiatrists (n=3 in interviews) | Video call, Mixed F2F and telemental health |
| Simpson et al. (2001b) Canada | Reporting patient experiences of a routine telepsychiatry service in Alberta, Canada. | Quantitative cross-sectional study | Tertiary-care psychiatry service | Mixed secondary mental health service users and service users with depression | Service users (n=230 in surveys, n=31 in structured telephone interviews) | Video call |
| Smith and Gillon (2021) UK | Exploring therapists’ views and experiences of processes and efficacy of online counselling. | Qualitative interview study | No specific setting | NA | Counsellors and Psychotherapist or analyst (n=5) | Video call, Phone call |
| The British Psychologica l Society (2020a) UK | Providing guidance for psychologists working with children and young people using online video platforms. | Guidance document | No specific setting | NA | Not applicable (guidance document) | Video call |
| The British Psychologica l Society, (2020b) UK | Providing guidance for working therapeutically with parents and their infants during pregnancy and postpartum using remote delivery platforms. | Guidance document | No specific setting | Prenatal and postnatal mental health disorders | NA (guidance document) | Video call, |
| The Health Foundation (2020) UK | Evaluating “Connecting With Telehealth to Children in Hospital” (CWTCH), a NHS telepsychiatry project within Child | Service evaluation/audit: Quantitative cross-sectional study and | Community mental health teams and outpatient services | NA | Patients, families, clinicians, wider public (n not reported) | Video call |

| | | | | | | |
|---|---|---|---|---|---|---|
| | and Adolescent Mental Health Service (CAMHS). | qualitative survey and interview study | | | | |
| The Mental Health Innovation Network (2020) USA | Providing guidance for supporting refugee mental health during COVID-19. | Webpage with relevant information | No specific setting | NA | Not applicable (guidance webpage) | Text messages |
| Uscher-Pines et al. (2020a) USA | Understanding how the shift in delivery modes has affected mental health care during the COVID-19 pandemic. | Qualitative interview study | Hospital outpatient clinic; Private practice; other setting | NA | Outpatient psychiatrists (n=20) | Video call, Phone call |
| Uscher-Pines et al. (2020b) USA | Describing how clinicians used telemedicine for Opiate Use Disorder in conjunction with in-person care during the early stages of the COVID-19 pandemic. | Qualitative interview study | Community mental health teams and outpatient services; Private hospital/clinic; General hospital/physical health service | Opioid use disorder | Clinicians (n=18) | Video call, Phone call |
| Vera San Juan et al. (2021) UK | Investigating the experiences of service users during the switch to telemental health care. | Qualitative interview study | Mixed mental health services (outpatient; crisis and emergency; inpatient; voluntary sector/non-profit; psychology/psychotherapy/counselling services; general hospital/physical health service ) | Mixed primary care or psychology/psychotherapy service users and mixed secondary mental health service users | Service users (n=44) | Video call, Phone call, Text message/SMS/WhatsApp |
| Watson et al. (2021) UK | Investigating which factors influence the uptake of remote therapy among individuals with psychosis. | Quantitative cross-sectional study | Psychology/psychotherapy/counselling service | Psychosis and associated emotional difficulties | Service users (n=51) | General telemental health |
| Whistance et al. (2020) UK | Evaluating the use and value of video consulting, including satisfaction, clinical suitability, and acceptability. | Service evaluation/audit: Quantitative cross-sectional study and qualitative survey | Mixed mental health settings (primary, secondary and community care) | Mixed primary care or psychology/psychotherapy service users and mixed secondary | Mixed groups of mental health professionals (n=6,090), service users (n=4,311) | Video call |

| | | | | | | |
|---|---|---|---|---|---|---|
| | | | | mental health service users | | |
| Williams et al. (2017) UK | Investigating the views of people who are homeless or insecurely housed on telemental health. | Quantitative cross-sectional study and qualitative focus group study and interview study | Local open access homeless and wellbeing daycentre | NA | Service users (N=64: n=51 interviewees, n=13 focus group members) | General telemental health |
| Wilson et al. (2021) UK | Exploring staff perceptions of the impact of the COVID-19 pandemic on mental health service delivery and outcomes for perinatal women. | Quantitative cross-sectional study and qualitative survey | Community mental health teams; Crisis and emergency mental health services*;* inpatient mental health services; perinatal services | Perinatal mental health problems | Mixed clinical staff working with women in the perinatal period (n=363) | Video call, Phone call, Mixed F2F and telemental health |
| Wyler et al. (2021) Switzerland | Exploring how adult patients with ADHD and their therapists experienced different modalities of therapy during the COVID-19 pandemic. | Quantitative cross-sectional study and quantitative survey | Community mental health teams and outpatient services | ADHD | Staff and service users (n=60 therapist/service user dyads) | Video call, Phone call, Other (F2F with the therapist wearing a face mask) |
| Yellowlees et al. (2020) USA | Describing the process, challenges, and lessons learned from the rapid conversion of an outpatient psychiatric clinic to a virtual telepsychiatry clinic. | Commentary/editorial with data | General hospital/physical health service | NA | NA (description of service change) | Video call, Phone call |
| YoungMinds (2020) UK | Exploring experiences of children and young people of using digital technology. | Discussion (Submission to COVID Committee's inquiry) | No specific setting | General population | NA | General telemental health |
| Zheng and Gray (2014) USA | Examining the feasibility of empirically supported treatments implemented via videoconferencing. | Case studies | Psychology Telehealth Clinic at the University of Wyoming | PTSD | Service users (n=2) | Video call |

#### Appendix 2. Overarching CMOs

This document contains the underlying CMOs and stakeholder consultation notes that contributed to the development of the overarching CMOs, which were iteratively refined through a process of stakeholder and research team consultation and discussion. The references for the underlying CMOs can be found in the reference section of the main paper, and further information on each of the studies can be found in the Study Characteristics table.

### Domain 1. Connecting Effectively

##### CMO 1.1: Providing access to digital devices

When service users who do not have access to digital devices are given access to up-to-date devices (and chargers), paid for or loaned to them (C), this results in improved access to and implementation of telemental health services (especially via video platforms) (O1), some inequalities in accessing a digital device are addressed (O2), exacerbation of existing inequalities is less likely (O3), and service users are more able to maintain personal contact with family and friends if they wish, and to access a range of online services (O4), as this reduces the burden of having to purchase a device for the service users, and provides more financially viable access to devices required for online connections (M).

**Underlying CMOs:**

- Bierbooms et al. (2020) Mental health care professionals in community mental health services in the Netherlands (C) who reported technical issues and lack of technical devices/internet connection, lack of privacy and confidence in security of the system (M) reported frustration and less positive attitudes towards telemental health (O).
- Chen et al. (2020) In a general hospital outpatient psychiatry service in the USA during Covid-19 (C), it was suggested that many patients may not have access to appropriate technology to facilitate video-based telehealth platforms (M), as 30% of the virtual visits completed in the department were delivered via phone (between March and April 2020) (O).
- Disney et al. (2021) Refugee clients who are at risk of digital exclusion (C) often did not have access to appropriate technological devices or internet data or reliable Wi-Fi (M), so struggled to access telemental health (O).
- Ghaneirad et al. (2021) When offering video consultations to service users who lack technological devices (C), if insurance companies provide support (M) then service users can access care (O).
- Ghosh et al. (2021) Among service users of a substance disorder centre in India with a stepped telemental health model who needed to access videocalls for mental health consultations (C), many did not have access to smartphones or computers, good internet connectivity, and a minimum technical expertise (M), which limited their ability to engage with services via video call.
- Johnson et al. (2021) When moving to telemental health (C), if resources are inadequate (equipment and internet connections are of low quality, processes and preferred platforms are not clearly established, and/or staff lack training and confidence) (M), then remote working works less well (O).
- Johnson et al. (2021) When moving to telemental health (C), if service users are digitally excluded (lack equipment, resources and/or confidence to connect or live in overcrowded housing) (M), then remote working works less well (O).
- Kanellopoulos et al. (2021) When a psychiatric inpatient service provided electronic tablets to each of the patients who were isolated in their rooms during a rapid switch to telemental health due to COVID-19 (C), patients continued to receive multidisciplinary treatment as well as support from their family and friends while reducing the risk of COVID-19 infections (O), as they were

able to attend meetings with the treatment team, virtual family visits, and to access therapeutic applications and content (M).

- Lakeman and Crighton (2021) Among staff working in psychology / psychotherapy or counselling services for people with personality disorder in Australia when COVID-19 forced an abrupt cessation of their programmes (C), if/when there was a lack of support for resources (such as adequate technology) (M), then there were issues connecting with service users (O).
- Lakeman and Crighton (2021) Among service users with personality disorder receiving psychology / psychotherapy or counselling services in Australia when COVID-19 forced an abrupt cessation of their programmes (C), if/when there was a lack of technology and lack of data to hold a group video call (M), THEN this form of support was unsustainable for many clients (O).
- The Mental Health Innovation Network (2020) For refugees who wanted to access telemental health support during COVID-19 (C), lack of access to technology or poor technological literacy (M), made implementation difficult (O).
- Mind (2021a) For people engaged in online therapy (C), the financial burden of having to purchase new technology (M) acted as an additional stress to existing mental health issues (O).
- Mental Health Network NHS Confederation (2020b) Among individuals who did not have access to a mobile phone, tablet or computer (C), when Mental Health Matters purchased pay-as-you-go phones for those with no existing means of contact (M1), and staff supported individuals with training on how to use these (including installing appropriate apps) (M2), then people were able to maintain contact with family and friends over the lockdown (O).
- Mental Health Network NHS Confederation (2020b) In order to improve digital inclusion for those with a lack of access to equipment (C), the City and Hackney Clinical Commissioning Group fast tracked a ‘stay connected’ personal health budget (M), which facilitated quick access to a choice of mobile phones and/or sim cards and has enabled 120 people to access and take part in remote support services, join online community activities, access resources and support online, and stay in touch with family and friends during lockdown (O).
- Mental Health Network NHS Confederation (2020b) In order to support health and social care technologies in Liverpool (C), the Liverpool 5G Consortium was awarded £4.3million by the Department for Digital, Media, Culture and Sport as part of a £7.2 million project Liverpool 5G Create: Connecting Health and Social Care (M), which will reduce digital poverty for vulnerable people in need by providing safe, free and accessible connectivity to services including health, social care and education (O).
- Simpson et al. (2001a) Among patients in rural Canada using a telepsychiatry service linking 5 local general hospitals with a specialist psychiatric hospital (C), if the quality of the equipment and room were good 96-99% of the time (M), 92% felt they were able to present the same information via video call as in person (O).
- Watson et al. (2021) Among service users with psychosis accessing psychological therapies before COVID-19 (C), if they did not own the right technology or could not afford the internet (M), then they declined remote therapy (O).

**Supporting information from stakeholder consultation:**

- Digital exclusion is an element of deprivation that shouldn’t be overlooked or underestimated: it may exacerbate other forms of inequality and deprivation.
- A variety of organisations (NHS, local authority, UK) are funding initiatives to provide devices: they may perceive this as cheaper than service users attending face-to-face.
- Service users including young people might not be aware of relevant initiatives to help them get connected, so these should be promoted.
- There is probably some cost-benefit analysis (by managers, trusts, commissioners) required to establish how to make device provision sustainable going forward, while preserving the quality of the service. There is a drive to shorten waiting lists and save money - perhaps telemental health can support these things.

- There are some service users who simply do not want to engage via technology, even if they could access it without issue, they are not ‘digitally excluded’ but they will not be receiving telemental health. Choice should be respected.

**Caveats and pitfalls identified by stakeholders:**

- Provision of devices does not necessarily provide a solution if there is no data included or possibility of home broadband. Some service users live in accommodation without phone lines. Small data packages are easily used up, for example, when they use the device to watch television, and additional data may be expensive.
- Important to highlight that these solutions will not result in the same outcomes for everyone. For example, those who are estranged from family and friends will continue to lack a social support network, and buying them a device to connect will not change that.
- Even when service users are given devices, they still need to ensure they are charged. There are issues with this when service users are unable to pay for electricity, cannot access charging points (e.g. homeless people) or are on inpatient wards where access to charging facilities may be challenging. Service users may rely on staff to charge their devices for them, but then cannot check that they are charging properly (e.g. for old phones which may need adjusting to charge fully).
- Additional consideration that the specification of the device/device software needs to match the application/programme that needs to run. The devices need to be up to date. If the device is not compatible then this limits the service user’s options and increases inequalities as an intervention can be offered to some people and not others.
- Staff do not always have their own computers in their office (or computers with webcams), and are having to use personal devices in lieu; provision of devices may apply to staff also.
- NHS phones do not always allow apps – technology in the NHS is behind what is needed.

#### CMO 1.2: Lack of access to stable, secure, and/or adequate internet connection

a. When staff deliver telemental health via video from workplaces or homes with unstable and poor internet connection (C), tele-consultations are difficult (or impossible) to conduct with service users (O1), fewer tele-consultations are conducted (O2) and telemental health is viewed less positively (O3), as staff experience frustration and there is reduced motivation to arrange online appointments (M).
b. When service users only have access to an insecure, unstable and poor-quality internet connection, and/or consistent technological problems (C), it is difficult for telemental health to be viewed positively (O1) they are able and/or willing to accept fewer online consultations (O2), may continue to struggle with their mental health (if face-to-face consultations are unavailable) (O3) and this may result in digital exclusion which could exacerbate existing inequalities (O4), as the service users struggle to engage in sessions with sufficient clarity and mutual comprehension, and experience frustration (M).

**Underlying CMOs:**

- Costa et al. (2021) During the COVID-19 pandemic in the USA, mental health service users who said that they were coping poorly with the pandemic (C) reported not feeling comfortable using telemental health, not having been able to connect to their provider, feeling that there was a decrease in number of sessions, challenges in using technology, and that the phone was less effective (M) and therefore in telemental health reduced access to care (O).

- Bierbooms et al. (2020) Mental health care professionals in community mental health services in the Netherlands (C) who reported technical issues and lack of technical devices/internet connection, lack of privacy and confidence in security of the system (M) reported frustration and less positive attitudes towards telemental health (O).
- Bommersbach et al. (2021) Having poor Wi-Fi (C) made it difficult for staff to perform their job duties (M), which caused frustration when using technology (inferred) (O).
- Buckman et al. 2021 When providing care using videoconferencing (C), a lack of appropriate technology or stable Wi-Fi connection (M) impacted staff's abilities to provide video sessions (O).
- Foye et al. (2020) Amongst mental health nurses who had to adapt quickly to remote working during the COVID-19 pandemic (C), many reported experiencing logistical challenges such as poor internet connection, hardware availability, and patient lack of privacy to conduct appointments (M) which negatively affected their ability to work and deliver care to service users (O).
- Frayn et al. (2021) For people experiencing binge eating spectrum disorders (BESD) attending virtual therapy (C), technical barriers and "logistical issues" related to home environment, e.g. internet connections, lack of privacy (M), may “hinder” engagement with therapy (O).
- Ghaneirad et al. (2021) When offering video consultations to service users due to the pandemic (C), one third of service users did not take up video consultations (O) because they lacked necessary technological devices (32%), experienced technological difficulties during the first session (19%), or did not want to attend video consultations despite functioning technological devices (49%) (M).
- Ghosh et al. (2021) Among service users of a substance disorder centre in India with a stepped telemental health model who needed to access videocalls for mental health consultations (C), many did not have access to smartphones or computers, good internet connectivity, and a minimum technical expertise (M), which limited their ability to engage with services via video call.
- Ghosh et al. (2021) Among service providers of a substance disorder centre in India with a stepped telemental health model (C), in more than 16% of cases patients could not be contacted due to technical difficulties such as wrong numbers, line-busy, and non-response; video calls were often interrupted by the difference of bandwidths between the service users and providers, poor network connection, patients non-familiarity with the technology (M); in sum, the quality of interaction through video calls were was only modestly satisfactory (O).
- Hawke et al. (2021) Young people accessing mental health services virtually via video platforms (C) require smooth audio/video, with good Wi-Fi connection, technology and lighting (M) to make them feel comfortable engaging with therapy (O).
- He et al. (2020) When moving to remote care due to COVID-19 (C), if the software/program could not be connected with the remote system due to poor Wi-Fi reception (M), THEN this resulted in interruptions during online psychological assessments (O).
- Johnson et al. (2021) When moving to telemental health (C), if resources are inadequate (equipment and internet connections are of low quality, processes and preferred platforms are not clearly established, and/or staff lacks training and confidence) (M), then remote working works less well (O).
- Lakeman and Crighton (2021) Among service users with personality disorder receiving psychology / psychotherapy or counselling services in Australia when COVID-19 forced an abrupt cessation of their programmes (C), if/when there was a lack of technology and lack of data to hold a group video call (M), then this form of support was unsustainable for many clients (O).
- Lecomte et al. (2020) Young people accessing early intervention psychosis services living in remote areas or who have social anxiety (C) who have good internet connection and have access to technology (M) find remote sessions acceptable and will miss fewer sessions (O).
- Medalia et al. (2020) Among demographically homogenous, high socioeconomic status mixed secondary mental health service users using community mental health teams and outpatient services in NYC during a conversion to video supported telehealth due to COVID-19 (C), the

service users had access to high-speed internet and technology (M), and this supported the conversion to video-supported telehealth (O).

- The Mental Health Innovation Network (2020) For refugees who wanted to access telemental health support during COVID-19 (C), lack of access to technology or poor technological literacy (M) made implementation difficult (O).
- Ogueji et al. (2021) Among the general population in Nigeria during COVID-19 (C), as there is unstable and expensive electricity and internet across Nigeria (M), this would constitute a significant barrier to engaging in e-therapy (O).
- Simpson et al. (2001a) A telepsychiatry service was set up linking 5 local general hospitals in rural Canada to a more distant psychiatric hospital (C). At sites where there were proportionally more technical problems in the consultations (M), fewer consultations were carried out (O).

**Supporting information from stakeholder consultation:**

- Using phone calls is a reliable and commonly used option for telemental health and does not rely on internet connection.
- If there is NO connection, then face-to-face sessions are needed.

**Caveats and pitfalls identified by stakeholders:**

- There are whole communities (e.g. in rural areas) who do not have access to the internet, creating a postcode lottery.
- Difficulties accessing the internet may affect service providers as well as service users, especially both working at home and on service premises.
- Phone data gets used up more quickly during video meetings/consultations (such as those on Zoom or Teams) than Wi-Fi, which can result in people running out of data.
- Service users, carers and clinicians cannot be expected to solve the problem of lack of internet, or unstable internet connection, as it is largely out of their control; it is a problem that should be solved higher up (by Trusts, national government).
- Wi-Fi in wards across the NHS and in crisis houses is unreliable also, not just in private homes. Publicly available Wi-Fi may have security/firewall issues and therefore may not be appropriate. When staff were unsure of a patient's data plan, and there was unreliable Wi-Fi in crisis houses, then they resorted to using a phone call only.
- Expensive data/Wi-Fi is a barrier to telemental health.

#### CMO 1.3: Benefits of providing support and guidance for using technology

a. When staff who lack the confidence and/or knowledge to deliver mental health care online (particularly via video calls) receive practical instruction and guidance on how to use technology to deliver mental health services, including clear information about how to operate within local policies, procedures and platforms, troubleshoot issues during telemental health sessions, and formulate and implement back-up plans (C), they feel an increased sense of confidence in managing and delivering telemental health services (M), which leads to increased use of telemental health services (O1) and fewer delays, resulting in more appointments being completed on time (O2).

b. When service users with access to a technology device who struggle with the confidence, knowledge and/or ability to use telemental health receive guidance, reassurance and instruction (tailored to their healthcare provider and to their language, reading ability and any sensory disability) on how to use technology (particularly video calls) to access mental health care, engage with back up plans, and receive timely technical support and troubleshooting during treatment sessions (C), they feel an increased sense of confidence in accessing telemental health (M), which reduces anxiety using telemental health and digital technologies (including in their personal lives) (O1), facilitates adoption of and adherence to telemental health (O2), improves service users' ability to adjust to remote care (O3), reduces interruptions in care delivered via telemental health (O4) and increases satisfaction with telemental health (O5).

**Underlying CMOs:**

- Arighi et al. (2021) Among older adults seeking services for dementia (C), if they have support from young generation family/friends, e.g. digital natives (M), they are more likely to succeed in connecting and using telemedicine (O).
- Arighi et al. (2021) Among older adults seeking services for dementia during restrictions and lockdowns (C), if restrictions are lessened to allow them to have support from family/friends (M), they are more likely to succeed in connecting and using telemedicine (O).
- Castillo et al. (2020) Medical students working in virtual clinics for the treatment of opioid dependency (C) provided technological assistance to clients and the clinic, through creating online appointment request forms, promoting the clinic via social media and supporting clients to sign forms virtually and access videoconferencing appointments (M), which was instrumental in overcoming "technological barriers" (unspecified) that had limited the implementation of telemental health support for opioid dependency prior to the pandemic (O).
- Conn et al. (2013) Amongst clinicians at a telepsychiatry service for older adults in rural Canada (C), healthcare professionals reported that the use of educational seminars which included how to use telepsychiatry (M) made it possible for them to expand their repertoire of skills and improve patient care (e.g. by being able to provide them with more information) (O).
- Foye et al. (2020) Amongst service users in Scotland who were being assessed for or who may have had dementia (C), many did not have or did not understand the technology (M) and as such remote video appointments were deemed to be nearly impossible as reported by an older adult team in Scotland (O).
- Foye et al. (2020) Amongst mental health nurses working remotely during the COVID-19 pandemic to deliver care to service users who had cognitive impairments, were young, had a diagnosis of autism, or were experiencing psychosis and/or paranoia (C), they reported that service users often had a lack of understanding, access, or trust in the technology being used (M), and therefore found it difficult to deliver appointments and assessments.
- Ghosh et al. (2021) Among service users of a substance disorder centre in India with a stepped telemental health model who needed to access videocalls for mental health consultations (C), many did not have access to smartphones or computers, good internet connectivity, and a minimum technical expertise (M), which limited their ability to engage with services via video call.
- Godleski et al. (2012) Among veterans using a home telemental health messaging service (C), when an educational component was included in the messages sent to the patient (M), then patient satisfaction was high (O).
- Greenwood et al. (2004) Where an undersupply of psychiatric services in rural and regional Australia is likely to persist in the foreseeable future (C), if the use of telepsychiatry is used as an alternative in an informative, well communicated way that treats service users with respect (M), it can be used as a satisfactory means of service delivery (O).
- Humer et al. (2020) During the COVID-19 pandemic (C), if mental health experts received insufficient training (M), then this negatively influenced their practice (O).
- Johnson et al. (2021) When moving to telemental health (C), if service users are digitally excluded (lack equipment, resources and/or confidence to connect or live in overcrowded housing) (M), then remote working works less well (O).
- Johnson et al. (2021) When moving to telemental health (C), if staff experienced technological difficulties with remote appointments (40.3%), had to learn to use new technologies too quickly

and/or without sufficient training and support (40.8%), and had difficulties providing sufficient support via remote care (44.9%) (M), then this was perceived as a challenge for carrying out remote working (O).

- Johnson et al. (2021) When moving to telemental health (C), if resources are inadequate (equipment and internet connections are of low quality, processes and preferred platforms are not clearly established, and/or staff lack training and confidence) (M), then remote working works less well (O).
- Juarez-Reyes et al. (2021) Among study participants (people being treated in primary care for stress) who were transitioned to video sessions due to shelter in place orders (C), suggestions for onboarding patients onto the video sessions (such as a guide on accessing the Zoom platform on multiple devices, the available features, and practice time for logging on and using the various capabilities (M) might help reduce distractions and increase familiarity with using the videoconferencing platform (O).
- Kanellopoulos et al. (2021) When a psychiatric inpatient service provided verbal and written instructions on how to use electronic tablets to inpatients who were isolated in their room due to COVID-19 and had limited ability to navigate technology due to psychopathology, cognitive impairment, or limited prior familiarity with technology (C), then this decreased barriers to telemental health use (O), as patients were able to gain necessary skills and knowledge to use telemental health technology (M).
- Lakeman and Crighton (2021) Among staff working in psychology / psychotherapy or counselling services for people with personality disorder in Australia when COVID-19 forced an abrupt cessation of their programmes (C), if/when there was a lack of experience of delivering programmes online among clinicians (M), then this led to a lack of confidence delivering programmes online (with 32% of clinicians reporting being 'not at all' confident) (O).
- Lin et al. (2020) Among psychological hotlines for the general population (C), when there was the provision of good supervision and training resources (M), then this differentiated between whether a hotline was a good hotline or needed improvement (O).
- Mental Health Commission of Canada (2014) In order to reduce attrition from online mental health interventions/treatments (C), an "e-case manager" role was created, whose job it was to coach people through online interventions or treatments (M), then there was evidence that this decreased dropouts and loss to follow up (O).
- The Mental Health Innovation Network (2020) For refugees who wanted to access telemental health support during Covid-19 (C), lack of access to technology or poor technological literacy (M), made implementation difficult (O).
- Mental Health Network NHS Confederation (2020b) When Cambridgeshire and Peterborough NHS Foundation Trust was aiming to make staff feel confident in providing digital services (C), brief guidelines were developed for staff which were used with all community teams and referrers to the service (M), which gave staff the confidence and permission to make informed decisions around digital referrals and where they were appropriate or not appropriate within their pathway (O).
- Open Excellence (2020) Among Hearing Voices Group facilitators in the US who could not facilitate in-person groups during the COVID-19 pandemic (C), when experienced online facilitators were funded to provide support and practical instruction to others about moving online (M), an increased number of Hearing Voices Groups were able to move online and provide ongoing support during the pandemic (O).
- Pote et al. (2021) Trainee clinical doctoral psychologists (C) need a strong and responsive IT infrastructure (e.g. ideally working collaboratively with IT experts to create workable solutions) (M) to help them feel comfortable engaging in providing digital mental health services (O).
- Pote et al. (2021) Trainee clinical doctoral psychologists (C) who receive very little guidance, training or experience of delivering digital mental health care during their training (M) feel fearful and reluctant to engage with this mode of delivery (O) / trainee clinical doctoral

psychologists (C) who receive support, guidance and experience of digital mental health during their training (M) will be more prepared to deliver this care in practice (O).

- Rabinowitz et al. (2010) Among older people in nursing homes receiving telepsychiatry via either video of telephone who experienced technical problems during their session (C), if the clinician was able to correct the issue quickly enough (M), then the appointment could be completed during the scheduled slot (O).
- Vera San Juan et al. (2021) Among older mental health service users receiving remote care during the COVID-19 pandemic (C), when they did not receive support or information to help them engage with remote care, or options to use services they were familiar with (M), then people were not as able to adjust to remote care (O).
- Scharff et al. (2020) Among inexperienced trainees who had to handle various challenges and learning opportunities due to remote supervision (C) if they had a strong supervisory relationship due to being supervised by faculty members of their doctoral programs (M) then this aided with challenges related to navigating changes in supervision (O).
- Shore et al. (2014) Among veterans enrolled in Home-Based Telemental Health video sessions experiencing technical issues (C), the presence of peer support technicians who could troubleshoot technology issues (M) led to more successful service delivery (O).
- Shore et al. (2014) Among veterans were enrolled in Home-Based Telemental Health video sessions who had never previously used video technology (C), they gained competency using technology and making video calls through the session (M) which led to veterans using their new skills to connect via Skype to see their grandchildren for the first time, connect with former platoon members or other important people in their past (O).
- Shore et al. (2014) When there was overlapping facility and peer technical support for veterans engaging in Home-Based Telemental Health video sessions (C), at times there were multiple technical support personnel troubleshooting one case and no communication between them (M), leading to confusion among providers (O).
- Williams et al. (2017) Among homeless individuals asked about what was needed in relation to the kind of support that might be required for people to access digital technology for health purposes (C), if there was more one-to-one support and digital training (M), then this might improve confidence and lessen anxiety around using the internet for health purposes (O).
- Yellowlees et al. (2020) Among clinicians who had to get accustomed to using their own computers and home setups when moving to remote working (C), if they received training and supervision and IT support to assist in the conversion process (M), then this reduced technology barriers for clinicians (O).

**Supporting information from stakeholder consultation:**

- Training also needs to include confidence building.
- Familiarity and confidence are key for service users and practitioners, hence helpfulness of using platforms that are already familiar.
- People googled user guides for unfamiliar platforms.
- Training people to use technology has wider benefits for their personal goals relevant to recovery.
- Showing people how to do things is important – especially when literacy levels may be low and written instructions might not help – and this training might need to be repeated if people have poor retention.
- New digital spaces are being established that include peer spaces where young people can interact with and learn from each other – this is happening more and more, so we need to understand it and adopt it whether we want to or not. We may need to moderate these conversations. There are risks and it is important to manage them.
- There is an assumption that staff are competent and confident with telemental health and have the equipment – this is not necessarily true.

- Staff have the added stress of making connections with unfamiliar technology, and the clinicians are meant to do technical support during sessions, and this itself can affect the therapeutic relationship – it is not a safe space, as it can be full of uncertainty and difficulty.
- For staff working remotely, we make assumptions that staff have things that they do not necessarily have (devices, connection, skills etc). We are changing the way staff are working, including a lack of the peer support that comes with working in a hard situation. If a risk situation comes up, there are always people around to call but it does not feel the same in terms of team connection, support and thinking.

**Caveats and pitfalls identified by stakeholders:**

- There are uncertainties over working remotely or over the phone with young children, as telemental health platforms are usually appropriate for older children, but younger children (and their parents) will need training, or at very young ages may not be able to engage with technologies.
- Telemental health training might not be accessible for service users who speak using sign language or a different language – tailored materials need to be provided. Training and guidance should include training on spam and malware as these may disproportionally target and affect vulnerable people. Training needs to ensure it covers safeguarding when using telemental health.
- Time between training and actual use is important to consider – a long time between them might reduce the effectiveness of the training.
- It may be difficult for service users to learn new skills, or use technology, devices and screens at all when they are unwell, for example those with psychosis.

#### CMO 1.4: Impact of technology related disruptions

When technological issues (including connection problems and device issues) lead to disruptions to online sessions and there is no pre-arranged back-up method of contact (e.g. a plan to connect by telephone instead of video-call if needed) (C), the quality of the intervention is diminished (O1) there is a loss of empathic connection between client and therapist (O2) and the sessions may not be able to continue (O3), as the flow of the conversation is interrupted and session time reduced, for example, when having to ask the other person to repeat what has been said, or when cut off completely, leaving staff and service users potentially feeling distracted, frustrated, awkward and upset (particularly if there is a threat of therapy withdrawal due to missed sessions) (M).

**Underlying CMOs:**

- Fogler et al. (2020) Moving a parents therapy group (for parents of children with ADHD) online due to the Covid-19 pandemic (C) resulted in some technical challenges, such as not being able to see all group members at the same time (M), which negatively impacted the flow of the conversation (O).
- He et al. (2020) When moving to remote care due to COVID-19 (C), if the methods of intervention, such as hotline and online videos, were relatively dull, and the signal transmission might be delayed (M), then the quality of psychological interactions was not as high as compared with face-to-face interactions, which might reduce the effect of the interventions (O).
- Healthwatch (2021) Among young people who experience connection issues during sessions (such as less than optimal internet) (C), they miss what the other person is saying and need to ask them to repeat themselves (M), which leads to frustration and awkwardness for the young person (O).

- Lichstein et al. (2013) Among older adult mental health service users who were receiving remote CBT via Skype for insomnia and depression (C), the audio and video were occasionally offset during sessions (M), which resulted in the service user feeling distracted (O).
- Lodder et al. (2020) When there were technological issues, such as poor sound quality, during a Zoom online intervention for parents of children with autism (C), this made the conversation feel less natural (O), as participants found it difficult to hear everything (M).
- McBeath et al. (2020) Among psychotherapists delivering online psychotherapy to mixed psychology service users during COVID-19 (C), if/when there were connection problems and time lags between visual images and sound (M), then this negatively impacted on the psychotherapists' ability to engage with their clients (O).
- Mind (2021a) For people engaged in online therapy (C), technological mishaps meaning missed sessions lead to the threat of therapy withdrawal (M), causing upset to the patient (O).
- Pugh et al. (2020) When chairwork, such as empty-chair dialogues and role-play, is moved online so service users took part at home (C), therapists reported that technical barriers, such as a poor internet connection (M), resulted in a loss of empathic connection between the client and therapist (O).
- Severe et al. (2020) Among mixed secondary mental health service users receiving remote care (C), if service users had unstable internet connection or reported video platforms to be otherwise problematic (M), then they thought that telephone visits were particularly advantageous (O).
- Uscher-Pines et al. (2020b) When switching to remote care during the pandemic (C), if patients had difficulties with technology (M), then this impacted the flow of visits and caused frustration (O).

**Support for CMO from stakeholder consultation:**

- None identified.

**Caveats and Pitfalls identified by stakeholders:**

- If the interruptions cannot be overcome, then face-to-face appointments may be needed.

#### CMO 1.5: Preparing service users for telemental health

When staff prepare service users for telemental health appointments and communicate clearly with service users about what to expect (C), this leads to more accepted calls and fewer missed service user contacts (O), because service users have more relevant knowledge of the process, including when to expect contacts and from what number, and feel more comfortable engaging with telemental health services (M).

**Underlying CMOs:**

- Aafjes-van Doorn et al. (2020) Staff who had to switch to remote working during the COVID-19 pandemic (C) who used a greater variety of methods to prepare their patients for the switch (M) reported higher levels of online alliance (O).
- Newbronner et al. (2021) For a service user with SMI user accessing remote therapy (telephone) during COVID-19 (C), appointments taking place later than scheduled) (M), resulted in potential conflict with other scheduled appointments (O).
- O'Dell et al. (2021) Among patients who were reluctant to answer telephone calls from their therapists who were using unfamiliar or blocked phone numbers (C), when the team trialed several platforms to route calls through local clinic numbers (M), then this led to increased responsiveness (O).

- O'Dell et al. (2021) Among behavioural health providers using a programme called Doximity (C), as this programme allowed calls to be sent from a familiar clinic number (M1) which prompted patients to return calls through standard hospital channels (M2), then this reduced missed patient contacts (O).

**Supporting information from stakeholder consultation:**

- Clear expectations and transparency of using telemental health are important.

**Caveats and pitfalls identified by stakeholders:**

- None identified.

#### CMO 1.6 Familiarity with platform and ease of use

Where service users already use remote technologies for social, educational or work purposes, and/or where the online platforms are relatively easy to use (C), offering a choice of familiar and accessible technology platforms that may be less difficult and time-consuming for staff and service users to understand or learn (M) may increase the likelihood of engagement with services via telemental health, especially video calls (O).

**Underlying CMOs:**

- Barney et al. (2020) Staff in community mental health teams in the USA who had previous experience with electronic communication (C) found telemedicine acceptable and convenient (O) as it was familiar to them (M).
- Connolly et al. (2021) Among the Veterans Affairs service which provides community and outpatient mental health support (C), as they had previously used and had widely implemented telemental health via videoconferencing (M), then they were able to rapidly increase capacity for online appointments due to COVID-19 (O).
- Ghaneirad et al. (2021) When offering video consultations to service users due to the pandemic (C), service users were more likely to take up video consultations (O), if they were younger and had higher educational levels (number of previous sessions, diagnoses, or language proficiency did not influence the uptake) (M).
- Hensel et al. (2021) Among postpartum women undergoing therapy for depression and/or anxiety in Canada (C), if those women were familiar with video technology such as FaceTime or Skype from their personal or professional lives (M), then they were more willing to try video therapy (O).
- Hopkins and Pedwell (2021) Young people who have already engaged with services (C) have a prior connection and relationship with providers (M), therefore making engagement and using telehealth easier for providers (O).
- Humer et al. (2020) During the first weeks of COVID-19 lockdown (C), if therapists already treated patients via the Internet (M), then they felt more prepared for the use of Internet in psychotherapy than those who didn't treat patients via the Internet (O).
- Peralta and Taveras (2020) Among minors seeking mental health support during the COVID-19 crisis in the Dominican Republic (C), if the teleconsultation platform was easy to use (M), then this allowed minors to seek timely help (O).

**Supporting information from stakeholder consultation:**

- Ease of use may arise through familiarity, but a programme could also be unfamiliar and easy to use.

- Lots of service users access platforms via their phones – familiarity with the format (e.g. video, phone calls via that platform) is important as well.

**Caveats and pitfalls identified by stakeholders:**

- Trusts need to be able to critically evaluate the safety of the tools that are on the market. Introducing technology can be hard and dependent on how ready trusts are to use a variety of digital tools and do that safely.
- Flexibility and providing support for a range of platforms has implications for service providers, including for workload, training, IOT support and resources, governance, etc.
- Members of the public, especially children and young people might not be familiar with platforms like Teams. Services tend to ask users to use the technology which the service has access to, rather than using technology that is familiar to the service user.
- Staff are sometimes unsure about which platforms they are allowed to use, and sometimes red tape about what can or cannot be used stands in the way, with considerable inconsistency between providers in what is acceptable. Communication and decision-making responsibilities need to be clear.
- There are sometimes mixed messages about what platforms people can use and clinicians are unsure if they can challenge that decision, e.g. if a platform is banned which leads to negative impact on service provision. Zoom appears to be a contested issue – popular because of its ease of use and familiarity to many of the public, but sometimes restricted – stakeholders suggested that this platform should be acceptable and has substantial advantages provided safety features are used and appropriate training provided.

#### CMO 1.7: Telemental health may be a better alternative to receiving no care during an emergency, such as the COVID-19 pandemic

When service users are offered telemental health appointments because face-to-face appointments are restricted (e.g. due to COVID-19 or another emergency) (C), these appointments are likely to be accepted by some service users on the basis that they are the main way by which mental health care can continue (O1) and there is a reduced risk of infection from COVID-19 (O2), as face-to-face options are lacking or very restricted and telemental health is seen as preferable to receiving no support at all and as an alternative to cancelling appointments entirely (M).

**Underlying CMOs:**

- Hensel et al. (2021) Among postpartum women undergoing therapy for depression and/or anxiety in Canada (C), when technical difficulties which occurred were minor and infrequent and barriers to attending face-to-face therapy were significant (M), then service users were willing to tolerate technical difficulties as a lesser evil (O).
- Kanellopoulos et al. (2021) When a psychiatric inpatient service provided electronic tablets to inpatients who were isolated in their room during a rapid switch to telemental health due to COVID-19 (C), if each night the service collected tablets from patients, disinfected, charged, and returned the tablets the following morning (M), then this secured patients’ physical safety (O).
- Liberati et al. (2021) Some users of secondary mental health care who were offered remote care during the pandemic (C), if they thought face-to-face consultations would be reintroduced soon (M), chose not to receive remote care (O).
- Oqueji (2021) Among the general population in Nigeria during COVID-19 (C), when health practitioners did not offer face-to-face appointments during COVID-19 due to not wanting to

| |
|---|
| have close contact with patients (M), then there was increased willingness to attend therapy via telemental health (O).<br>• Sehlo et al. (2021) Among service users of an online therapy site in Egypt during covid-19 (C), online therapy was reported as helpful during the pandemic (O), as it avoided COVID-19 infection (M).<br>• Severe et al. (2020) Among mixed secondary mental health service users, especially those with underlying medical conditions, during the COVID-19 pandemic (C), if they thought that virtual visits would reduce their ability to contract COVID-19 (M), then this was associated with an increased likelihood of wanting to receive telemental health care in the future (O). |
| **Supporting information from stakeholder consultation:**<br>• None identified. |
| **Caveats and pitfalls identified by stakeholders:**<br>• This CMO was relevant in the context of the COVID-19, but may have diminished in importance with the reinstatement of more face-to-face care as the pandemic progressed. Mask-wearing requirements may also have influenced preferences for telemental health vs. Face-to-face care.<br>• This is also relevant for staff who may be shielding or in isolation during COVID-19: staff were able to give appointments and/or continue training even when isolating, meaning the patients could continue to receive services. |
| **Additional sources utilised**:<br>• Kanellopoulos D, Castellano CB, McGlynn L, Gerber S, Francois D, Rosenblum L, et al. Implementation of telehealth services for inpatient psychiatric Covid-19 positive patients: A blueprint for adapting the milieu. General Hospital Psychiatry. 2021;68:113-4.<br>• Ogueji IA, Amusa AO, Olofe OJ, Omotoso EB. Willingness and barriers to utilizing e-therapy services: A Nigerian general population qualitative study. Journal of Human Behavior in the Social Environment. 2021;32(2):214-28.<br>• Sehlo MG, Youssef UM, Elshami MI, Elrafey DS, Elgohari HM. Telepsychiatry versus face to face consultation in COVID-19 Era from the patients' perspective. Asian Journal of Psychiatry. 2021;59:102641.<br>• Severe J, Tang R, Horbatch F, Onishchenko R, Naini V, Blazek MC. Factors influencing patients' initial decisions regarding telepsychiatry participation during the COVID-19 pandemic: Telephone-based survey. JMIR Formative Research. 2020;4(12):e25469. |

### Domain 2. Being Flexible and Personalised

#### CMO 2.1: Taking individual preferences into account – offering alternatives

When services using remote mental health care allow service users to choose the modality of telemental health and/or a choice of remote versus face-to-face care, and regularly check their preferences (C), this allows service users to have greater autonomy and choice (M) leading to them feeling more satisfied with and able to engage with the type of care received (O1), leading to improved uptake (O2), and improved therapeutic relationships with their clinician (O3).

**Underlying CMOs:**

- Adamou et al. (2021) When service users with Autism and ADHD (C) felt that they had sufficient time and space to communicate clearly and explain themselves (M) they feel more positively about the use of remote technology (O).
- Schueller et al. (2019) In a tailored digital mental health intervention using text messaging as an adjunct to group CBT (C), because they provided the service was provided to Latinx service users in Spanish (M), then this resulted in significantly increased engagement, as evidenced by patients attending more sessions and staying in treatment longer (O).
- The British Psychological Society (2020) For staff providing remote counselling (C) they should assess the service users preferences/ needs/ access and suitability to using technology (M) to ensure access and engagement in this type of modality (O).
- Costa et al. (2021) During the Covid-19 pandemic in the USA (C), some of those mental health service users who said that they were coping well during the pandemic (O), reported meeting with their providers over the telephone but wanted the option to choose whether to have phone sessions or in person sessions after the restrictions were lifted (M).
- Eagle (2020) Amongst service users using video platforms for therapy (e.g. zoom) (C) they may feel more self-conscious (M) therefore disengage or cancel sessions (O).
- Greenwood et al. (2004) Where there is limited provision for psychiatry in rural and remote locations and telepsychiatry services are offered (C), some service users report not feeling comfortable in front of the camera (30%) (M) which may lead to lower levels of patient satisfaction (O).
- Guinart et al. (2020) Among patients receiving telepsychiatry during COVID-19 (C), some reported that they felt more comfortable at home and could express themselves more freely (M), and thus suggested that remote assessments should be maintained (O).
- Healthwatch et al. (2021) Among young people feeling pressure to have their camera on during a video call (C), if the professionals clarify that it is okay for them to type in the chat rather than speak on camera (M), then this can reduce the young person's nervousness around the session (O).
- Hensel et al. (2021) Among postpartum women undergoing therapy for depression and/or anxiety in Canada (C), when they had the option of video therapy but chose to attend in person (M), video therapy was felt to be an acceptable backup option in case attending became too difficult (O).
- Juarez-Reyes et al. (2021) Among study participants (people seen in primary care for "stress") who were transitioned to video sessions due to shelter in place orders (C) when given the choice about video sessions (M) participants' preferences varied with some appreciating the convenience and comfort of video sessions, some preferring face-to-face, and others open to either format (O).
- King et al. (2006) Young people engaging with text-based counselling (C) are able to take more time to reply - thus feel more in control - and cannot be overheard by family in their house as it is typed (M) and therefore, they feel more comfortable and engage more (O).

- Liberati et al. (2021) Service users who had been using secondary mental health services, and staff, when services initially moved to remote because of COVID-19 (C) if only phone calls were available (M) both service users and staff felt frustrated at the lack of choice in how to stay in contact with services (O).
- Liberati et al. (2021) When services were transferring to remote care because of COVID (C) taking a 'one-size-fits-all' approach to remote care (M) excludes some people, including people who choose to withdraw from care (O).
- Liberati et al. (2021) In the move to remote care because of COVID-19 (C) if staff recognised the challenges in providing remote care, understood service user choice and offered face-to-face care once when allowed (M) then some service users wanted to resume face-to-face care (O).
- Liberati et al. (2021) Among staff and service users who had been using secondary care mental health services before COVID when services moved to remote care (C) if they were offered limited influence or choice about how care was provided (M) then services couldn't provide tailored or personalised services (O).
- Mind (2021b) Young people accessing mental health support services remotely (C) felt that greater flexibility about when they could access remote support (M) made engagement easier (O).
- National Health Service England (accessed 2022) Among crisis line handlers who do not speak the same language/preferred language as the caller (C), as the crisis handler had the option to contact the Language shop in Newham where someone who speaks the required language was identified (M), then this allowed for the call hander to call the person in crisis back and has a three-way conversation (O).
- Nissling et al. (2020) Among people undergoing primary care internet-mediated CBT in Sweden (C) when peer support was provided via phone calls and asynchronous text messages (M) this enabled service users to access treatment when it suited them and reflect on content at their own pace (O).
- Nissling et al. (2020) Among people undergoing primary care internet-mediated CBT in Sweden, with peer support provided via phone calls and asynchronous text messages (C), when the digital nature of the contact removed pressure normally felt in social settings (M) then it was easier to open up to the peer support worker (O).
- Orlowski (2016) For staff working in youth mental health services before the pandemic (C) a willingness and curiosity to seek clarification and learn from the young person (in contrast to fluency in current tech language) (M) was useful to the relationship (C).
- Orlowski (2016) Some staff working in youth mental health services before the pandemic, with sufficient resources (phone tablet or laptop and internet connection) (C) used technology with consumers (M) to increase engagement and develop rapport (O).
- Schueller et al. (2019) In a tailored digital mental health intervention using text messaging as an adjunct to group CBT (C), as they provided this service to Latinx service users in Spanish (M), then this resulted in significantly increased engagement, as evidenced by patients attending more sessions and staying in treatment longer (O).
- Schueller et al. (2019) A depression intervention called SPARX (C), as it contained elements that made the services more appropriate for the LGBTQ population struggling with symptoms of depression (for example  the rainbow version includes multiple culturally sensitive script changes to the intervention modules, avatars that can be customized without strict gender norms, and content addressing specific challenges among this population (e.g., coming out, getting bullied) (M), then participants strongly endorsed the effectiveness and acceptability of Rainbow SPARX for treating their depression and it significantly reduced symptoms of depression and anxiety (even after 3 month follow up) (O).
- Severe et al. (2020) Among paediatric patients receiving telemental health (C) if the children had difficulties focusing and building a therapeutic relationship using telemental health while using

video technology (M) then this decreased the likelihood of wanting to receive telemental health in the future (O).

- Shklarski et al. (2021) Clinicians who conducted psychotherapy with children over Zoom (C) found that this modality made it harder to engage them in therapy (when compared to face-to-face) as it was difficult to do play or art therapy (M), which negatively impacted the therapeutic relationship (O).
- Simon et al. (2021) NHS commissioners and managers in charge of implementing telemental health initiatives felt that amongst patients (C) convenience of accessing treatments at own pace and having access post the treatment period (M) would be of value to service users (O).
- Vera San Juan et al. (2021) For some mental health service users utilising telemental health during COVID-19 (C), phone calls felt less intrusive than video calls (M) and then phone calls were therefore the preferred method of receiving telemental health support (O).
- Vera San Juan et al. (2021) Among mental health service users who had access to remote care during the COVID-19 pandemic (C), when service providers were inflexible (such as regarding the type of technology used or did not offer a choice regarding the delivery mode of care) (M), then service provision was impeded and perceived as inequitable (O).
- Vera San Juan et al. (2021) Among mental health service users relying on services other than secondary mental health services, such as their GP, charities, crisis helplines and/or online support groups during the COVID-19 pandemic (C), as they offered new forms of remote contact that were seen as broadening choice and service availability (M), then participants reported receiving high quality care, information and resources (O).
- Watson et al. (2021) Among service users with psychosis accessing psychological therapies before COVID-19 (C), if they preferred in-person therapy (M), then they tended to decline remote therapy (O).
- Yellowlees et al. (2020) For patients who could not/did not want to videoconference, for example because they were elderly and/or did not have the necessary technology (C) if the service remained flexible and arranged phone calls while encouraging video calls (M) then patients felt more at ease, were willing to try virtual visits (if possible) and technology-related appointment cancellations were avoided (O).
- YoungMinds (2020) Among young people accessing Children and Adolescent Mental Health Services support (C) where they do not have a choice in the mode of delivery (e.g. face-to-face or digital) (M) there is lower engagement and higher dropout rates (O).

**Supporting information from stakeholder consultation:**

- Telemental health will not be appropriate for everyone, although could be used where appropriate. Some people still need a personal face-to-face connection, or others need support in place to help people use telemental health.
- Clinicians might find it easier to build rapport face-to-face or using videoconferencing vs telephone, but for some service users, telephone technology might be preferable for building a therapeutic relationship (e.g. due to sensory overload). Some service users may also feel less self-conscious over the telephone than using video.
- Giving service users choice is vital rather than imposing telemental health (links to power imbalances). Services need to make people feel more empowered and to feel in control. Highlights the importance of shared decision making about whether to use telemental health and how it is used.
- Every service needs to make sure they are using the best platforms for their service users regardless of costs to ensure patient safety, however sometimes this does not match up with patient preference. This can risk digital exclusion because some service users do not want to learn how to use a new platform.
- For autistic adolescents, doing session virtually can be easier than having to be in a room with people, e.g. they can choose to turn their camera off and type in the chat, rather than speak. For autistic young people who have special interests in video games, for example, having a digital platform makes it easier because it is closer to their special interests.
- Texting is central to how young people communicate. If there is an existing relationship and the clinician sends a text on an important day to the young person wishing them well then this is really valued by young people. Texting also has the advantage of being instant, young people are more likely to reply via text than pick up a phone call. Texting can also be used to prepare young people for a phone call, especially before the first contact – they may be unlikely to answer calls from unknown numbers. It also allows healthcare to be delivered on their terms, again showing importance of shared decision making.

**Caveats and pitfalls identified by stakeholders:**

- Individual characteristics and difficulties may prevent some people engaging with telemental health at all, e.g. service users with social anxiety and paranoid ideation or those who do not feel confident in the platform.
- Individual preference might be related to their previous experiences of face-to-face versus online care. Choice should always be informed based on an explanation of different platforms.
- If people are in their homes, their domestic situation may compromise the quality of the therapy: practitioners need to be aware and check this out at the beginning.
- Stakeholders reported examples of people being told: "You can have a telehealth appointment today, and an in-person appointment in 6 weeks"'– there needs to be equality of care across formats.
- Giving service users the choice over platforms can be difficult as clinicians are often restricted by policies of the service e.g. only having a licence for one videoconference platform, or only allowing one platform due to security concerns. Some services could only offer Attend Anywhere, whereas service users were used to using Teams or Zoom professionally or personally, so would feel more comfortable with one of these platforms.
- Service users should also have the choice not to see themselves on the screen (whilst allowing staff to still see them). Some children and young people were bothered about their appearance (e.g. because of self-harming, having an eating disorder etc.). Zoom has this option, however, not on all devices and not everyone knows how to use it. Whilst doing this, there is a need to balance autonomy (of turning off the camera) and handling risk.

- In inpatient units, there is a need to invest in proper equipment to allow for care to be provided remotely. For inpatients, negative experiences are often related to quality of technology and connection when having remote therapy or talking to family and friends via video call rather than quality of the therapy or the clinician.
- Choice and power are often taken away from service users in crisis: services need to offer as much as choice as they possibly can.

#### CMO 2.2: Removing barriers – greater convenience for users and family/friends

Among some service users and family and other supporters experiencing specific practical barriers to attending face-to-face services (childcare or other caring responsibilities, location, work, mobility limitations travel difficulties/costs, work commitments) and who have good access to telemental health (C), telemental health may provide increased flexibility that addresses individual practical barriers (M), which can lead to telemental health being viewed by some service users and carers as more convenient and accessible than face-to-face care (O1), easing attendance (O2), increasing uptake and reducing missed appointments (O3).

**Underlying CMOs:**

- Chen et al. (2020) Patients with psychiatric pathologies that interfere with their ability to leave home (e.g. obsessive-compulsive rituals) attending general psychiatry service in the USA (C) were able to access care more consistently due to the shift to telemental health (M), which further resulted in decreased no show rates (O).
- Choi et al. (2014) When older adults (many of whom lacked transportation) received problem solving therapy using video conferencing software rather than face-to-face care (C), they found telehealth acceptable (O) as they liked the increased convenience and additional privacy of accessing therapy at home (M).
- Fogler et al. (2020) Moving group therapy for parents of children with ADHD online due to the COVID-19 pandemic (C) led to increased flexibility for parents (M), which improved attendance (being remote meant that both parents could attend as one didn't have to stay at home with the children/didn't have to get childcare) (O).
- Fogler et al. (2020) Moving group therapy for parents of children with ADHD online due to the COVID-19 pandemic (C) led to increased convenience for parents and cost savings due to not having to travel (M), which led to this modality being positively evaluated by parents (O).
- Frayn et al. (2021) People experiencing binge eating spectrum disorders (BESD) during the pandemic using virtual therapy (C) felt reductions in practical barriers to attending appointments, e.g., finding car parking, reduced anxieties related to being on time for an appointment (M) so reduced overall stress of attending appointments (M), which made them more convenient and easier to attend (O).
- Frayn et al. (2021) People experiencing Binge Eating Disorder using teletherapy during the pandemic (C) felt that increased flexibility of appointments made them easier to fit into their day (M), making them more convenient and easier to attend (O).
- Frayn et al. (2021) For some people experiencing binge eating spectrum disorders during the pandemic (C), the increased convenience of accessing therapy provided virtually (M) increased motivation to engage with support (O).
- Gaddy et al. (2020) For service users who were either in remote locations or who were among the very sick or immunocompromised (C), telehealth was a means for service users to access services during the COVID-19 pandemic (O), as it avoided infection control-related risks (M).

- Ghosh et al. (2021) Among service users of a substance disorder centre in India with a stepped telemental health model (C), follow-up care was easier to deliver (O) through telemedicine as a majority required only telephone consultation (M).
- Guinart et al. (2020) Among patients receiving telepsychiatry during COVID-19 (C), as patients could request less time off work (M), then they suggested that remote assessments should be maintained (O).
- Guinart et al. (2020) Among patients receiving telepsychiatry during COVID-19 (C), as they could save transportation time and costs for patients (M), then patients suggested that remote assessments should be maintained (O).
- Hawke et al. (2021) Young people accessing mental health services (C) who are able to easily book sessions and where services are accessible and convenient, and there is a lack of travel (M) will engage well with virtual services (O).
- The Health Foundation (2020) Among young people who live in remote areas who are attending Child and Adolescent Mental Health Service (CAMHS) appointments alongside school (C), using video calls saves time and avoids long travel therefore saves money, the need for parents to take time off work to drive them, and avoids the young person taking time off school (M) therefore they are more likely to attend their appointments (O).
- Hensel et al. (2021) Among postpartum women undergoing therapy for depression and/or anxiety in Canada (C), if those women faced additional barriers in accessing face-to-face therapy such as difficulty travelling with a new-born or arranging childcare for older children (M) they were more willing to take up the offer of video therapy (O).
- Hernandez-Tejada et al. (2014) Veterans receiving exposure therapy for PTSD (C) who face external pressures or costs such as parking, fatigue after work or childcare issues (M) are likely to miss more sessions/ drop out of treatment earlier than those receiving telemedicine delivery (O).
- Johnson et al. (2021) Among service users who find traveling and public places challenging (C) if remote care allows to receive treatment from home (M) then this improves access to care (O).
- Juarez-Reyes et al. (2021) Among study participants (people in primary care seeking help for stress) who were transitioned to video sessions due to shelter in place orders (C) if the participants found the video sessions convenient, eliminating travel time and reducing some of the stresses of in-person contact (M) then the participants preferred the video format over meeting in person (O).
- Liberati et al. (2021) During the pandemic, when services were provided remotely, service users who were shielding or who had logistical or financial travel difficulties (C) could continue to access services (M) so that some inequalities of access were reduced (O).
- Lodder et al. (2020) When delivering a Zoom online intervention to parents of children with autism (C) this increased the likelihood and possibility of participants' attendance (O) as parents did not have to arrange childcare (M).
- Mind (2021a) For people from both a minority ethnic and white background (C) not having to travel (M) made phone and online support easier to take up than face-to-face care (O).
- Mind (2021b) Young people accessing mental health support services remotely (C) felt that greater flexibility on when they could access remote support (M) made engagement easier (O).
- Newbronner et al. (2021) Among some service users with severe mental illness (Psychosis, bipolar and other severe mental illness undefined) accessing remote therapy during COVID-19 (C), some found telephone and online support more convenient and less stressful (i.e. didn't involve travelling or leaving pets alone) (M) and therefore preferred telephone and online forms of remote therapy (O).
- Open Excellence (2020) Among voice-hearers in the USA without access to in-person peer support from hearing voices groups (C), when online groups are available (M), this facilitates access to a novel kind of peer support in addition to any other help available (O).

- Pugh et al. (2020) When chairwork, such as empty-chair dialogues and role-play, is moved online so service users took part at home (C), service users were not limited by geographic or health related barriers (M) which made therapy more accessible (O).
- Sehlo et al. (2021) Among service users of an online therapy site in Egypt during Covid-19 (C), online therapy was found advantageous (O) because it saved money and time for travel and waiting time at the clinic (M).
- Severe et al. (2020) Among a mixed group of secondary mental health service users (C) if they thought that virtual visits were more convenient, and that video technology was easy to use and associated with greater provider availability (M) then they were more likely to want to receive telemental health in the future (O).
- Shore et al. (2014) Among veterans enrolled in Home-Based Telemental Health video sessions (C), convenience of being seen in the home was universally acknowledged by veterans (M), and they strongly agreed that they would rather use telehealth than travel to see the provider (O).
- Sheehan et al. (2020) Among clients who struggle to attend services for counselling (C) if access is improved through telephone or online counselling (M) then this leads to a decrease in rate of non-attendance (O).
- Simpson et al. (2001b) Among patients in rural Canada using a routine telepsychiatry service linking 5 local general hospitals with a specialist psychiatric hospital Cc), when most patients (23/31) saved money due to not having to travel up to 200km to a specialist service, take extra time off work, or find childcare (M), most patients (25/31) preferred telepsychiatry to travelling for an in-person appointment (O).
- Uscher-Pines et al. (2020a) AMONG certain underserved patients who did not have access to care (C) if moving to remote care solved logistical challenges that made it difficult to access care before (M) then this increased access for these patients (O).
- Uscher-Pines et al. (2020b) When remote care was provided during the pandemic for patients with opioid disorder who lack access or for whom it is inconvenient or burdensome to travel to appointments (C) as did not have to travel to appointments (M) this increased access and convenience and thus reduced the no show rate (O).
- Uscher-Pines et al. (2020b) When remote care was provided during the pandemic for patients with opioid disorders (C) if remote care was experienced as more convenient than face-to-face (M) then patients were more likely to want to continue to use remote care after COVID-19 (C).
- Wilson et al. (2021) When a switch was made to remote care for busy new mothers (C) if virtual appointments provided greater flexibility (M) then mothers experienced remote consulting to be beneficial (O).
- Yellowlees et al. (2020) Among patients with children who worked from home and for whom it was increasingly risky to bring children to the health care setting or difficult to arrange childcare due to COVID-19 (C) if they did not have to worry about childcare because they received telemental health care (M) then patients responded positively to virtual consultations (O).
- Vera San Juan et al. (2021) Among mental health service users who switched to remote care during COVID-19 (C), as it facilitated access for those who would otherwise not have been able to receive some care (such as those admitted to mental health wards or participants living in remote areas that have only limited services available) (M), resulting in successful service delivery to some excluded groups (O).

**Supporting information from stakeholder consultation:**

- Where parents have multiple children and may also be accessing mental health services themselves, telemental health has made it more convenient and reduced some financial barriers. Parents with young children also do not need to worry about childcare which makes a huge difference to accessibility of mental health support, and if their child is ill they can still engage with a programme online.

- Young people can be anxious about meeting new people, telemental health may make made it less intimidating to meet a new person.
- Remote care has been great for people with mobility problems therefore reducing their exclusion from care.
- The pros and cons of face-to-face care when people are wearing masks (due to the pandemic) vs. telemental health may be different from the choice between in person contact without masks and telemental health. Service users and clinicians might miss more cues and subtleties of interactions when masked versus using telemental health, and mask wearing might make contacts seem more impersonal.
- In inpatient care, telemental health can be positively used to bridge the transition between inpatient/crisis care and community care. Often people struggle when going back to the community due to a lack of activities (compared to inpatient care). This is especially the case in rural areas. Telemental health can be used to connect people to activities and support are in the service user's area or nearby areas.
- For students, telemental health has been easier for them to access in some ways because they feel at home in the online world and in control in the technical sphere – not a new physical environment and there is the control that you can leave at any time and close your laptop and you don't have to be there anymore. It has helped students who are anxious, but also those with accessibility issues. Being able to access things online and advancements in communicative technology have been great for those with accessibility issues and things that they have needed for a while, but now it is needed for a more mainstream audience. It has helped them, nevertheless.
- Especially if you have children and young people in period of crisis, if they are struggling to deal with issues in the family, having easy access to telemental health has been particularly helpful. It's easier to offer frequent contacts during a crisis via telemental health, compared to if someone has to attend for appointments face-to-face.
- For adolescents who are back in school, it can be convenient to have a telemental health session as it saves time as they do not have to miss school, as long as they can go to a private space.

**Caveats and pitfalls identified by stakeholders:**

- Sometimes the convenience of not having to travel is not an improvement, as people appreciate the process of the travel to and from the session if it means they can engage more in a face-to-face session. This also prevents bad feelings associated with treatment being in your home, you can leave it behind at the session.
- Some service users believe that telemental health created more barriers than opportunities, as not all service users took up telemental health at the start of the pandemic.
- Many members of service user groups were excluded when moved online. They need digital skills acquisition and access to technology and internet connectivity.
- Online support might be useful for milder mental health presentations, and it should be part of the toolbox but not used across the board as a substitute for face-to-face care.
- NHS phones do not always allow apps – technology in the NHS is behind what is wanted and needed by the population, and tended to prioritise security and uniformity (e.g. in using Teams) rather than technology that is familiar to service users.
- Not being able to use interactive and practical exercises and play as is usual in face-to-face contacts results in challenges engaging younger children, and using technology independently is also problematic for this group.
- It is considered that telemental health improves things for disabled people, but this is mainly for those who have mobility issues. For sensory issues telemental health probably does not improve things.
- Other barriers are increased e.g. poverty - cost of data or Wi-Fi.
- The provision of telemental health should not lead to relocating service users to geographically remote (e.g. sending someone from London to Manchester due to bed availability).
- Crisis houses often have very poor and unreliable Wi-Fi so that service users need to use their phone data (something not accessible to everyone).
- Home visits are a key part of community mental health services in many countries, and this may also be a solution for those who cannot or don't want to travel to appointments. Telemental health is not the only solution for these individuals.

#### CMO 2.3: Involvement and support for family and friends

When family and other supporters are invited (with service user agreement) to join telemental health sessions (C), this may result in more holistic treatment planning and greater engagement of family and others in supporting service users (O1), may help improve therapeutic relationships and treatment success (O2), increase engagement (O3), reduce some uncertainty and anxiety around treatment (O4), and may increase satisfaction of and support for family and friends (O5), as family and other supporters may be able to participate in care planning meetings and assessments that they would have found it difficult to attend face-to-face, increasing their engagement in supporting service users and their understanding of their difficulties and care plans (M).

**Underlying CMOs:**

- Brooks et al. (2013) When adapting an existing PTSD telemental health program for American Indian veterans (C), involving family in remote monitoring (M) improved the success (not defined in paper) of the programme (O)
- Liberati et al. (2021) When assessments were conducted remotely because of COVID-19 (C), if carers, family and friends weren't able to participate in the assessment (M), staff thought service users were disadvantaged (O).

- Lodder et al. (2020) When delivering a Zoom online intervention to parents of children with autism (C), both parents were able to join the sessions, which would have not been possible face-to-face (O) as they were able to join from home (M).
- Moslehi et al. (2021) For an inpatient with schizophrenia and COVID-19 requiring isolation (C), when the ward team facilitated regular video consultations allowing the patients next of kin to communicate with their son and the team (M), then this allowed the family to contribute in a meaningful way to their son's care (O).
- Moslehi et al. (2021) Among the family members of an inpatient with schizophrenia and COVID-19 requiring isolation (C), video consultations reduced the anxiety of the family members (M), and then this resulted in an improvement of carer satisfaction scores received by the ward (O).
- Moslehi et al. (2021) Among the family members of an inpatient experiencing first episode psychosis who had COVID and required isolation (C), when his parents could virtually attend and participate in care planning meetings, and observe the doctors, nurses and ward environment (M), then this reduced some of their uncertainty and confusion surrounding his treatment and greatly alleviated their anxiety about his care (O).
- Whistance et al. (2021) Amongst carers for individuals with dementia (C), the flexibility of video calling meaning they do not need to travel or leave home (M) improves access to support and appointments (O) because they it helps fit these around their other dementia caring responsibilities (M).

**Supporting information from stakeholder consultation:**

- When supporting children and young people with ADHD and autism and their families, input from schools and social is helpful and is much easier to coordinate online than in person. A digital interview works better as everyone can dial in. Reassuring to include lots of different voices involved in one patient's care and then patients feel that things are being addressed.
- Telemental health makes it easier to involve both parents, even if they one or both are working and might not otherwise be able to attend appointments.
- Remote therapy takes the focus off the therapist and puts it on the parent/carer. People (from non-chaotic homes) had positive feedback about feeling more empowered and able to contribute.
- It has been helpful for service users whose families are international to maintain contact, and to collaborate in the care plans. Families have been emailing queries, attending ward rounds on MS Teams, and Face Timing with service users.
- In inpatient and crisis wards, several Trusts rolled out iPads as patients could not have face-to-face visits from friends/family on wards mid-pandemic - in-person visits now re-introduced but many have kept iPads.
- One advantage is that relatives of inpatient service users who live abroad have been able to join ward rounds and be involved in care.
- Staying in touch with relatives is very important. In forensic units and forensic care, the signal is poor so people cannot use mobile phones, and limited access to anything digital, meaning these people are all digitally excluded.

**Caveats and pitfalls identified by stakeholders:**

- There have been young people who have needed mental health support, but their families did not speak English (although the young person did). The pandemic may have made it harder for those who do not speak English unless satisfactory interpreting arrangements are made.
- Provision of telemental health should not lead to restriction of seeing carers/relatives face-to-face. Stakeholders expressed concern that funders might not fund travel for carers/relatives any more due to access to telemental health.
- It is also important to note that not all service users would want their family to be involved in their care.

- Families and carers of children may have had less of a say in care pathways during the pandemic.

#### CMO 2.4: Widening the range of available mental health services and treatments via telemental health

For service users who may benefit from services that they would not usually be able to access locally and which are connected to regional and national specialist services (C), telemental health can widen the range of specialist assessment, treatment and support available (M), which potentially leads to improved access to services tailored to individual needs and culturally appropriate or specialist services (O1), and to improved satisfaction and treatment outcomes (O2), although an impoverished range of local face-to-face provision may be a risk if referral to distant specialist care via telemental health becomes routine (O3).

**Underlying CMOs:**

- Colle et al. (2020) Among secondary mental health service users during a switch to online appointments due to the COVID-19 pandemic (C) if/when patients had hearing impairments (M) then face-to-face appointments were needed unless telemental health was adapted to their needs (O).
- Crowe et al. (2016) Using video conferencing with a therapist who is trained in American Sign Language (ASL) (M) resulted in high satisfaction with telepsychiatry (O) for deaf or hard of hearing service users in remote areas in the USA ©.
- Crowe et al. (2016) Among people with hearing impairments living in remote areas (C), providing videoconferencing and a therapist trained in ASL (M) may improve their access to care and thus their mental health (O).
- Disney et al. (2021) When working with clients who require a translator (C), using telemental health made it harder to find an interpreter, meaning it was harder for clinicians to understand the service user (M), which impacted quality of treatment and therapeutic relationship (inferred) (O).
- Zheng and Gray (2014) For service users who do not speak the native language and therefore could not access therapy in their local area (C), providing therapists who spoke the language of the service users using telemental health (M) removed geographical barriers to care and enabled them to access PTSD therapy (O).

**Supporting information from stakeholder consultation:**

- Telemental health might remove the need for an interpreter entirely as it can connect people who speak the same language. This is something that in theory works very well, but in practice it may be challenging (e.g. due to insurance coverage limitations). There are examples of wards in London providing iPads that were then used for translation, which was very useful.
- There are also cultural perspectives that may need to be considered. There may be benefits of therapists having cultural understanding of mental illness, and understanding the stigma of having mental illness, and this may be more possible to access using telemental health. This has not been widely observed in practice in the UK yet.
- Telemental health interventions might also facilitate the organisation of smaller groups to discuss cultural differences; it is possible to convene service users from all over the country who share a cultural background, and this peer support can be powerful.
- Service users can access services in a different city/place, which means they are able to access tailored support (specialists, services for marginalised communities including peer support).

**Caveats and pitfalls identified by stakeholders:**

- Service users expressed concern over the possibility that offering telemental health services might be used as a justification for not offering face-to-face services in remote areas, or for concentrating more specialist services in a small number of centres.
- Sometimes the convenience of not having to travel is not more convenient: people appreciate the process of the travel to and from the session if it means they can engage more in a face-to-face session.
- While telemental health might in theory a good way to access a range of interpreters, booking good quality interpreters for online consultation is reported to have proved challenging in practice. Phone interpreting is sometimes not a very satisfactory experience.

#### CMO 2.5: Face-to-face appointments required for those with sensory or psychological barriers to telemental health

Offering face-to-face (or telephone) appointments to people who struggle to cope with sensory (visual or auditory) aspects of telemental health or have symptoms that are exacerbated by it (C), may help to improve engagement with mental health care (O) as these symptoms and sensory or cognitive impairments might be less affected by face-to-face compared to telemental health care and service users are able to access their preferred modality of care (M).

**Underlying CMOs:**

- Chen et al. (2020) Among patients with specific medical and mental health conditions, such as auditory and/or visual impairments and migraine headaches, attending a general psychiatry outpatient service in the USA (C), reliance on telecommunications and/or screens (M) limits the engagement of this group with therapy (O).
- Costa et al. (2021) During the COVID-19 pandemic in the USA, some people with certain conditions (e.g. concussion or dementia) or older age who said that they were coping well during the pandemic and felt well supported by their providers nonetheless identified limitations of telemental health (C) which might make their health problems worse (O), as these service users felt less able to deal with screen time (M).
- Healthwatch (2021) For one young person with autism who was startled by the sound of a phone ringing (C), if there was a delay in the call or if there was a problem with the connection resulting in the call only coming through later (M), then led to significant anxiety such as they were then unable to use the phone (O).
- Holland et al. (2020) One participant who required face-to-face appointments due to experiencing command hallucinations which prevented her from engaging with telephone therapy (C), as she could maintain eye contact with her therapist (M), she felt safer, and the sessions were more productive (O).

**Supporting information from stakeholder consultation:**

- Specific difficulties such as paranoia and voices can make telemental health a difficult and unpleasant experience. Some people find it difficult to see themselves on video calls: supporting them to switch off their image on calls can be helpful.

**Caveats and pitfalls identified by stakeholders:**

- There is an assumption that all users see/hear, but there needs to be consideration around how to deliver equitable service for those with visual and hearing impairments, e.g., captions to video calls for people with hearing impairments.

#### CMO 2.6: Inclusion of multidisciplinary and interagency teams in meetings

When mental health consultations are conducted using telemental health, this enables inclusion of people in appointments who are based geographically far away or who have schedules that would not have allowed them to join a face-to-face session (C), meaning care and support has potential to be more holistic and integrated (O), as it is possible for staff from different services and sectors to provide perspectives and contribute to plans (M).

**Underlying CMOs:**

- Peralta and Taveras (2020) When a special telephone, video call and electronic messaging service was enabled, organized by governmental and non-profit agencies and staffed by professionals including volunteer psychologists and psychiatrists in the Dominican Republic during the COVID-19 crisis (C), by mobilising and facilitating contact among a multidisciplinary and multi-institutional team (M), this facilitated access to mental health services and facilitated a large number of teleconsultation interventions that would otherwise not have been possible (O).
- Whistance et al. (2021) Amongst professionals delivering dementia care across remote Wales using video consultations (C) have improved access and opportunities to collaborate with dementia experts (O) due to the timely and cost effectiveness of this platform over travelling (M).

**Supporting information from stakeholder consultation:**

- Some practitioners have really liked this about working with Teams and other video consultation platforms because they are not travelling from site to site and multidisciplinary working, especially between people in different teams, is facilitated. Multidisciplinary teams can benefit from remote working.
- A benefit of telemental health is that clinicians are able to talk with more than one person at a time, and people can join with consultations from further away.
- It has been easier for practitioners and social workers to get together and not have to travel – more convenient to have professional meetings and discussions to create better outcomes.
- In inpatient units, care coordinators have been able to join ward rounds using videocall, which can help efficiency as they do not have to travel.
- From an organisational point of view, for both tribunals and crisis care, not having to send people around the country is better. It is easier to combine experts from across the country online rather than in person - the same number of patients get discharged when tribunals are conducted online compared to when conducted face-to-face. However, it takes people time to get used to the idea.

**Caveats and pitfalls identified by stakeholders:**

- Service users may feel under pressure to include certain teams, family members or carers in their care on the basis that, with telemental health, it is more feasible to include them in their appointments. This must be avoided.
- Services need to avoid creating pressure for appointments to be digital whether people want it done that way or not. Travel time isn't factored in, then when people want someone to travel to them, it might no longer be a possibility.
- The potential for telemental health to connect service user to care coordinators, clinicians, social workers, relevant members of school staff, family and friends, should not be used as a justification to relocate service users far away from their support networks (e.g. from London to Manchester due to bed availability).

#### CMO 2.7: Continuing to offer face-to-face care

When service providers offer care of equivalent quality and timeliness face-to-face rather than via telemental health to service users who do not wish or do not feel able to receive their care remotely (C), this ensures that care can continue and that inequalities in provision are not created or exacerbated (O), because it provides a choice to service users (M).

**Underlying CMOs:**

- Colle et al. (2020) Among mixed groups of secondary mental health service users within community mental health teams and outpatient services during a switch to online appointments due to the COVID-19 pandemic (C), if patients did not have a private room at home (M), then face-to-face appointments were needed (i.e. could not deliver service using telemental health) (O).
- Dores et al. (2020) During the switch to telemental health due to COVID-19 at a general psychology service in Portugal (C), some clients who did not have access to private space or appropriate technology for remote therapy, preferred face-to-face contacts, or had problems managing new routines or caring responsibilities at home (M) had lower adherence to telemental health care (O).
- Foye et al. (2020) Amongst mental health nurses who had to adapt quickly to remote working during the COVID-19 pandemic (C), many reported their work was hampered (O) by logistical challenges such as poor internet connection, hardware availability, and patient lack of privacy to conduct appointments (M).
- Pugh et al. (2020) When chairwork such as empty-chair dialogues and role-play is moved online so service users could take part at home (C), therapists reported that clients struggled with a lack of a therapeutic space (M) and chairwork in the home could be unhelpfully intrusive and disruptive to the client's everyday life (O).
- Sehlo et al. (2021) Among service users of an online therapy site in Egypt during COVID-19 (C), a major advantage of face-to-face consultations was a feeling of more security (O) associated with the privacy of personal information (M).

**Supporting information from stakeholder consultation:**

- For some people attending face-to-face appointments, the time travelling there and back may be helpful for processing/dealing with the emotions released. It is harder to do this in your own home, and for some this may be a safety concern.
- A screening tool may help to provide clarity on whether telemental health is right for a given service user - it has been previously found that in Improving Access to Psychological Therapies (IAPT) services many prefer digital therapy, while in community mental health teams (CMHT) more want face-to-face.

**Caveats and pitfalls identified by stakeholders:**

- It is good to have procedures in place to rearrange appointments to face-to-face if a risk is perceived by clinicians, however it is not possible for clinicians to pick up on every risk relating to domestic abuse, for example, remotely.
- Screening tools to make decisions about needs and support can sometimes be counterproductive as they can induce people to stick rigidly to them in an unhelpful way. Important to instead ask service users about preferences in a discussion.

#### CMO 2.8: Communication between staff

| |
|---|
| When remote technology platforms are used to facilitate real-time communication between staff members, including managers or clinicians working in different teams (C), this can lead to improved efficiency and more convenient working and staff management (O1), improved communication and collaborative planning (O2), and process improvement opportunities (O3), as staff have the ability to rapidly share information, keep track of evolving telemental health procedures (for example, during emergencies) and make collaborative decisions (M). |
| **Underlying CMOs:**<br>• Johnson et al. (2021) When moving to telemental health (C), when remote working improved efficiency (allowing prompt responses, saving travelling time, potentially reducing environmental impact, increasing convenience for staff (including by allowing home working) allowing staff to connect easily with each other) (M), then it was seen as enabling staff and services to work more efficiently and effectively well (O).<br>• Khanna et al. (2020) Among staff, including psychologists and psychiatrists, working in community mental health teams and/or outpatient services for individuals with PTSD (C), when Microsoft Teams was used by staff to connect with each other, then it allowed for rapid feedback, shared decision making, and brief messages (M), then this increased the ability to action “idle ideas”, develop strategies, and increase communication (O).<br>• Khanna et al. (2020) Among staff, including psychologists and psychiatrists, working in community mental health teams and/or outpatient services for individuals with PTSD (C), if/when Microsoft Teams was used, allowing for rapid feedback, shared decision making, and brief messages (M), then some team members felt this eroded their work-life balance (O).<br>• O’Dell et al. (2021) Early after the transition to Tele-Behavioural Health services (when emails were primarily used for questions and problem solving) (C), when the PCBH leadership created a spreadsheet on Teams for collection, response and tracking of procedural details (M), then this led to less burdensome email traffic and adequate tracking of procedures (O). |
| **Supporting information from stakeholder consultation:**<br>• Clinicians can more readily join other teams’ meetings e.g. for liaison, joint planning, exchange of information about team practices and policies, or education.<br>• Working from home, at least at times, may increase efficiency and convenience. |
| **Caveats and pitfalls identified by stakeholders:**<br>• Online working may negatively affect staff’s work-life balance as there is less of a clear divide when work is done from home/outside normal working hours. Pressure to keep working without breaks from work and screen exposure (e.g. for travel) might also be detrimental for well-being: further investigation of telemental health impacts on staff well-being is warranted. |

### Domain 3. Safety, Privacy and Confidentiality

#### CMO 3.1: Lack of privacy

When accessing telemental health sessions without access to a private space or secure private connection (C), service users and staff are at an increased risk of being overheard (M1), potentially leading to breaches of privacy and confidentiality (O1), risk of harm to those in unsafe domestic situations (O2) and reluctance to speak openly about sensitive topics (O3). It may also cause some service users to experience frustration, distress and anxiety (M2) leading to impacts on service user engagement and interactions (O4) and reduced willingness to use telemental health and continue therapy (O5).

**Underlying CMOs:**

- Bierbooms et al. (2020) Mental health care professionals in community mental health services in the Netherlands who reported technical issues and lack of technical devices/internet connection, lack of privacy and confidence in security of the system (C) reported lack of success in making connections and of confidence that calls were private and secure (M) resulting in frustration and less positive attitudes towards telemental health (O).
- Boldrini et al. (2020) For service users receiving psychotherapy at mental health services in Italy during the COVID pandemic who do not have access to a private space (C), when staff did not pragmatically discuss the difficulties service users faced in achieving an intimate, reassuring and safeguarded setting in which to participate in tele-psychotherapy sessions (M) led to higher rates of interrupted care (O).
- Boldrini et al. (2020) Service users receiving psychotherapy at mental health services in Italy (C) who did not have access to a private space to use for appointments (M) experienced higher rates of interrupted care during the Covid-19 pandemic (O).
- Dores et al. (2020) During the switch to telemental health due to Covid-19 at a general psychology service in Portugal (C), some clients who did not have access to private space or appropriate technology for remote therapy, preferred face-to-face contacts, or had problems managing new routines or caring responsibilities at home (M) and thus had lower adherence to telemental health care (O).
- Frayn et al. (2021) For people experiencing binge eating spectrum disorder attending virtual therapy (C), technical barriers and "logistical issues" related to home environment e.g. internet connections, lack of privacy (M) may "hinder" engagement with therapy (O).
- Ghosh et al. (2021) Among service users of a substance disorder centre in India with a stepped tele mental health model (C), sometimes patients used devices of family and friends (M), which could threaten the privacy and confidentiality of information (O).
- Ghosh et al. (2021) Among service users of a substance disorder centre in India with a stepped tele mental health model (C), sometimes family members were seen to be mostly around during the videoconference, even when it was done from patients' personal devices. For some cultures the concept of personal space and privacy is blurred in the family context (M) which could threaten the privacy and confidentiality of information (O).
- Hawke et al. (2021) Young people accessing mental health services via virtual platforms (C) who experience a lack of privacy or security during sessions (e.g. living at home, having distractions, security concerns regarding digital connections) (M) are more likely to experience distress and anxiety when using virtual platforms (O).
- Healthwatch (2021) Among young people attempting to avoid being overheard in the home (C), some take calls outside (M), which has led to further barriers to receiving care (such as noise issues, including the wind, device's battery life) (O).
- Hensel et al. (2021) Among therapists working with postpartum women experiencing depression and/or anxiety in Canada who opted to have therapy via video (C), when those women had other

children in the home or lacked privacy for other reasons (M), therapists felt this interfered with the therapy (O).

- Liberati et al. (2021) Staff, carers, and service users who were previously using secondary mental health care before COVID, when they transferred to remote care (C) if they missed the privacy and safety of the consulting room and felt their private space (at home) was 'invaded' (M) they weren't able to make the full use of therapeutic interventions (O).
- Lodder et al. (2020) When delivering an online intervention to parents of children with autism (C) parents had problems fully concentrating on what other people were saying (O) as they were interrupted by children talking and asking for their attention (M).
- Mad Covid (2020) Amongst service users receiving digital mental health care remotely (C) staff should seek to create a safe and private environment (e.g. no team members walking around in the background) (M) and check in with the service users if they are able to have a private space (M) to avoid cancelling and increase engagement with sessions (O).
- McBeath et al. (2020) Among mixed psychology service users receiving online psychotherapy (video-link platforms and telephone) from home during COVID-19 (C) if they had people or family around (e.g. in the home) during psychotherapy (M), then clients reported feeling unable to engage as they didn’t want to risk becoming upset in the home environment (O).
- McBeath et al. (2020) Among mixed psychology service users receiving online psychotherapy (video-link platforms and telephone) at home during COVID-19 (C) if they had no confidential space to talk in or were living with the person who was the main problem (M) then the client found it difficult to talk knowing that people/that person was nearby (O).
- Medalia et al. (2020) Among demographically homogenous, high socioeconomic status mixed secondary mental health service users using community mental health teams and outpatient services in NYC during a conversion to video supported telehealth due to COVID-19 (C), (if/when) the service users had living spaces which provided privacy (M) then this supported the conversion to video supported telehealth (O).
- Mental Health Network NHS Confederation (2020a) Among staff who were delivering group-based remote services (C), the trust suggested a range of ways that individuals could protect their own confidentiality and that of their fellow users on the call (M). Examples included disguising or blocking their face or the use of an avatar, and blurring the background and using a fictitious name (with agreement between clinician and user in advance.) This broadened the responsibility of protecting confidentiality to all attending the call (O).
- Mind (2021a) People having online therapy in their home environment (C) felt unsafe and feared being overheard (M) which impacted how much they were willing to share in these sessions (O).
- Mind (2021b) Young people accessing mental health support (C) felt uncomfortable speaking by phone or online using remote mental health support (M) and were concerned about privacy when using telephone or video call, e.g. being overheard at their home or where the health professional was working (M) thus were less likely to engage with this type of support (O).
- Oqueji et al. (2021) Among the general population in Nigeria during COVID-19 (C), as there were concerns about being overheard and lacking a private location to speak (M), then there was a resulting lack of willingness to use e-therapy (O).
- Pugh et al. (2020) When chairwork such as such as empty-chair dialogues and role-play is moved online so service users took part at home (C), therapists reported that clients could benefit from being in a safe space (M) which may make them feel as though they have more ownership of the therapeutic process (O).
- Shklarksi et al. (2021) For service users who did not have a private space to have therapy at home (C), using the chat function on Zoom (M) meant they could discuss sensitive topics without being overheard (O).
- Simon et al. (2021) patients using internet-based interventions at home (C) when they may be overheard by others nearby others (M) may have challenges to their boundaries and sense of safety (O).

- Uscher-Pines et al. (2020a) When moving to remote care (C) if this led to 1) decreased clinical data for assessment (physical exam) 2) diminished patient privacy 3) increased distraction in the patient's home setting (M) then this affected the quality of provider-patient interactions (O).
- Uscher-Pines et al. (2020a) When offering telemedicine to patients who might be concerned about privacy for the first time (C) if psychiatrists take steps to ensure that the patient is in a private place for visits, e.g., by offering and brainstorming about options for finding a private place, or otherwise reschedule the session (M) then this improves the quality of the telemedicine visits (O).
- Uscher-Pines et al. (2020a) When offering telemedicine to patients who are self-conscious about video visits and don't want clinicians to see their home environment for the first time (C) if staff provides FAQs that explain how to change the backgrounds to not show the surroundings on Zoom (M) then this improves the quality of telemedicine visits (since it increases privacy and patients feel more comfortable (inferred) (O).
- Watson et al. (2021) Among service users with psychosis accessing psychological therapies before COVID-19 (C), if they had data privacy concerns (M), then they declined remote therapy (O).

**Supporting information from stakeholder consultation:**

- For both service users and staff, private homes with others present are a problem because these others are not bound by confidentiality agreements like NHS staff are.
- Clinicians should disclose to the patient if they are not in a completely private environment, for example a shared office or a private home with other family members present. Telemental health may not be appropriate if clinicians cannot access a private space. Headphones should be available to staff and may be helpful in shared environments.
- It is important that clinicians check that service users can find a space they feel both physical and psychological privacy, and this was raised as a particular concern for children and young people, especially if there are family difficulties. It is important to continuously check whether young people are in a safe and private location, rather than checking once and assuming this remains the case for the whole session, given that family members could move around the young person's home.
- Noisy or busy environments can be distracting and thus impact the therapeutic relationship and engagement.
- It should be a given that staff are  in private and non-distracting environments. Staff and service users should be transparent if there are potential environmental interruptions, such as children being in the background or colleagues in the office.
- Staff should be aware of how to deal with the impact of environmental distractions on telemental health sessions.

**Caveats and pitfalls identified by stakeholders:**

- Private spaces should also prevent people from being seen by others so that they also have psychological privacy- example of providing glass boxes situated within mental health centres to service users for their telemental health sessions. While this ensures physical privacy (i.e. not being overheard by others and not being in your own personal space), it doesn't provide psychological privacy as others can still see you during your telemental health session-service users felt they were in a "fishbowl" environment, and this was stigmatising.
- Provision of private spaces is not just for those who do not have one, but also for people who do have one but do not want this private space to be invaded (for example a bedroom). This may be particularly salient for inpatients.
- Any strategies that involve provision of spaces for privacy need to consider accessibility e.g. they should be wide enough for wheelchair users.

- One potential strategy discussed to provide spaces for young people was working with schools to provide spaces away from home that can be used during school hours. This is possible, but issues of this are that many young people would not want to alert teachers/other pupils to their need for a space to use for therapy. Ways to provide this in a confidential way should be considered.
- Potentially distracting elements in the service user's or staff's environment can also serve as a means to develop a therapeutic relationship, e.g. talking about a person's pet or instrument in the background.

**Additional sources utilised:**


- Barratt C. Mental health and houses in multiple occupation: The challenge of turning research into practice. [31 January 2022]; Available from: https://www.emcouncils.gov.uk/write/Mental_Health_and_HMOs_Caroline__Barratt.pdf.
- De Witte NA, Carlbring P, Etzelmueller A, Nordgreen T, Karekla M, Haddouk L, et al. Online consultations in mental healthcare during the COVID-19 outbreak: An international survey study on professionals' motivations and perceived barriers. Internet Interventions. 2021;25:100405.
- Moore J. Severe mental illness and Covid-19: Service support and digital solutions 2020.
- The British Psychological Society. Considerations for psychologists working with children and young people using online video platforms. 2020.


#### CMO 3.2: Privacy, anonymity, and reduced stigma

For some service users who feel stigmatised when attending a mental health service in person and who have access to a private and secure space to receive therapy remotely (C), being provided with the option of telemental health as an alternative means there is an option to receive care with more anonymity (M), which helps ensure their privacy and safety (O1), thereby increasing the accessibility of services (O2).

**Underlying CMOs:**

- Chen et al. .(2020) Due to the psychiatry service changing to remote working due to Covid-19, eliminating the need to travel to psychiatric clinics (C) can increase privacy and decrease stigma related barriers to treatment (M), which in turn can help increase accessibility of mental health care (O).
- Choi et al. (2014) When older adults (many of whom lacked transportation) received problem solving therapy using video conferencing software rather than face-to-face care (c), they found telehealth acceptable (O) as they liked the increased convenience and additional privacy of accessing therapy at home (M).
- Kanellopoulos et al. (2021) When a psychiatric inpatient service provided electronic tablets to inpatients who were isolated in their room during a rapid switch to telemental health due to COVID-19 (C) IF the service purchased temporary email addresses from a software company that patients could use to access telecommunications software and that were kept in a "directory" on a secured server so that staff could contact patients directly (M) this protected patient privacy (O).
- Sehlo et al. (2021) Among service users of an online therapy site in Egypt during covid-19 (C), a major advantage (O) of online consultations was avoiding embarrassment and stigma attached to attending a psychiatric clinic (M).
- YoungMinds (2020) Offering remote interventions to young people with diverse gender and sexual identities (C) may increase their access to mental health care (O) as remote interventions can reduce stigma and shame related to mental health problems and accessing mental health support (M).

**Supporting information from stakeholder consultation:**

- Young people may think about privacy of information differently, e.g. they do not realise the problem of answering a teletherapy call on a bus, for example. This may put them at higher risk as the information they share is not private.
- Feeling stigma may be less of an issue for people switching from face-to-face care to telemental health, as they may have already overcome stigma to attend the service prior to the switch. However, experiencing less stigma is a benefit for people beginning treatment and having the option to choose telemental may mean that more people feel able to access care.
- Evidence from the literature and stakeholder discussion suggests this may be especially relevant for young people who experience stigma, such as those with related challenges linked to their gender identity and/or sexuality (Youth Access, 2020)

**Caveats and pitfalls identified by stakeholders:**

- Service users feeling that they have more privacy is beneficial, but care should be taken to consider how much risk can perceived via telemental health: a lived experience researcher commented that for a telemental health session in the past they had made a lot of effort to appear 'fine' but didn't feel fine. Telemental health relies heavily on outward appearance and so risk may be missed.
- Telemental health can reduce anonymity as some staff members share office space and sessions can recorded and put on record. The feeling of reduced anonymity may be very specific to certain contexts.

**Additional sources utilised:**


- Healthwatch. Remote mental health survey. 2021.
- James K. Remote mental health interventions for young people: A rapid review of the evidence. Youth Access, 2020.
- Lannin DG, Vogel DL, Brenner RE, Abraham WT, Heath PJ. Does self-stigma reduce the probability of seeking mental health information? Journal of Counseling Psychology. 2016;63(3):351-8.
- National Health Service England. Mental health COVID-19 children and young people case studies. 2021 [31 January 2022]; Available from: https://www.england.nhs.uk/mental-health/case-studies/children-and-young-people-cyp-case-studies/mental-health-covid-19-children-and-young-people-case-studies/.
- YoungMinds. YoungMinds submission to the COVID Committee's inquiry into 'Living Online: The Long Term Impact of Wellbeing'. 2020.


#### CMO 3.3: Managing risk

When services provide adaptations to remote therapy to incorporate tailored risk management procedures for service users receiving remote care (C), this encourages consideration of the risks associated with remote care specific to each individual, including risk of self-harm or suicide as well as risk from others in situations of domestic abuse, and ensures staff are aware of the procedures to try to assess and respond to risk or safeguarding concerns despite challenges associated with remote care (M), which has the potential to improve the safety and wellbeing of service users and others (O1). However, a disadvantage of telemental health is that real-time risk assessment limits an immediate response to be organised when someone is at imminent risk of harm and some distance away (O2).

**Underlying CMOs:**

- Benaque et al. (2020) A dementia service in Spain (C) had detailed contingency plans (including preserving safety of both service users and staff, continuity of care and ethical responsibility) (M) which enabled them to successfully adapt to telemental health during the Covid-19 pandemic (O).

- Martin et al. (2020) For clinicians working with young people in an early intervention service (C), using digital communications such as text and email allowed them to follow up with the young person if usual contact was lost to encourage them to check in with the team (M), which allowed improved risk management (O).
- Martin et al. (2020) When digital communications via text and email were integrated into early intervention services for young people (C), setting clear boundaries and being transparent regarding staff availability and accountability protocols (M) prevented the communications between clinicians and service users becoming overly intimate (O).
- Martin et al. (2020) When an early intervention service for young people with psychosis facilitated treatment with digital communications between clinician and service user via text and email (C), if in the event of crisis messages were sent to personal accounts which may not be picked up until the following day or longer if staff were on holiday instead of 24/7 crisis lines (M), staff felt this option risked safety (O1) and young people felt abandoned (O2).
- Mental Health Network NHS Confederation (2020a) Among staff delivering group-based remote services for service users (C), the Trust assigned a clinical safety officer, who ran a hazard workshop using MS Teams to consider the possible ways that confidentiality could be breached in an online group setting (M). For example, a group member attending a group call from a place where a non-group member can see or hear the conversation without the knowledge of the rest of the group. By considering the range of breaches possible, they were able to consider and put in place mitigating steps to avoid them (O).
- Mental Health Network NHS Confederation (2020a) Among staff delivering group-based remote sessions for service users (C), having an open conversation with the service user joining the group about confidentiality and expectations was important (M). This created a safe space to ask questions and builds trust between the clinician and service user to reduce the likelihood of confidentiality breaches taking place (O).
- O'Dell et al. (2021) As behavioural health providers (BHPs) worked from home and patients could be at home or in the clinic (C), existing plans and procedures for high-risk patients were revised. including for clinic staff in crisis situations, correspondence between team members via electronic means, and increased co-ordination with patients/ families to facilitate safe transport to Emergency Department if inpatient psychiatric hospitalization was indicated) (M), in order to manage crises safely and decrease patients' likelihood of presenting in the Emergency Department for behavioural health concerns (O).
- Pierce et al. (2021) Among psychologists who delivered primary care and psychology /psychotherapy to service users in a variety of mental health care settings which moved to telepsychology during the COVID-19 pandemic (C), if they perceived safety concerns for treating antisocial personality disorder, behavioural issues and bipolar disorder using telepsychology (M) then they tended to be reluctant to use telepsychology to treat these issues (O).
- Rosen et al. (2020) Among veterans' mental services working remotely (C) if services are planning in advance (e.g. having backup phone numbers, identifying an emergency contact, knowing what local emergency services are available) and have established protocols for managing suicidality and for remotely triaging patients (M) then clinicians can respond to a crisis immediately (O).
- Vera San Juan et al. (2021) Among mental health service users who were accessing video support groups during COVID-19 (C), when there was an incident of risk or self-harming behaviours on camera during the session (M), then participants experienced severe distress during or after the session (O).
- Vera San Juan et al. (2021) Among mental health service users who were accessing video support groups during COVID-19 (C), when there was an incident of risk or self-harming behaviours on camera during the session (M), then new ground rules, risk management and support were put in place in subsequent sessions (O).

- Smith and Gillon (2021) Therapists concerned about ethical and risk issues in online therapy (C) would adopt methods such as personal safety checks (M) to ensure the client and their own safety (O)
- Uscher-Pines et al. (2020a) When offering telemedicine for the first time and patients are in a different location than the psychiatrist (C) if psychiatrists ask patients for their location and a call-back number at the start of each session (M) then this is helpful in the case of an emergency (O).
- Wilson et al. (2021) When switching to remote perinatal consultations among women experiencing domestic abuse (C) if they are not able to be honest about DVA, symptoms and risk during video or phone consultations given their abuser may be present (M) then this poses a barrier to risk assessment and actioning safeguarding concerns (O).
- Holland et al. (2020) One participant who felt unable to engage with telephone therapy due to command hallucinations (C), felt much safer and had more productive sessions when offered face-to-face care (O) as she could maintain eye contact with her therapist (M).
- Kanellopoulos et al. (2021) When a psychiatric inpatient service provided electronic tablets to each of the patients who were isolated in their rooms due to COVID-19, staff did daily risk and safety screenings before patients were given a tablet (C) to determine ongoing eligibility for tablet use (M) and thus ensured patient safety (O).
- Liberati et al. (2021) During the move to remote care during Covid, staff providing remote care from home doing some types of therapeutic work such as trauma processing (C) if they (staff) felt unsafe because of the removal of the boundary between private and therapeutic work (M) they paused the care (O).

**Support information from stakeholder consultation:**

- Due to the common problem of connection instability, having a strategy in place for what to do when line breaks, particularly if during a sensitive point in the call is extremely important. One webinar attendee mentioned that they developed a manual for service users on what to do if a call ends abruptly in the middle of a sensitive moment in the session. For example, instructions on alternative points of contact or remote tools they can use.
- Some arrangements in place for face-to-face settings need to be adapted for remote therapy, for example, safeguarding arrangements. It is more difficult to accurately establish the family circumstances and put in adequate safeguarding measures for young people during remote consultation, and it is more difficult to assess the risk and act quickly.

**Caveats and pitfalls identified by stakeholders:**

- Fast, in person responses are unlikely to happen with current mental health service strains.
- Risk to the patients should be considered *before* providing telemental health (to ensure successful risk management, *not* to prevent certain service users from receiving it).
- Unlike face-to-face sessions, people can easily disengage from the call. This is their right; however this could happen during a difficult moment and so could pose a risk to the service user or result in concerns about whether risk has been fully assessed or how to respond to it.
- Also important to moderate peer-to-peer platforms to try to understand and manage risks.
- For staff working remotely, we make assumptions that staff have all these things that they do not necessarily have (devices, connection, skills etc). We are changing the way staff are working, including a lack of peer support for staff. If a risk situation comes up, there are always people around to call but it does not feel the same in terms of team connection, support and thinking.

#### CMO 3.4: Technological support and information security

| |
|---|
| When services provide technology support, software with appropriate security and devices (including mobile phones and headphones) to staff specifically for work use (C), this helps ensure privacy and confidentiality for both service users and staff (O), because staff can store information securely on devices that are not shared with others (M1) and are able to ensure that service users are aware of when they will have access to their work devices (M2). |
| **Underlying CMOs:**<br>• O'Dell et al. (2021) Among behavioural health providers providing telephone therapy using Google Voice (C), as Behavioural Health Providers did not realise patients could send text messages (M), this resulted in an incident of a patient texting the BHP overnight during a mental health crisis [when support couldn't be given] (O).<br>• O'Dell et al. (2021) Among behavioural health providers (these are individuals, rather than a health 'provider') providing telephone therapy to patients using their own personal cell phone (C), they struggled to keep their own cell phone numbers confidential (M), leading to the team trialling several platforms to route calls through local clinic numbers (O). |
| **Supporting information from stakeholder consultation:**<br>• Whether personal equipment is used by clinicians and whether this is safe is an issue- for example through sharing equipment with family members. The NHS still has people sharing devices which can cause data safety issues.<br>• It is assumed that staff have the expertise and knowledge to safely use technology to conduct telehealth sessions, and that they have the correct equipment. It may also be assumed that staff can be as capable from home as in an office where there is often someone to ask for support. Staff have the added stress of trouble-shooting problems with connection for service users, even when they don't feel very knowledgeable on the subject.<br>• It is often assumed that service users will be more concerned with privacy than the clinician, but this may not always be the case- a lot of clinicians worry that they will be recorded, and this will be used to judge their performance.<br>• It is important to set boundaries for both service users and clinicians in relation to privacy of personal life (through not having to use private devices for work) and maintaining a work-life balance (through not being contacted outside of office hours due to being called on private mobile phone). |
| **Caveats and Pitfalls identified by stakeholders:**<br>• Practitioners and service providers struggle with balancing service-user preference with risk e.g., what platforms they are allowed to use/feel capable of using safely.<br>• Publicly available Wi-Fi may have security/firewall issues and therefore may not be appropriate.<br>• Trusts need to be able to critically evaluate the safety of the tools that are in the market. Introducing technology can be hard and dependent on how ready trusts are to use a variety of digital tools and do that safely. There is a balance to be struck between safety and the benefits of using familiar platforms. |

### Domain 4. Therapeutic Quality and Relationship

#### CMO 4.1: Change in non-verbal cues and informal chat impacting the therapeutic relationship

When switching from face-to-face to telemental health care (C), some service users and particularly staff perceived the relationship to staff, service users, and/or other service user group members to be negatively affected or found it more difficult to develop a therapeutic relationship (O1) and were thus less willing to take up or use telemental health (O2), more likely to be dissatisfied (O3), and viewed care as less effective compared to previously received face-to-face care (O4). This was because they perceived telemental health to be impersonal and found it more difficult to discuss personal information due to a lack of non-verbal feedback, eye contact, and social cues as well as informal chat before, after, and during sessions (M).

**Underlying CMOs:**

- Bambling et al. (2008) Amongst counsellors providing online support via text-based interventions (C) the lack of social cues and the time delays in responses (M) creates a sense of disconnection and may lead to misunderstandings (M) that create strains on the relationship and may result in a lack of therapeutic bond (O).
- Chen et al. (2020) A general psychiatry service in the USA that switched to telemental health due to Covid-19 (C), found that the technology does not yet fully capture the richness of an in-person interaction, and the sense of intimacy provided by a closed-door office space (M), which negatively impacted therapeutic alliance (O).
- Disney et al. (2021) Refugee mental health providers working in a service for refugees (C) who had their first appointment with their client using telemental health (M) found it harder to develop a therapeutic relationship (O).
- Eagle (2020) Amongst service users receiving video therapy (C) the impersonal nature of video calls and lack of non-verbal communication cues (M) lead to feelings of invalidation and poor mentalisation (M) which had an impact on the therapeutic relationship (O).
- Fogler et al. (2020) Moving a parents therapy group (for parents of children with ADHD) online due to the Covid-19 pandemic (C) resulted in some parents finding it harder to discuss personal information due the impersonal nature of the online setting (M), which negatively impacted rapport between group members (O).
- Frayn et al. (2021) For people experiencing binge eating spectrum disorders (BESD) using teletherapy during the pandemic (C), perceptions of this method as impersonal reduced feelings of connection [with the therapist] (M), leading to views of reduced effectiveness and of dissatisfaction with the method/having preference for face-to-face) (O).
- Grover et al. (2020a) When moving to telemental health (C) if staff experienced certain challenges such as poor rapport (M) then this led to a lower level of satisfaction (O).
- Hawke et al. (2021) Young people (C) who have previously been connected to mental health services (M) are more likely to consider using virtual mental health services (O).
- Healthwatch (2021) Among young people who are starting interventions (C), if the professional goes straight into the appointment and does not spend time at the beginning getting to know the young person (M), then this is a barrier to the young person wanting to open up and being able to trust the professional (O).
- Hensel et al. (2021) Among postpartum women undergoing therapy for depression and/or anxiety in Canada (C), if those women had an existing relationship with and trust in their therapist (M), they felt more able to take up the offer of video therapy (O).

- Johnson et al. (2021) When moving to telemental health (C) if a client is new (M) then clinicians find it harder to conduct an assessment and establish a good therapeutic relationship (compared to face-to-face and established clients) (O).
- Juarez-Reyes et al. (2021) Within study participants (people in primary care for stress) who were transitioned to video sessions due to shelter in place orders (C) if the participants had met each other in person beforehand and built trust (M) then some participants attributed the trust to meeting in person and said they felt comfortable and willing to share in subsequent video conference groups (O).
- King et al. (2006) For young people accessing support via a text-based system (C) the lack of non-verbal feedback and social cues (M) create a space of lower emotional intensity (M) which may lead to misunderstandings which create strains on the relationship (O).
- Liberati et al. (2021) Staff, carers, and service users who were previously using secondary mental health care before COVID (C) when they transferred to remote care, noticed that non-verbal cues which were important to consultations were missing (M) which in turn may have affected the therapeutic relationship (O).
- Liberati et al. (2021) For people who were already using secondary mental health services before COVID-19, when services moved to remote (C) if they had already had a relationship with a clinician (M) they were more likely to report positive experiences of remote care (O).
- Lodder et al. (2020) When delivering a Zoom online intervention to parents of children with autism (C) this affected the communication style compared to face-to-face sessions (O) as facilitators had to work harder to make sure everyone would get a chance to speak, and participants talked less freely compared to face-to-face settings (M).
- Lodder et al. (2020) When the first session of a Zoom online intervention for parents of children with autism was face-to-face (C) people felt more comfortable opening up and sharing their personal experiences with the rest of the group during the remainder of the online intervention (O) as they were able to build trust during the first face-to-face session (M).
- Lodder et al. (2020) When delivering a Zoom online intervention to smaller groups of 6 of parents of children with autism compared to larger groups (C) it was easier to create a more natural group conversation and facilitating was less challenging (O).
- Mind (2021a) For people from both a minority ethnic and white background (51% and 65% respectively) (C) not being able to speak face-to-face and gauge therapist reactions (M) made it more difficult to engage in therapy (O).
- Ogueji et al. (2021) Among the general population in Nigeria during COVID-19 (C), as there is a lack of visual and body language cues in e-therapy which may mean that nonverbal cues are missed by health professionals (M), then this is a barrier to willingness to use e-therapy (O).
- Olwill et al. (2020) Among psychiatrists with lower levels of training working in community mental health teams and outpatient services for a mixed group of secondary mental health service users in Dublin during a conversion to telephone consultations due to COVID-19 (C), (if/when) they struggled with the change in visual cues this (M) then this negatively affected rapport with the client (O).
- Pugh et al. (2020) When chairwork such as empty-chair dialogues and role-play is moved online (C), therapists reported that clients found it harder to go deeper in their therapeutic work compared to face-to-face (M) which led to online chairwork being viewed as less acceptable (O).
- Sasangohar et al. (2020) Among team members and clients who find it difficult to generate and sustain interpersonal connections when delivering/receiving telemental health (especially new clients who lack rapport) (C) IF social signals and efforts are amplified (M) then this can help overcome loss of intimacy and belonging derived from physical proximity (O).
- Sehlo et al. (2021) Among service users of an online therapy site in Egypt during covid-19 (C), a major advantage (O) of face-to-face consultations was a feeling of direct contact between the doctor and the patient (M).

- Severe et al. (2020) Among paediatric patients receiving telemental health (C) when the children had difficulties focusing and building a therapeutic relationship using telemental health while using video technology (M) then this decreased the likelihood of wanting to receive telemental health in the future (O).
- YoungMinds (2020) Young people accessing mental health support digitally (C) find building a trusting relationship challenging via video or phone modes as they are unable to read and provide social cues (M) thus find this method of support less effective than face-to-face (O).
- Vera San Juan et al. (2021) Mental health service users who started mental health support or changed their therapist/psychiatrist during the COVID-19 lockdown (C), found that the relationships with the new care provider often felt less personal compared to face-to-face (M), which was felt to affect the quality of their care negatively (O).

**Supporting information from stakeholder consultation and literature:**

- It is difficult to develop a welcoming, personal, and non-threating environment via telemental health. There is often less small talk before, during, and after sessions when using telemental health.
- Communication is impacted by a limited view on the other person's body language, such as restlessness, and lack of eye contact. It is important to use high quality equipment and good camera placement during video calls (LeRouge et al. 2015; Lopez et al. 2019).
- Non-verbal communication, such as pausing to reflect, feels less natural via telemental health. Staff should make greater efforts to communicate clearly, enhance gesture, and to provide verbal and non-verbal reinforcement, such active listening. It is important to employ service user-centred communication, such as being reassuring and supportive (Pinto et al. 2012).
- Consistent check-ins at the start of each session about experiences, preferences, and potential difficulties related to engagement and how to best address these when using telemental health are crucial.
- Telemental health is less suitable for first and last appointments, including assessments and discharge. Staff and service users might find it especially difficult to develop a therapeutic relationship when service users are new to a specific service or clinician, or to mental health care generally. Staff should take more time to get to know a new patient to facilitate developing a therapeutic relationship via telemental health. Service users, especially new services users, should also have the option to receive first and last sessions face-to-face.
- Staff with more experience might be more confident in interpreting service users' behaviour and communication. This helps with the lack of visual cues when using telemental health. However, staff appear more concerned about this than services users (Hubley et al. 2016; Lopez et al. 2019; Olwill et al., 2021). Staff should be offered training to increase their comfort with technology and to interpret social cues and conduct assessments when using telemental health (Lopez et al.,2019).
- Staff should receive reassurance that some services users perceive the therapeutic relationship to be less impacted by telemental health than staff do.
- Service users who are apprehensive of technology use; service users who are concerned about the violation of their privacy; and service users in crises situations might especially struggle with developing a therapeutic relationship. Staff should 1) take time to build trust and discuss concerns, such as risk and privacy, 2) involve family members (in agreement with service users), 3) be fully transparent about the purpose of appointments.
- Group therapy sessions are particularly challenging via telemental health as it is more difficult to facilitate engagement and interactions between group members. Staff need to balance sharing resources with group members being able to see themselves on screen and interact with each other. On some platforms it may not be possible to view all members of a group at once. The larger the group the more technical or connection problems are likely to arise. Additionally, Zoom etiquettes, such as having to raise one's hand to speak, impede the flow of conversations

further and staff need to work harder to make sure everyone gets the chance to speak (Lodder et al., 2020). This impacts relationships between group members, especially in groups larger than six (Lodder et al., 2020). Staff should facilitate peer-led, unstructured social time for service users group members.

- A lack of therapeutic relationship can impact engagement and commitment to sessions and thus lead to higher dropout rates.
- Greater active participation from parents to ensure intervention success compared to face-to-face delivery for young children with autism – telehealth relies on the caregiver to facilitate the clinician-child relationship. There is also the suggestion that interacting remotely is less intimidating for some children than face-to-face, whereas for particularly younger children, virtual interactions might in fact be difficult to engage in, especially given the requirements of focusing on the small laptop screen, the physical and social disconnect compounded by internet connectivity issues, and the need to follow remotely delivered instructions.

**Caveats and pitfalls identified by stakeholders:**

- Some service users might take time to become comfortable with a clinician and do not feel comfortable with extended small talk during the first sessions. Clinicians should employ therapeutic judgement to assess how to best approach developing a therapeutic relationship.
- Staff appears to generally more concerned about the therapeutic relationship than service users. However, this might be since service users are not able to compare to other service users' sessions and therapeutic relationships and thus, have less insight into what the "norm" is.

**Additional sources utilised:**


- Hubley S, Lynch SB, Schneck C, Thomas M, Shore J. Review of key telepsychiatry outcomes. World Journal of Psychiatry. 2016;6(2):269-82.
- LeRouge CM, Garfield MJ, Hevner AR. Patient perspectives of telemedicine quality. Patient Preference and Adherence. 2015;9:25-40.
- Lodder A, Papadopoulos C, Randhawa G. Using a blended format (videoconference and face to face) to deliver a group psychosocial intervention to parents of autistic children. Internet Interventions. 2020;21:100336.
- Lopez A, Schwenk S, Schneck CD, Griffin RJ, Mishkind MC. Technology-based mental health treatment and the impact on the therapeutic alliance. Current Psychiatry Reports. 2019;21(8):1-7.
- Olwill C, Mc Nally D, Douglas L. Psychiatrist experience of remote consultations by telephone in an outpatient psychiatric department during the COVID-19 pandemic. Irish Journal of Psychological Medicine. 2021;38(2):132-9.
- Pinto RZ, Ferreira ML, Oliveira VC, Franco MR, Adams R, Maher CG, et al. Patient-centred communication is associated with positive therapeutic alliance: A systematic review. Journal of Physiotherapy. 2012;58(2):77-87.


#### CMO 4.2: Assessment via telemental health vs face-to-face

When using telemental health for assessments (C), staff report finding it more difficult to ascertain diagnoses, assess mental health problems, care needs and/or risk (O), as they are less able to observe non-verbal and visual cues (depending on the telemental health modality used) and some service users may find it more difficult to have in-depth conversations about their problems and experiences (M).

**Underlying CMOs:**

- Bambling et al. (2008) Amongst counsellors providing online support services for young people (C) lack non-verbal feedback/ communication results in lower emotional intensity (M) which creates a risk of missing serious issues or suicidality or under-estimation of risk (O).
- Chen et al. (2020) Using telepsychiatry due to the Covid-19 pandemic in a general outpatient psychiatry service in the USA (C) resulted in some clinicians being less able to assess certain mental health markers such as hygiene/odour, gait, eye contact, and linguistic nuances (M), which potentially led to less effective mental health care (O).
- Grover et al. (2020b) When moving to telemental health (C) if staff experienced certain challenges such as experiencing difficulties in diagnosing problems while providing care through teleconsultations (M) associated with low staff satisfaction with the care they were providing (O).
- Grover et al. (2020a) When moving to telemental health (C) if staff experienced certain challenges such as difficulties in diagnosing problems (M) this was associated with a low level of satisfaction with care provided (O).
- Khanna et al. (2020) For a staff member (registrar) aiming to conduct assessments via telephone for a service user with homicidal ideation (in community mental health teams and/or outpatient services for individuals with PTSD) (C), when there was a lack of ability to see the consumer in order to facilitate a good mental state examination over the phone (M), then this led to challenges evaluating risk (O).
- Martin et al. (2020) Within early intervention for young people services which integrated digital communications such as text and email into contact with service users (C), staff were not able to use non-verbal cues to assess risk (M), potentially reducing the accuracy of risk assessments (O).
- Sehlo et al. (2021) Among service users of an online therapy site in Egypt during Covid-19 (C), a major advantage (O) of face-to-face consultations was a feeling that the doctor can diagnose more accurately (M).
- Uscher-Pines et al. (2020a) When moving to remote care (C) if remote care reduced the ability to observe nonverbal cues (including extrapyramidal symptoms from antipsychotics) (M) then this resulted in a lack of information for diagnosis or treatment (O).
- Uscher-Pines et al. (2020a) When moving to remote care (C) IF this led to 1) decreased clinical data for assessment (physical exam) 2) diminished patient privacy 3) increased distraction in the patient’s home setting (M) then this affected the quality of provider-patient interactions (O).
- Uscher-Pines et al. (2020b) When a service for patients with opioid disorder experiences switches to remote care (C) if staff find it more difficult to pick up on non-verbal cues using remote care compared to face-to-face (M) then this makes it more challenging to detect lies about drug use or diversion (O1) and clinicians have to ask more questions about physical symptoms and rely more on self-report (O2).
- Wyler et al. (2021) Among patients receiving telephone sessions (C) when there was a lack of access to body language and facial expression cues (M) then staff had limited insight into patients’ affective or emotional reactions (O).

**Supporting information from stakeholder consultation:**

- Assessments are more difficult due to a lack of non-verbal and visual cues and having to fully rely on self-reports of service users.
- Physical health checks are particularly impacted by telemental health.
- Staff might miss crucial information when they are not able to assess a person in their home. This limits a holistic approach.
- This may be especially applicable for service users who experience domestic violence and abuse (DVA) as they may not be able to be honest about their wellbeing and current situation in the presence of their abuser.

**Caveats and pitfalls identified by stakeholders and literature:**

- Staff may overestimate their ability to interpret the body language of service users, particularly of neurodivergent service users (Milton, 2012).
- Staff may be more prone to misinterpreting service users' body language and discomfort when using telemental health and thus make incorrect assumptions about service users' mental health.
- Staff should not (in any context) give more significance to external indicators than service user reports: clinicians can over-estimate their ability to read people's state of mind from non-verbal cues.
- Different non-verbal and visual cues, such as eye contact or extrapyramidal symptoms are not necessarily of equal importance and depend on the service user.
- It might be easier to assess service users when they are in their home environment, especially children and young people.
- If a person is at high-risk staff should offer to see them in person, e.g. at a clinic or at home.
- Service users might unconsciously or consciously present themselves differently via telemental health, e.g., speaking with a different voice on the phone or turning off their camera to conceal their current state. This might also be the case when service users can be overheard by others.

**Additional sources utilised:**


- Milton DE. On the ontological status of autism: the 'double empathy problem'. Disability & Society. 2012;27(6):883-7.


#### CMO 4.3: Staff support and training

When staff receive specific instructions as well as training, for example, on how to build rapport using telemental health and support from colleagues with prior telemental health experience (C), this facilitates good quality of care (O1), building therapeutic relationships (O2), and increased engagement (O3), as staff are able to ask questions and acquire new skills and knowledge about the interventions and thus build confidence in delivering telemental health (M).

**Underlying CMOs:**

- Conn et al. (2013) Amongst clinicians at a telepsychiatry service for older adults in rural Canada (C), healthcare professionals reported that the use of educational seminars had telepsychiatry (M) has made it possible for them to expand their repertoire of skills and improve patient care (e.g. by being able to provide them with more information) (O).
- Disney et al. (2021) During the rapid switch to telemental health (C), refugee mental health providers benefitted from training and checking in with colleagues who had prior telemental health experience (M), which enabled them to acquire new skills (O).
- Lindsay et al. (2015) Therapists who were trained by an external facilitator in order to introduce psychotherapy for PTSD via video telehealth in Veterans Affairs medical centres (C), deemed the regular calls with the facilitator very important in establishing video telehealth (O), as it provided an opportunity to discuss technical, logistical, and clinical issues specific to delivery of evidence-based psychotherapy via video telehealth and foster communication (M).
- Lindsay et al. (2015) Therapists who were trained by an external facilitator in order to introduce psychotherapy for PTSD via video telehealth in Veterans Affairs medical centres (C), deemed the regular calls with the facilitator as helpful in establishing video telehealth services (O), due to the interpersonal skills of the highly skilled facilitators (e.g., understanding, support, motivation, good communication skills) (M).
- Simon et al. (2021) NHS employees involved in intervention commissioning and implementation felt (C) that guided intervention such as specific instructions and opportunity to ask questions (M) improved treatment engagement (O).

- Simon et al. (2021) Among NHS commissioners involved in telemental health commissioning and implementation it was thought that NHS clinicians had (C); limited knowledge about the different interventions and who they were aimed at helping (M) and this was one barrier to internet-based therapy adoption (O).

**Supporting information from stakeholder consultation:**

- Staff should receive training on how to deliver care via telemental health beyond technology training and IT support. This includes training on how to develop therapeutic relationships and deal with the lack of non-verbal and visual cues. Training programmes should be adapted to specific service user groups.

**Caveats and pitfalls identified by stakeholders:**

- Services might assume that staff is confident in delivering telemental health care based on technological knowledge while this might not the case. Thus, service should provide support and training more widely.

#### CMO 4.4a: Service users finding it easier to establish a therapeutic relationship online

When delivering telemental health to some services users who feel uncomfortable in clinical settings and social situations (C), these service users find it easier to build a therapeutic relationship and are more willing to use telemental health (O), as they feel safer, are more relaxed and less anxious being in their own environment and/or outside of clinical settings and in-person social situations and thus feel more empowered and comfortable to open up and speak freely (M).

**Underlying CMOs:**

- Bambling et al. (2008) For young people receiving counselling online (C) the distance from the counsellor and being in their own home creates emotional safety for the young person (M) therefore they are less resistant to being open and are likely to communicate sensitive issues faster (O).
- Bambling et al. (2008) Amongst staff providing online video or text counselling for young people (C) the online mode was less emotionally intense that face-to-face (M) making it easier to communicate about complex issues (O - could also be M) and safer for both the client and counsellor (O).
- Bierbooms et al. (2020) Mental health professionals delivering telemental health (C) reported that service users being able to have appointments in their own homes (M) led to more open communication between service users and professionals (O).
- Holland et al. (2020) For one couple (one trans client with their husband) attending therapy online (C), meant that both the clients and the therapist felt that the couple were more relaxed because they were in their own environment (M1) enabling them to speak more freely (M2), the clients said the online sessions worked well (O).
- King et al. (2006) For young people engaging with text-based counselling (C) the lack of personal contact can feel less confronting than the traditional counselling format (M) therefore helps encourage engagement (O).
- Lodder et al. (2020) When delivering a Zoom online intervention to parents of children with autism (C) one participant found it easier to participate and talk to others in the intervention group (O) as they said they weren't very good at talking out loud in front of people (M).
- Nissling et al. (2020) Among people undergoing primary care internet-mediated CBT in Sweden, with peer support provided via phone calls and asynchronous text messages (C), when the digital

nature of the contact removed pressure normally felt in social settings (M) then it was easier to open up to the peer support worker (O).

- Ogueji et al. (2021) Among one participant from general population in Nigeria during COVID-19 (C), as she felt she would be more comfortable speaking to a therapist to who is not physically sat next to her and would feel less awkward (M), there was increased willingness to attend e-therapy (O).
- Smith and Gillon (2021) Therapists engaged in online counselling (C) suggested that working within a patient's living environment (M) led to a greater sense of empowerment for the client (O).

**Supporting information from stakeholder consultation:**

- Some service users are more comfortable and relaxed when using telemental health from home. This can help them engage and build a therapeutic relationship.
- Service users with social anxiety might prefer using telemental health.
- Clinic settings which may be upsetting for children and young people due to having to travel and being surrounded by others who have severe mental health problems can be avoided when using telemental health.

**Caveats and pitfalls identified by stakeholders and literature:**

- Telemental health might exacerbate safety behaviours of service users with social anxiety as they can avoid in person encounters. This may maintain and potentially exacerbate their social anxiety (McManus, Sacadura and Clark, 2008; Rapee and Heimberg, 1997).
- Face-to-face care should continue to be the default in terms of crisis care (when moving away from the pandemic).
- It can be difficult to engage children living in "chaotic home environments" or children with ADHD.

**Additional sources utilised:**


- McManus F, Sacadura C, Clark DM. Why social anxiety persists: An experimental investigation of the role of safety behaviours as a maintaining factor. Journal of Behavior Therapy and Experimental Psychiatry. 2008;39(2):147-61.
- Rapee RM, Heimberg RG. A cognitive-behavioral model of anxiety in social phobia. Behaviour Research and Therapy. 1997;35(8):741-56.


#### CMO 4.4b: Service users finding it easier to establish a therapeutic relationship via video vs phone

When service users and staff who prefer video calls use them (instead of telephone calls or text-based chats) for telemental health (C), this can facilitate a stronger therapeutic relationship (O1), satisfaction (O2) and engagement (O3), as it is easier to see visual and non-verbal cues, gauge the therapist's reaction, and connect with the service user/staff member compared to other telemental health modalities (M).

**Underlying CMOs:**

- Buckman et al. (2021) When delivering care remotely (C), clinicians reported that using video rather than telephone made it easier to see non-verbal cues and connect with the service user (M), which led to a stronger therapeutic relationship (O).

- Choi et al. (2014) Older adults who received problem solving therapy (C) stated a preference for videoconferencing over telephone appointments (O) as this meant that sessions felt more similar to face-to-face sessions (M).
- Costa et al. (2021) During the Covid-19 pandemic in the USA (C), those mental health service users who said that they were coping well during the pandemic (O), tended to report being able to see the same provider through video sessions (M).
- Eagle (2020) Amongst service users using remote technology for therapy (C) there was a preference for video over telephone modalities (O) as video calls allowed for non-verbal communication and cues similar to face-to-face therapy which weren't present in telephone therapy (M).
- Hawke et al. (2021) Young people accessing mental health services (C) who can see their providers face and are able to connect in human ways (M) engage better and have a preference for video calls over other platforms/ mediums of care e.g. online chat (O).
- Lichstein et al. (2013) Among older adults mental health service users who were receiving remote CBT via Skype for insomnia and depression (C), as they could see the therapist over Skype (M), preferred Skype over telephone, but would still overall prefer in-person treatment (O).
- Vera San Juan et al. (2021) Among mental health service users who had initial assessments over the phone during COVID-19 (C), they found it difficult to convey how distressed they were (M), and then felt this negatively affected the care they were offered afterwards (O).
- Severe et al. (2020) Among pediatric patients receiving telemental health care (C) the visual element of video calls was engaging (M) meaning that children preferred video formats to telephone calls (O).

**Supporting information from stakeholder consultation:**

- Some service users and clinicians are more comfortable using video calls for telemental health sessions as it allows observation of non-verbal and visual cues compared to phone calls.
- Some service users find it easier to build a therapeutic relationship via video calls compared to phone calls.
- Holding a phone can make it difficult to engage in telemental health sessions.

**Caveats and Pitfalls identified by stakeholders:**

- Service users' preference for video, phone, or face-to-face session can depend on the function of these sessions, e.g. relational vs non-relational purposes.
- Some service users are uncomfortable and self-conscious seeing themselves on the screen.
- Video calls and camera use can restrict movement and thus negatively impact engagement.

#### CMO 4.4c: Service users finding it easier to establish a therapeutic relationship via the phone vs face-to-face or video-calls

When services offer phone calls and text messages instead of video calls (C) some service users were more satisfied with their care (O) as they did not have to sit still and see themselves on screen, were less conscious/aware of their body language and facial gestures, were less distracted by non-verbal cues of the clinician, were able to move around freely, and thus less inhibited and able to open up more quickly (M).

**Underlying CMOs:**

- Costa et al. (2021) During the Covid-19 pandemic in the USA (C), some of those mental health service users who said that they were coping well during the pandemic (O), reported that they met with their providers over the telephone instead of video sessions (M), but that this did not pose much of a problem (O).

- Greenwood et al. (2004) Where there is limited provision for psychiatry in rural and remote locations and telepsychiatry services are offered (C), some SUs report not feeling comfortable in front of the camera (30%) (M) which may lead to lower levels of patient satisfaction (O).
- McBeath et al. (2020) Among a mixed group of psychology service users receiving online psychotherapy (video-link platforms and telephone) during COVID-19 (C), if/when telemental health made clients less conscious/aware of their body language and facial gestures during online therapy (M) then clients were less inhibited, opening up more quickly, and bringing more material to session (O).
- Uscher-Pines et al. (2020a) Among older patients and those who were self-conscious about their appearance, had social anxiety disorder, lacked suitable devices or had only limited broadband connection (C) if the service offered different modalities (M) then these patients opted for phone calls (and thus were able to continue to receive care (inferred) (O).
- Uscher-Pines et al. (2020a) When offering telemedicine to patients who are self-conscious about video visits and don't want clinicians to see their home environment for the first time (C) if staff provide FAQs that explain how to change the backgrounds to not show the surroundings on Zoom (M) then this improves the quality of telemedicine visits (since it increases privacy and patients feel more comfortable (inferred) (O).

**Supporting information from stakeholder consultation:**

- Some service users are uncomfortable and self-conscious seeing themselves on the screen.
- Video calls can be distracting and lead to sensory overload due to visual cues for some service users, e.g. some neurodivergent service users. Thus, they find it easier to build a therapeutic relationship via the phone.
- Service users can fidget and move around freely when receiving telemental health via the phone.
- Phones are easier to use and less likely to lose connection compared to video calls which facilitates building a therapeutic relationship.

**Caveats and pitfalls identified by stakeholders and literature:**

- Service users' preference for video, phone, or face-to-face session can depend on the function of these sessions, e.g. relational vs non-relational purposes.
- Some service users are uncomfortable and self-conscious seeing themselves on the screen.
- Video calls and camera use can restrict movement and thus negatively impact engagement.

#### CMO 4.5: More frequent telemental health sessions plus text messages

When services adapt flexibly to service users' preferences regarding the pattern and frequency of telemental health sessions, including offering more frequent, shorter rather than infrequent, long sessions, and/or additional asynchronous text messages and calls to check in between sessions (C), this may lead to stronger therapeutic relationships (O1), increased engagement (O2), and improved quality of care (O3), as service users receive regular and more frequent support depending on their preference (M).

**Underlying CMOs:**

- Chen et al. (2020) Due to Covid-19, patients attending a general psychiatry outpatient service in the USA who were in crisis or undergoing medication titration (C) were able to receive more frequent but briefer meetings with their clinician using telemental health care (M), which led to improved care (O).

- Chong and Moreno (2012) Amongst Hispanic patients receiving video call-based care for depression (C), participants felt that 30 minutes was too short for sessions, and they needed time to adapt to virtual care (M), which led to low acceptability rates (O).
- Bierbooms et al. (2020) Telemental health due to Covid-19 (C) resulted in more flexibility of the time and place of appointments (M1), which in turn increased accessibility to treatment (O1), and more frequent, shorter consultations (M2) which adds to connectedness between client and therapist (O2).
- British Psychological Society (2020a) For staff providing counselling remotely via video calls should consider doing outreach between sessions (e.g. texts and calls) (C) as this improves engagement (O) as it shows the service users the staff member cares and reminds them about the time of the next session (M).
- Nissling et al. (2020) Among people undergoing internet-mediated CBT in primary care in Sweden, with peer support provided via phone calls and asynchronous text messages (C), when peer support workers responded flexibly to requests for more verbal/phone contact (M) then participants felt a stronger connection with their peer support worker (O).

**Supporting information from stakeholder consultation:**

- Choice and agency regarding the frequency, length, and platform of telemental health sessions are key for building therapeutic relationships.
- When new patients have frequent sessions at the beginning stage of receiving telemental health this can help reduce anxiety and facilitate developing trust. Once a suitable appointment style and pattern as well as a therapeutic relationship have been established, sessions can be less frequent depending on the service user's preferences and needs.
- Short sessions can increase concentration and are less exhausting which can lead to increased engagement. This might be especially the case for children and young people.
- More frequent sessions might be especially important for less stable service users.

**Caveats and pitfalls identified by stakeholders:**

- Some service users might prefer longer and less frequent sessions depending on the sessions' purpose.

#### CMO 4.6: Enhancing quality of care through use of telemental health enhancements

When clinicians make appropriate and personalised use of enhancements and extensions of telemental health (such as using chat, voice activation to instruct phones, SMS and other text-based messaging, online appointment schedules, screen sharing and apps accessed during sessions) (C), this can lead to success engaging in telemental health (O1) and broadening the range of strategies and interventions available during clinical meetings (O2), as these features made engaging with services easier, and provided a functional method useful for exchanging practical information, such as reminding service users about the date and purpose of an appointment, with less room for ambiguity and more creative methods of engagement (M).

**Underlying CMOs:**

- British Psychological Society (2020b) For staff providing counselling remotely via video calls (C) should consider doing outreach between sessions (e.g. texts and calls) (M) as this improves engagement (O) as it shows the service users the staff member cares and reminds them about the time of the next session (M).

- Jones et al. (2014) For caregivers of children with disruptive disorders from low-socioeconomic backgrounds taking part in behavioural parent training with technology-enhancement (C), enhancement via video feedback and text message reminders increase the therapist and program's connection to and support for the family between sessions (M), increasing the family's autonomy with implementing learnt skills in the home setting (O).
- Shklarski et al. (2021) Clinicians who conducted psychotherapy with children over Zoom who found this modality made it harder to engage them in therapy (when compared to face-to-face) (C) who were able to adapt care using share screen options on zoom to play a game or draw together (M) found young people could be successfully engaged in therapy (O).
- Vera San Juan et al. (2021) Among mental health service users utilising text-based communications for mental health care during COVID-19 (C), they found this functional method useful for exchanging practical information with less room for ambiguity (M), and then therefore preferred it as a method of scheduling appointments and requesting medication (O).
- Williams et al. (2017) Among one homeless participant with dyslexia who has anxiety and depression (C), as he could use google voice activation to instruct his phone (M1) then he was able to contact friends and family (M2), which improved his mood (O).
- Williams et al. (2017) Among homeless people with memory issues who struggled to remember appointments (C), as digital health services allowed them to view appointments online and receive digital prompts/reminders (M), then they felt that online services would improve their experience of health services (O).

**Supporting information from stakeholder consultation and literature:**

- Clinicians need to use telemental health platforms and extensions flexibly and creatively to engage children and young people, e.g. using interactive platforms.
- Adolescents, especially those with social anxiety or autism, benefit from, and at times prefer, the use of a chat function. It allows them to turn their camera off and respond at their own pace. There is evidence which supports this in other settings, for example, this was also observed among inpatients with complex and severe mental health problems when engaging with video telemental health during covid-19 lockdowns (Riches et al., 2021).
- Texting is central to the communication of young people. It can be used to prepare young people for a phone call or to check in with them between sessions.
- Communicating the purpose and content of the next telemental health session is crucial for telemental health contacts and can reduce anxiety and/or stress prior to telemental health sessions. Service users can also be sent reminders via text.
- Telemental health cannot be a solution for everything, e.g. art therapy, and treatment for traumatic stress.

**Caveats and pitfalls identified by stakeholders:**

- The use of telemental health features should be driven by patient choice.
- Some telemental health platforms lack features that facilitate engagement, e.g. screen sharing.
- telemental health features can enhance an online session well when they operate without issue, however they can cause significant frustration and anxiety when there are unexpected problems during the session.
- Engaging children and young people via telemental health successfully depends on the age group, e.g. children who are younger than 12-years-old or have special needs can be difficult to engage.
- Telemental health features do not provide a substitute for engaging very young children with toys in face-to-face sessions.

**Additional sources utilised:**

- Riches S, Azevedo L, Steer N, Nicholson S, Vasile R, Lyles S, et al. Brief videoconference-based dialectical behaviour therapy skills training for COVID-19-related stress in acute and crisis psychiatric staff. Clinical Psychology Forum. 2021:57-62.

#### CMO 4.7: Staff wellbeing and quality of care

When staff utilise the time saved on travel to take breaks in between telemental health sessions (C), this may increase staff wellbeing (O1) and improve quality of care (O2), as it provides the opportunity to reflect and recharge after telemental health sessions, which are often experienced as tiring, and thus reduces fatigue, tension, and anxiety among staff (M1), and staff can use some of the time on clinical work, catch up on administrative tasks, or engage in professional developmental activities (M2).

**Underlying CMOs:**

- Bommersbach et al. (2021) Using teleconferencing due to the Covid-19 pandemic (C) improved efficiency for mental health staff due to reduced travel (M), which led to more time for patients and their own professional development (O).
- Buckman et al. (2021) When delivering care using telemental health rather than face-to-face (C), clinicians reported finding delivering sessions via video (M) more tiring, draining, or a strain on the eyes, compared with delivering sessions face-to-face (O).
- Foye et al. (2020) Amongst mental health nurses (C), if they were pressured to have back-to-back appointments and thus more meetings or appointments in one day than previously expected due to working remotely and cutting back on travel time (M), then this added to workload pressures and feelings of burnout amongst staff (O).
- The Health Foundation (2020) Staff who are providing video consultations in remote areas (C) have more flexibility in their working day and have less travel (M) therefore there are fewer costs associated and staff are able to do more calls, later and quicker (O).
- Hopkins and Pedwell (2021) Among staff providing mental health care for young people during the pandemic (C) if they have flexibility to manage their work/life balance, consider logistics of life (e.g. reduced travel, managing childcare) and are given autonomy to work flexibly (M), then staff are more accepting of integrating telehealth approaches into their work (O).
- Liberati et al. (2021) In the move to remote care because of the pandemic, for staff who were providing remote consultations (C) if they had back-to-back appointments (M) then they (staff) had no opportunity to reflect or process after consultations (O).
- Mahmoud et al. (2021) Amongst telepsychiatrists providing telemental health care (C) where the job allows flexibility, no commuting and easy ways of working (M), staff are happier and feel they have a better work-life balance (O).
- McBeath et al. (2020) Among psychotherapists delivering online psychotherapy (video-link platforms and telephone) to mixed groups of psychology service users during COVID-19 (C) if they created spaces between sessions and engaged with fewer clients per day (M) THEN this led to an alleviation of tension and anxiety among the psychotherapists which was occurring due to increased intensity of online sessions (O).
- Shklarski et al. (2021) For clinicians who used Zoom for all their therapeutic appointments due to the covid-19 pandemic (C) if they were able to spread out their appointments and take breaks between them (M) then they were able to reduce Zoom fatigue (O).
- Wilson et al. (2021) When switching to remote care (C) if remote care led to a reduction in travel time for staff (M) then this helped with time management, increased time for clinical record keeping and improved work life balance (O).

**Supporting information from stakeholder consultation:**

| |
|---|
| • Video calls are draining and tiring, can cause eye strain and headaches, and people may need to concentrate more during video calls compared to face-to-face sessions.<br>• Travel time is often used to take a break between sessions, reflect on sessions or take notes. This is lost when staff have booked back-to-back telemental health sessions. |
| **Caveats and pitfalls identified by stakeholders:**<br>• Time gained through lack of travelling could be used to regroup instead of writing-up notes.<br>• Services should encourage staff to take breaks and not push staff to book back-to-back sessions. This is important for the wellbeing of staff as well as safety and confidentiality of sessions (taking time to process sessions).<br>• Wider context: The wider work culture of mental health services needs to promote staff wellbeing; services or staff might be resistance to change; organisational policies might hinder staff to work flexibly (using face-to-face care and telemental health).<br>• We are changing the way staff are working, including a lack of peer support for staff. If a risk situation comes up, there are always people around to call but it does not feel the same in terms of team connection, support and thinking. |
